# Hospital-wide, dynamic, individualized prediction of central line-associated bloodstream infections - development and temporal evaluation of six prediction models

**DOI:** 10.1101/2024.11.04.24316689

**Authors:** Elena Albu, Shan Gao, Pieter Stijnen, Frank E. Rademakers, Christel Janssens, Veerle Cossey, Yves Debaveye, Laure Wynants, Ben Van Calster

## Abstract

**Background:** Central line-associated bloodstream infections (CLABSI) are preventable hospital-acquired infections. Predicting CLABSI helps improve early intervention strategies and enhance patient safety.

**Aim:** To develop and temporally evaluate dynamic prediction models for continuous CLABSI risk monitoring.

**Methods:** Data from hospitalized patients with central catheter(s) admitted to University Hospitals Leuven between 2014 and 2017 were used to develop five dynamic models (a cause-specific landmark supermodel, two random forest models, and two XGBoost models) to predict 7-day CLABSI risk, accounting for competing events (death, discharge, and catheter removal). The models’ predictions were then combined using a superlearner model. All models were temporally evaluated on data from the same hospital from 2018 to 2020 using performance metrics for discrimination, calibration, and clinical utility.

**Findings:** Among 61629 catheter episodes in the training set, 1930 (3.1%) resulted in CLABSI, while in the test set of 44544 catheter episodes, 1059 (2.4%) experienced CLABSI.

Among individual models, one XGBoost model reached an AUROC of 0.748. Calibration was good for predicted risks up to 5%, while the cause-specific and XGBoost models overestimated higher predicted risks. The superlearner displayed a modest improvement in discrimination (AUROC up to 0.751) and better calibration than the cause-specific and XGBoost models, but worse than the random forest models. The models showed clinical utility to support standard care interventions (at risk thresholds between 0.5-4%), but not to support advanced interventions (at thresholds 15-25%). A deterioration in model performance over time was observed on temporal evaluation.

**Conclusion:** Hospital-wide CLABSI prediction models offer clinical utility, though temporal evaluation revealed dataset shift.

## Introduction

Central line-associated bloodstream infection (CLABSI), defined as a bacteraemia occurring at least 48 hours after admission in the absence of infection at another site^1^, is a priority for prevention due to its association with prolonged hospital stays, increased healthcare costs and elevated morbidity and mortality^2–4^. Developing a risk prediction model for CLABSI can assist healthcare professionals, including clinicians and infection control teams, with timely risk assessments to inform preventative interventions and improve patient outcomes.

A previous systematic review^5^ assessed existing risk prediction models for CLABSI and found that 15 out of 16 developed models were static, with only one dynamic model developed. Static models may be less effective, as they fail to capture the changes in a patient’s health throughout their hospital stay. The increasing availability of continuously updated electronic health records (EHR) datasets offers a new opportunity. As demonstrated by other longitudinal studies on EHR data for various clinical outcomes^6–8^, these datasets enable more frequent measurements that may better reflect the patient’s evolving condition or disease progression. By continuously updating patient information and renewing predictions, dynamic prediction becomes feasible, providing a more nuanced and timely assessment of the patient’s health status^9^.

We developed and temporally evaluated dynamic risk prediction models for capturing the 7-day risk of CLABSI among hospitalized patients with central venous catheters, utilizing EHR data from the University Hospitals Leuven. We developed six models: a cause-specific regression model, two random forest (RF), two eXtreme Gradient Boosting (XGB) models, and a superlearner ensemble.

## Methods

The study reporting follows the TRIPOD+AI statement^10^.

### Study design and data

Patient data were extracted from the EHR system of the University Hospitals Leuven for hospital admissions in the period January 2014 -December 2020. We included patient admissions with registration of central catheters: centrally inserted central catheter (CICC), tunnelled cuffed and non-cuffed central venous catheter, port catheter (Totally Implanted Vascular Access Devices -TIVAD), peripherally inserted central catheter (PICC) and dialysis catheter. No exclusion criteria were applied; all patients from all medical wards were included, from neonates to geriatric patients, including intensive care (ICU) admissions, reflecting the target population for a hospital-wide CLABSI alert system. The training set used for model development included hospital admissions starting between January 2014 and December 2017. The test set used for temporal evaluation included hospital admissions between January 2018 and December 2020. The data extraction and preparation procedure has been described in our previous studies conducted on data collected prior to January 2014^11,12^ and is repeated.

The following levels of the outcome are considered:

- **CLABSI**: any laboratory-confirmed bloodstream infection (LC-BSI) for a patient with central catheter or within 48 hours after the central catheter removal that is not present on admission, that is not secondary to another infection and is not a mucosal barrier injury LC-BSI. The CLABSI definition was applied retrospectively on the extracted data following the definition of the Belgian public health institute Sciensano, published in 2019^1^. While Sciensano does not impose a specific time window for secondary infections, we used 17 days, considering the infection window period of 14 days plus the repeat infection timeframe of 3 days^13^.
- **Discharge**: Hospital discharge or 48 hours after catheter removal, whichever happens first. According to the Sciensano definition, the patient remains at risk of CLABSI for 48 hours after catheter removal.
- **Death**: The first contact with palliative care during admission, transfer to palliative care or patient death, whichever happens first. Patients stop being closely monitored in palliative care and predictions on this ward are not actionable.

Patient admissions were split in catheter episodes. A catheter episode starts at catheter placement (if placed in ICU) or at the first structured catheter observation (e.g. monitoring observations like dressing status, lumens flushed, bandage change, etc.) during the admission (outside ICU). A catheter episode ends when no catheter observation is made for 48 hours (as a proxy for catheter removal) or at catheter removal, or when discharge, death or CLABSI occurs. A catheter episode was further split in landmarks (LMs) marking every 24 hours: LM0 is the time of the first catheter observation, LM1 is 24h later, and so on (Supplementary Material 1). The risk of CLABSI within the following 7 days is predicted at each landmark throughout the catheter episode.

The complete dataset consisted of 160 baseline and time-varying variables comprising patient demographics, ward transfers, medication, laboratory test results, comorbidities, vital signs and catheter registrations. They were selected out of 302 extracted variables based on a priori information, including insights from analyses on data prior to 2014, clinical expertise, and reliability of variables (i.e. expectation that the variable will be recorded consistently in future data). Whenever multiple measurements were available during a time window, these were aggregated into a single landmark value (e.g.: maximum temperature in last 24 hours, Supplementary Material 2).

Missing data were encountered for 66 variables and were imputed using missForestPredict^14,15^ on the training dataset using the complete set of variables. The outcome was not included in the imputation process. The test set was imputed using the imputation model learned on the training set. (Supplementary material 3 provides further information).

### Model development and evaluation

We built five different dynamic models that use time-varying predictor information and jointly model CLABSI, death, discharge and “no event” within 7 days as the outcome levels: a regression model, specifically a landmark cause-specific supermodel (CS), considering death and discharge as competing events for CLABSI, two multinomial RF models and two multinomial XGB models.

The CS model used a limited set of 62 variables, including interaction and polynomial terms (Supplementary Material 2). The limited set of variables was selected from the complete set of variables based on their predictiveness for CLABSI in data prior to 2014 (Supplementary Material 4). One RF and one XGB model were built on the same variables, excluding interaction and the polynomial terms, resulting in 58 variables (RF-LIM and XGB-LIM). Another RF and XGB model were built using the complete set of 160 variables with either explicit data-driven variable selection (RF-ALL) or implicit data-driven variable selection through regularization hyperparameters (XGB-ALL).

Further, we combined the predictions of the five models into a superlearner model, which previously demonstrated improved predictive performance^16,17^. We used ten-fold cross-validation (called outer CV) to parametrize the superlearner. Within the outer CV we performed nested 5-fold cross-validation for tuning the XGB models and used the out-of-bag samples for tuning the RF models. (Supplementary Material 5)

Internal evaluation was performed for the individual models (CS, RF and XGB) by evaluating performance metrics using the outer CV held-out folds. To assess the model performance over time, temporal evaluation was performed for all models including the SL model on the test set (2018-2020). Threshold-independent metrics for discrimination (Area Under the ROC curve) and calibration (assessing how well the predicted CLABSI probabilities align with the observed outcomes) are reported at each landmark in a catheter episode. Calibration plots over all landmarks are presented. Threshold-dependent metrics (net benefit, sensitivity, specificity, positive predictive value, negative predictive value and alert rate) were evaluated on temporal evaluation, for all landmarks combined. We considered two risk thresholds for generating alerts: a medium-risk threshold for which the nursing team can intervene with standard care practice (standard care bundle with enhanced routine maintenance check) and a high-risk threshold for enhanced interventions for additional protection (e.g.: chlorhexidine-impregnated dressings and washcloths or chlorhexidine-coated catheters). Following consensus of different representatives of the model’s users (nurses, clinicians and infection preventionists), for the medium-risk threshold we considered the values: 0.5%, 1%, 2%, 3%, 4%, 5% and for the high-risk threshold 15%, 20%, 25%. Decision curves^18,19^ were also presented to assess clinical utility of intervention decisions at the proposed thresholds.

## Results

### Data characteristics

The training set consisted of 55910 admissions, 61628 catheter episodes with complete follow up for all event types (1930 CLABSI, 3206 deaths and 56492 discharges) and 541815 landmarks. The test set consisted of 40994 admissions, 44544 catheter episodes (1059 CLABSI, 1901 deaths and 41584 discharges) and 391353 landmarks. Supplementary material 6 presents the outcome prevalence and incidence. Table 1 presents the patient demographics and catheter information at baseline.

**Table 1:** Patient characteristics at baseline (start of the catheter episode); median (IQR) for continuous variables, after missing data imputation; n (%) for categorical variables. Patients can have two or more different catheter types simultaneously, therefore catheter type has been coded as a binary rather than a categorical variable with multiple categories. IQR = interquartile range; F = female; M = male; CLABSI = Central Line-Associated Bloodstream Infection; ICU = Intensive Care Unit; ORL = Otorhinolaryngology; CICC = Centrally Inserted Central Catheter; CVC = Central Venous Catheter; PICC = Peripherally Inserted Central Catheter; TIVAD = Totally Implanted Vascular Access Devices; tc-CICC = tunnelled cuffed CICC; t-CICC = tunnelled CICC. UNKNOWN_VALUE = The paediatric ward has changed name ^*^ done in training data did not cover the values in the test data. The emergency ward started using the EHR system consistently for catheter registrations in 2018. ICU, Abdomen and Gynecology are less prevalent at baseline (as entry-wards) in the test set, while emergency and cardiac are more prevalent; over all landmarks, the number of landmarks per ward remains rather stable (Supplementary Material 2)

| Variable | Statistic | Train | Test | Total |
| --- | --- | --- | --- | --- |
|  |  | n = 61628 | n = 44544 | n = 106172 |
| Admission ward | Abdomen, n(%) | 9944 (16.1) | 5553 (12.5) | 15497 (14.6) |
|  | Cardiac, n(%) | 3096 (5.0) | 3608 (8.1) | 6704 (6.3) |
|  | Emergency, n(%) | 2063 (3.3) | 3818 (8.6) | 5881 (5.5) |
|  | Endocrinology, n(%) | 421 (0.7) | 239 (0.5) | 660 (0.6) |
|  | Geriatrics, n(%) | 745 (1.2) | 561 (1.3) | 1306 (1.2) |
|  | Gynecology, n(%) | 3474 (5.6) | 4616 (10.4) | 8090 (7.6) |
|  | Hematology, n(%) | 2535 (4.1) | 1822 (4.1) | 4357 (4.1) |
|  | ICU, n(%) | 10620 (17.2) | 4259 (9.6) | 14879 (14.0) |
|  | Internal Medicine, n(%) | 554 (0.9) | 392 (0.9) | 946 (0.9) |
|  | Neonatology, n(%) | 1456 (2.4) | 1100 (2.5) | 2556 (2.4) |
|  | Nephrology, n(%) | 646 (1.0) | 508 (1.1) | 1154 (1.1) |
|  | Neuro, n(%) | 1255 (2.0) | 996 (2.2) | 2251 (2.1) |
|  | ORL, n(%) | 503 (0.8) | 450 (1.0) | 953 (0.9) |
|  | Oncology, n(%) | 8600 (14.0) | 5749 (12.9) | 14349 (13.5) |
|  | Other, n(%) | 84 (0.1) | 59 (0.1) | 143 (0.1) |
|  | Pediatrics, n(%) | 4331 (7.0) | 1051 (2.4) | 5382 (5.1) |
|  | Pneumology, n(%) | 4335 (7.0) | 2804 (6.3) | 7139 (6.7) |
|  | Thoracic Surgery, n(%) | 1940 (3.1) | 1359 (3.1) | 3299 (3.1) |
|  | Transplant, n(%) | 1073 (1.7) | 759 (1.7) | 1832 (1.7) |
|  | Traumatology, n(%) | 2601 (4.2) | 1545 (3.5) | 4146 (3.9) |
|  | UNKNOWN_VALUE, n(%) | 0 (0.0) | 2642 (5.9) | 2642 (2.5) |
|  | Urology, n(%) | 1352 (2.2) | 654 (1.5) | 2006 (1.9) |
| CLABSI history | n(%) | 1061 (1.7) | 591 (1.3) | 1652 (1.6) |
| Catheter type CICC | n(%) | 25560 (41.5) | 15893 (35.7) | 41453 (39.0) |
| Catheter type PICC | n(%) | 6789 (11.0) | 5965 (13.4) | 12754 (12.0) |
| Catheter type TIVAD | n(%) | 25257 (41.0) | 19083 (42.8) | 44340 (41.8) |
| Catheter type tc-CICC or t-CICC | n(%) | 2789 (4.5) | 2690 (6.0) | 5479 (5.2) |
| Dialysis_CVC | n(%) | 1637 (2.7) | 1182 (2.7) | 2819 (2.7) |
| Patient age at admission | Median (IQR) | 61.0 (46.0 to 71.0) | 62.0 (46.0 to 72.0) | 61.0 (46.0 to 71.0) |
|  | Range (min, max) | (0 to 101) | (0 to 100) | (0 to 101) |
| Patient sex | F, n(%) | 28723 (46.6) | 22178 (49.8) | 50901 (47.9) |
|  | M, n(%) | 32905 (53.4) | 22366 (50.2) | 55271 (52.1) |
| Unplanned readmission | n(%) | 9610 (15.6) | 7918 (17.8) | 17528 (16.5) |
| Days to event | Median (IQR) | 5.0 (2.3 to 9.8) | 5.0 (2.1 to 10.0) | 5.0 (2.2 to 9.9) |
|  | Range (min, max) | (0 to 298) | (0 to 255) | (0 to 298) |

### Models evaluation

Considering the individual models, the XGB-ALL model had the highest AUROC on temporal evaluation, reaching a maximum of 0.748 at landmark 8 (i.e. 8 days after the start of the catheter episode), while the cause-specific model has the lowest AUROC. The RF-ALL model selected slightly less variables compared to the RF-LIM model yet had slightly better performance (45 versus 58 variables out of which 25 are common). The XGB-ALL model did not perform variable selection. The performance decreased with each year on the test set. In Supplementary Material 7 we present the models evaluated on each year of the test set: 2018, 2019 and 2020.

The calibration plot showed agreement between observed and predicted risks for all models at low predicted risk levels (lower than 0.05). The cause-specific and XGBoost models produced overly extreme estimates at high predicted risk levels (Figure 2). The E:O ratios higher than one indicate that the models tend to overestimate the CLABSI risk. The E:O ratio for all models temporally evaluated on each year also showed an increasing trend over year, which suggested more severe overestimation at later years in the test data (Supplementary Material 7).

**Figure 1.**
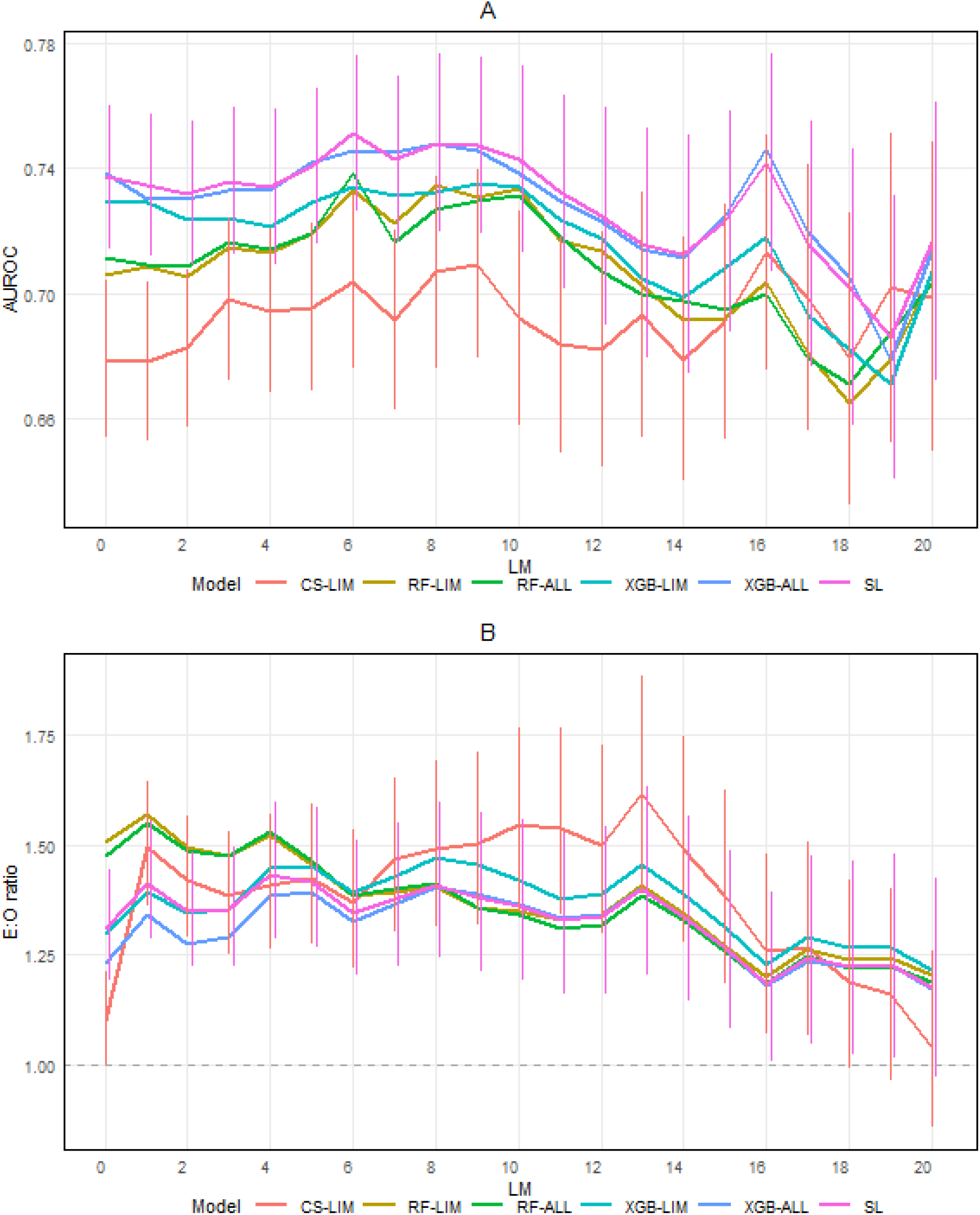
Prediction performance at each landmark on the test. AUROC = Area Under the ROC curve; E:O = Expected:Observed. The vertical bars represent confidence intervals; for better visualisation, these are shown only for two models: CS-LIM and SL. The confidence interval bars for the SL model are shifted to the right with a small offset for better visualisation.

**Figure 2.**
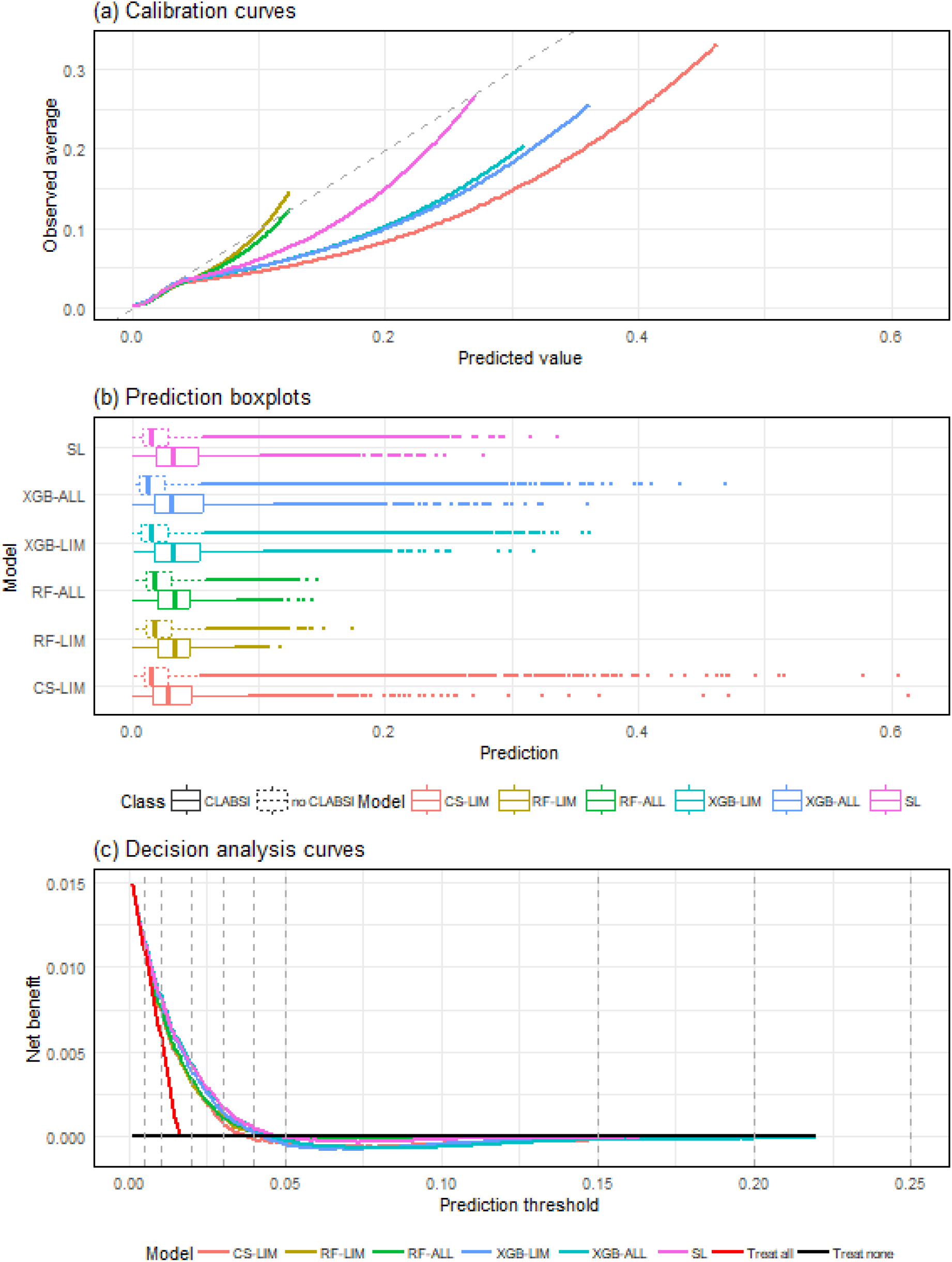
Calibration curves using restricted cubic splines (a); prediction boxplots (b); decision analysis curves (c) for all models on temporal evaluation using all landmarks. (a) The dashed grey line represents the identity function (perfect calibration). (c) The dashed grey lines mark the proposed thresholds for model evaluation.

The superlearner model showed a minimal improvement in discrimination: maximum AUROC of 0.751 at landmark 6. It also displayed better calibration than the cause-specific and XGB models, but its calibration was still inferior to that of the RF models. (Figure 2). The proportional contributions of each model in the superlearner were: 0.02 for CS, 0.06 for RF-LIM, 0.27 for RF-ALL, 0 for XGB-LIM and 0.65 for XGB-ALL.

The models showed no benefit to support specific technological interventions (high-risk thresholds 15%-25%), but they had clinical utility to support standard care interventions for medium-risk thresholds up to 3% for the CS model and up to 4% for the other models (Figure 2 (c) and Table 2). Nonetheless, the utility of the models differed by ward. For example, the superlearner had utility for all (0.5%-5%) medium-risk thresholds in haematology, traumatology, gynaecology, neonatology, thoracic surgery, neurology, internal medicine, and urology (Supplementary Material 7), while it depended on the specific threshold in other wards.

**Table 2:** Threshold dependent metrics at different thresholds. The Net Benefit has been multiplied by 100 and represents the Net true positives per 100 landmarks (catheter-days). For Net Benefit, * indicates values that exceed both the net benefit of the ‘treat all’ intervention and the net benefit of the ‘treat none’ intervention, -0 indicates small values but negative. PPV = Positive Predictive Value; NPV = Negative Predictive Value; NB = Net Benefit.

| Model | metric | 0.005 | 0.01 | 0.02 | 0.03 | 0.04 | 0.05 | 0.15 | 0.2 | 0.25 |
| --- | --- | --- | --- | --- | --- | --- | --- | --- | --- | --- |
| CS-LIM | 100 NB | 1.11* | 0.74* | 0.32* | 0.08* | -0.02 | -0.05 | -0.03 | -0.01 | -0.01 |
| RF-LIM | 100 NB | 1.13* | 0.76* | 0.31* | 0.11* | 0.03* | -0.01 | -0 | 0 | 0 |
| RF-ALL | 100 NB | 1.13* | 0.76* | 0.34* | 0.11* | 0.02* | -0.02 | 0 | 0 | 0 |
| XGB-LIM | 100 NB | 1.14* | 0.8* | 0.39* | 0.14* | 0.02* | -0.05 | -0.02 | -0.01 | -0 |
| XGB-ALL | 100 NB | 1.15* | 0.83* | 0.43* | 0.17* | 0.05* | -0.03 | -0.02 | -0.02 | -0 |
| SL | 100 NB | 1.14* | 0.82* | 0.42* | 0.18* | 0.05* | -0.01 | -0.01 | -0 | -0 |
| CS-LIM | Sensitivity | 0.99 | 0.92 | 0.67 | 0.48 | 0.33 | 0.22 | 0.01 | 0 | 0 |
| RF-LIM | Sensitivity | 0.99 | 0.96 | 0.77 | 0.57 | 0.36 | 0.18 | 0 | 0 | 0 |
| RF-ALL | Sensitivity | 1 | 0.96 | 0.76 | 0.56 | 0.37 | 0.19 | 0 | 0 | 0 |
| XGB-LIM | Sensitivity | 0.99 | 0.91 | 0.7 | 0.52 | 0.39 | 0.28 | 0.02 | 0 | 0 |
| XGB-ALL | Sensitivity | 0.97 | 0.89 | 0.7 | 0.51 | 0.39 | 0.29 | 0.03 | 0.01 | 0 |
| SL | Sensitivity | 0.99 | 0.93 | 0.73 | 0.54 | 0.38 | 0.27 | 0.01 | 0 | 0 |
| CS-LIM | Specificity | 0.06 | 0.27 | 0.63 | 0.78 | 0.87 | 0.92 | 1 | 1 | 1 |
| RF-LIM | Specificity | 0.08 | 0.22 | 0.55 | 0.74 | 0.87 | 0.94 | 1 | 1 | 1 |
| RF-ALL | Specificity | 0.08 | 0.23 | 0.57 | 0.74 | 0.86 | 0.94 | 1 | 1 | 1 |
| XGB-LIM | Specificity | 0.14 | 0.35 | 0.63 | 0.77 | 0.85 | 0.9 | 1 | 1 | 1 |
| XGB-ALL | Specificity | 0.2 | 0.41 | 0.66 | 0.79 | 0.86 | 0.91 | 1 | 1 | 1 |
| SL | Specificity | 0.12 | 0.33 | 0.63 | 0.78 | 0.86 | 0.92 | 1 | 1 | 1 |
| CS-LIM | PPV | 0.02 | 0.02 | 0.03 | 0.03 | 0.04 | 0.04 | 0.06 | 0.07 | 0.09 |
| RF-LIM | PPV | 0.02 | 0.02 | 0.03 | 0.03 | 0.04 | 0.05 | 0 | NaN | NaN |
| RF-ALL | PPV | 0.02 | 0.02 | 0.03 | 0.03 | 0.04 | 0.05 | NaN | NaN | NaN |
| XGB-LIM | PPV | 0.02 | 0.02 | 0.03 | 0.04 | 0.04 | 0.05 | 0.09 | 0.09 | 0.06 |
| XGB-ALL | PPV | 0.02 | 0.02 | 0.03 | 0.04 | 0.04 | 0.05 | 0.1 | 0.1 | 0.17 |
| SL | PPV | 0.02 | 0.02 | 0.03 | 0.04 | 0.04 | 0.05 | 0.1 | 0.2 | 0.06 |
| CS-LIM | NPV | 1 | 1 | 0.99 | 0.99 | 0.99 | 0.99 | 0.98 | 0.98 | 0.98 |
| RF-LIM | NPV | 1 | 1 | 0.99 | 0.99 | 0.99 | 0.99 | 0.98 | 0.98 | 0.98 |
| RF-ALL | NPV | 1 | 1 | 0.99 | 0.99 | 0.99 | 0.99 | 0.98 | 0.98 | 0.98 |
| XGB-LIM | NPV | 1 | 1 | 0.99 | 0.99 | 0.99 | 0.99 | 0.98 | 0.98 | 0.98 |
| XGB-ALL | NPV | 1 | 1 | 0.99 | 0.99 | 0.99 | 0.99 | 0.98 | 0.98 | 0.98 |
| SL | NPV | 1 | 1 | 0.99 | 0.99 | 0.99 | 0.99 | 0.98 | 0.98 | 0.98 |
| CS-LIM | Alert rate | 0.94 | 0.74 | 0.38 | 0.23 | 0.13 | 0.08 | 0 | 0 | 0 |
| RF-LIM | Alert rate | 0.92 | 0.78 | 0.46 | 0.27 | 0.14 | 0.06 | 0 | 0 | 0 |
| RF-ALL | Alert rate | 0.92 | 0.78 | 0.44 | 0.26 | 0.14 | 0.06 | 0 | 0 | 0 |
| XGB-LIM | Alert rate | 0.86 | 0.65 | 0.37 | 0.23 | 0.15 | 0.1 | 0 | 0 | 0 |
| XGB-ALL | Alert rate | 0.81 | 0.59 | 0.34 | 0.22 | 0.14 | 0.1 | 0 | 0 | 0 |
| SL | Alert rate | 0.88 | 0.67 | 0.38 | 0.23 | 0.14 | 0.09 | 0 | 0 | 0 |

The sensitivity, specificity and alert rate at medium-risk thresholds varied widely; the PPV remained below 5% regardless of threshold. The highest ward-specific alert rates at medium-risk thresholds are reached at threshold 0.5% in emergency (99.1%) and ICU (98.9%) and the lowest at threshold 5%: 0.2% in traumatology and gynaecology.

Among the individual models, XGB-ALL had the best discrimination and the best net benefit at medium-risk thresholds. The superlearner showed slightly better discrimination and calibration than XGB-ALL, and superior net benefit at thresholds 3% and 4%, but slightly lower net benefit at the other three thresholds (0.5%, 1%, 2%). From a practical perspective, the superlearner comes with the disadvantage of deploying four models (considering that XGB-LIM got zero weight). Both models include the same number of variables, as XGB-ALL did not perform variable selection.

### Temporal shift

All models showed a performance deterioration on temporal evaluation (test set) compared to the internal evaluation (Supplementary Material 7). For example, the median CV AUROC for XGB-ALL reached the maximum value of 0.774 at landmark 3,.

To understand the reduced performance in temporal evaluation, we first analysed predictor shifts across training and test data (Supplementary Material 8). Fluctuations in missing data and variable sparsity were observed across years. Additionally, significant shifts in log(D-dimer) distributions negatively impacted temporal evaluation. A lab machine upgrade in May 2019 expanded the maximum measurable D-dimer range from 7,650 (training) to 284,780 (test), leading to inflated predictions for patients with extremely high D-dimer levels. Removing log(D-dimer) from the landmark cause-specific supermodel improved overall AUC and calibration, making it comparable to the XGB models’ calibration.

Furthermore, we analysed the outcome prevalence shift across training and test data (Supplementary Material 8). A declining trend in CLABSI cases was observed across medical specialties like geriatrics and thoracic surgery, along with a continuous hospital-wide decrease in outcome prevalence, despite relatively stable counts of positive cultures and catheter episodes. This reflects improvements in clinical care, likely drive by the gradual implementation of the line care bundle from 2012 to full hospital-wide adoption by 2016.

To assess the impact of shifts in predictor-outcome associations, we fitted landmark cause-specific supermodels using the limited set of variables on yearly data from 2014 to 2020 (Supplementary Material 8). Results showed lower apparent AUCs for 2019 and 2020 at the first six landmarks compared to other years, particularly the apparent AUCs for 2019 worsened after landmark nine. As training the model on test data did not improve the ssprediction model’s performance, these results suggest the decreasing performance reflect complex changes in the evolving clinical environment that made the prediction problem harder, rather than the issues with the model. Due to time and computerization constraints, only the landmark cause-specific models were fitted on the full training and test data. XGBoost models on the full dataset may be explored in future studies.

## Discussion

We developed and evaluated dynamic models for predicting the 7-day risk of CLABSI in patients with central catheters using EHR data. XGBoost models showed the best discrimination and random forest models showed the best calibration. Combining multiple models through a superlearner framework marginally improved discrimination and partially corrected the miscalibration of XGBoost. Temporal evaluation revealed a decrease in model performance compared to internal evaluation. The CLABSI incidence has also decreased over time, which is reflected in the increasing overestimation of predicted risks over time.

The CLABSI models have practical relevance when implemented in an early warning system targeting users like nurses and clinicians on different wards, as well as infection preventionists and the Vascular Access Specialty Teams who oversee the care of all hospitalized patients. We aimed for two implementation thresholds: medium-risk alerts, for which nurses can intervene by verifying the catheter maintenance care bundle and infection preventionists can check the adherence to the care bundle items, and high-risk alerts, which prompt for specific technological interventions (e.g.: dressing type, catheter replacement). The models demonstrated both hospital-wide and ward-specific utility for medium-risk alerts (up to a threshold of 3%), but not for high-risk alerts.

In comparison to previously published models^5^, which are mostly static, we developed and temporally evaluated dynamic models. While our systematic review identified one dynamic model^20^, it was limited to patients in the cardiac ICU or cardiac ward, whereas our models targeted a hospital-wide population. Additionally, their definition of CLABSI as any new positive blood culture was much broader than the surveillance definition of CLABSI. Moreover, previously published models were at high risk of bias, e.g. by exclusions based on future data, temporal data leakage, or using predictors unavailable at the time of prediction.

Our study has limitations. Although we performed a comprehensive data extraction, we did not particularly focus on extracting strong predictors for the competing events included in our models: death and discharge. Further refining the extracted predictors or variable selection strategy might lead to improved model performance. Second, we did not account for variable dynamics in the model. Even with dynamic predictions, a prediction at a given time relies only on the values at the prediction time, or in a fixed time window before the prediction time (typically of 24 hours) without incorporating previous values during a catheter episode (e.g.: values 48 hours before the prediction time) or trends for individual variables within a catheter episode (e.g.: increase / decrease over time in lab values). In a previous analysis on data prior to 2014, we tested models with two lagged values or with differences between values at consecutive landmarks, but these did not improve the performance (unpublished results). However, this approach may have been limited, and more advanced models like recurrent neural networks, transformers^21^, or DeepHit^22^, could be explored in the future. Moreover, incorporating more diverse models within the superlearner framework may improve its overall performance. Third, we observed a deterioration in performance on temporal evaluation, reflecting potential shifts in patient care practices, catheter management strategies, patient populations or data recording procedures in the EHR system. Calibration deterioration is more common than discrimination deterioration and can often be corrected by model recalibration^23,24^. When we refitted the models on data per calendar year separately, we observed a decline in discrimination with each successive year. This suggests that the CLABSI prediction task is becoming increasingly challenging, raising concerns about the utility of these models for data beyond 2020 if this trend continues in the coming years. Lastly, we acknowledge limitations related to fairness in our study. Although we included age and gender as predictors in our models, we did not extract data on race, ethnicity or socioeconomic status. This limits our ability to fully explore how the predictive performance varies across different demographic groups. Furthermore, wealthier patients who can afford single rooms may be at a lower risk of hospital-acquired infections, including CLABSI, compared to patients in shared rooms. However, it is difficult to determine whether socioeconomic bias is reflected in our models.

Further research is needed to determine if CLABSI models have practical applicability or are generalizable to other hospitals. Given the uncertain intervention effectiveness, the practical applicability of these models remains unclear. Following the CLABSI definition, our models predict infections in patients with central catheters where no other identifiable source of infection can be established; these infections are not always directly attributed to the central catheter. Consequently, interventions like improved line care do not guarantee prevention of infections classified as CLABSI, especially in a hospital where the infection prevention strategy is already of high standard. Implementation studies assessing the impact on clinical decision-making, patient outcomes, user-perceived usefulness or even adverse outcomes (e.g.: unnecessary catheter removals) could help determine if the model adds practical value in healthcare settings. The data extraction from various clinical databases and the proprietary format of catheter observations would hinder generalization to hospitals using a different EHR system. To ensure model compatibility across different EHR systems, it is essential to standardize data extraction for both model development and implementation, and perform clinical concepts mapping, using standards like FHIR^25^ and OMOP CDM^26^. Furthermore, we adhered to the Belgian Institute of Public Health’s definition of CLABSI, which might differ slightly from other local or international definitions. However, testing the models’ performance in Belgian hospitals with the same EHR system may prove feasible, after adapting the mapping configuration between extracted items and the clinical concepts (without changing the extraction format) and assessing the quality of the data extraction and preparation.

## Conclusion

Our study proposed a number of dynamic models for predicting the 7-day risk of CLABSI in patients with central catheters. The overall performance of the models was modest: the maximum AUROC of 0.751 was achieved by the superlearner model at day 6. Temporal evaluation revealed dataset shifts. The models showed clinical utility for generating alerts to check standard care interventions based on medium-risk thresholds, but not for alerts to consider technological interventions based on high-risk thresholds.

## Supporting information

Supplement - TRIPOD+AI checklist

## Data Availability

The data underlying this article cannot be shared publicly due to privacy of individuals that participated in the study. Data are located in controlled access database at UZ Leuven.

## Data and code availability

The code to build the RF and XGB models has been run on a high-performance computing cluster using 36 cores and 128 GB RAM for the RF model and an NVIDIA V100 GPU for the XGB model. R version 4.2.1, randomForestSRC^27^ 3.2.2, ranger^28^ 0.15.1 and xgboost^29^ 2.0.0.1 have been used to build the models.

## Acknowledgements

The study adhered to the principles of the Declaration of Helsinki (current version), the principles of Good Clinical Practice (GCP), and all relevant regulatory requirements. Ethical review was sought from the Ethics Committee Research UZ / KU Leuven, Belgium, which is the local ethics committee at UZ Leuven (https://admin.kuleuven.be/raden/en/ethics-committee-research-uz-kuleuven#). The collection, processing and disclosure of personal data, such as patient health and medical information were in compliance with applicable personal data protection and the processing of personal data (Directive 95/46/EC and Belgian law of December 8, 1992 on the Protection of the Privacy in relation to the Processing of Personal Data). Patient stay identifiers were coded using the pseudo-identifier available in the data warehouse of the participating hospital.

The resources and services used in this work were provided by the VSC (Flemish Supercomputer Center), funded by the Research Foundation - Flanders (FWO) and the Flemish Government.

## Authors’ contribution

ALBU Elena: Conceptualization, Data curation, Formal analysis, Investigation, Methodology, Software, Visualization, Writing - original draft. GAO Shan: Conceptualization, Data curation, Formal analysis, Investigation, Methodology, Software, Visualization, Writing - original draft. Stijnen Pieter: Conceptualization, Data curation, Funding acquisition, Writing - review & editing. RADEMAKERS Frank: Supervision, Writing - review & editing. COSSEY Veerle: Supervision, Writing - review & editing. DEBAVEYE Yves: Supervision, Writing - review & editing. JANSSENS Christel: Supervision, Writing - review & editing. VAN CALSTER Ben: Conceptualization, Funding acquisition, Methodology, Project administration, Supervision, Writing - review & editing. WYNANTS Laure: Conceptualization, Funding acquisition, Methodology, Project administration, Supervision, Writing - review & editing

## Abbreviations

AUPRC: Area Under the Precision Recall Curve
AUROC: Area Under the Receiver Operating Characteristic curve
BS: Brier Score
BSI: Bloodstream Infection
BSS: Brier Skill Score
CICC: Centrally Inserted Central Catheters (CICC)
CIF: Cumulative Incidence Functions
CLABSI: Central Line-Associated Bloodstream Infections
CR: Competing Risks
CRP: C-reactive protein
CVC: Central Venous Catheter
ECI: Estimated Calibration Index
EHR: Electronic Health Records
FWO: Research Foundation - Flanders
ICD: International Classification of Diseases
ICU: Intensive Care Unit
IQR: Interquartile Range
KWS: Klinisch Werkstation (EHR System)
LC-BSI: Laboratory-Confirmed Bloodstream Infection
LM: Landmark
LR: logrank
LRCR: logrankCR (logrank competing risks)
LWS: Laboratorium Werkstation (Laboratory System)
ML: Machine Learning
NMSE: Normalized Mean Square Error
OOB: Out of bag
PICC: Peripherally Inserted Central Catheter
PDMS: Patient Data Management System (ICU Management System)
RAM: Random Access Memory
RF: Random Forest
tc-CICC: tunnelled cuffed Centrally Inserted Central Catheters (CICC)
TIVAD: Totally Implanted Vascular Access Devices
tnc-CICC: tunnelled non-cuffed Centrally Inserted Central Catheters (CICC)
TPN: Total Parenteral nutrition
VSC: Flemish Supercomputer Center
WBC: White Blood Cells count

