## Supplementary materials for "Hospital-wide, dynamic, individualized prediction of central line-associated bloodstream infections - development and temporal evaluation of six prediction models"

### Supplementary material 1 - Example of landmark dataset for dynamic model building

An example of the landmark dataset is presented in Table 3. Admission identifiers are anonymized and noise has been added to the variables.

| admission_id | CAT_catheter_episode | LM | CAT_consecutive_current_days_CVC | CAT_catheter_location_binary_all_Neck | MED_L5_7d_B05BA10_combinations | CARE_VS_temperature_max | LAB_WBC_count_last | type | eventtime |
| --- | --- | --- | --- | --- | --- | --- | --- | --- | --- |
| 1 | 1 | 0 | 1 | 1 | 0 | 37.5 | 8.9 | Discharge | 4.7 |
| 1 | 1 | 1 | 2 | 1 | 0 | 36.9 | 8.5 | Discharge | 4.7 |
| 1 | 1 | 2 | 3 | 1 | 0 | 36.5 | 8.4 | Discharge | 4.7 |
| 1 | 1 | 3 | 4 | 1 | 0 | 36.4 | 8.9 | Discharge | 4.7 |
| 1 | 1 | 4 | 5 | 1 | 0 | 36.5 | 7.7 | Discharge | 4.7 |
| 2 | 1 | 0 | 0 | 0 | 0 | 39.1 | 9.8 | Discharge | 2.3 |
| 2 | 1 | 1 | 0 | 0 | 0 | 38.5 | 12.3 | Discharge | 2.3 |
| 2 | 1 | 2 | 0 | 0 | 0 | 38.3 | 8.7 | Discharge | 2.3 |
| 2 | 2 | 0 | 0 | 0 | 0 | 36.9 | 7.2 | Discharge | 5.3 |
| 2 | 2 | 1 | 0 | 0 | 0 | 37.7 | 10.9 | Discharge | 5.3 |
| 2 | 2 | 2 | 0 | 0 | 0 | 37.5 | 11.4 | Discharge | 5.3 |
| 2 | 2 | 3 | 0 | 0 | 0 | 37.1 | 12.5 | Discharge | 5.3 |
| 2 | 2 | 4 | 0 | 0 | 0 | 37.3 | 12.2 | Discharge | 5.3 |
| 2 | 2 | 5 | 0 | 0 | 0 | 37.5 | 6.3 | Discharge | 5.3 |
| 3 | 1 | 0 | 1 | 0 | 0 | 37.5 | 16.6 | Discharge | 3.5 |
| 3 | 1 | 1 | 2 | 0 | 0 | 37.8 | 11.2 | Discharge | 3.5 |
| 3 | 1 | 2 | 3 | 0 | 0 | 37.6 | 17.6 | Discharge | 3.5 |
| 3 | 1 | 3 | 4 | 0 | 0 | 37.5 | 10.6 | Discharge | 3.5 |
| 3 | 2 | 0 | 1 | 1 | 1 | 37.3 | 7.9 | CLABSI | 5.5 |
| 3 | 2 | 1 | 2 | 1 | 1 | 37.5 | 9.3 | CLABSI | 5.5 |
| 3 | 2 | 2 | 3 | 1 | 1 | 37.6 | 14.3 | CLABSI | 5.5 |
| 3 | 2 | 3 | 4 | 1 | 1 | 37.6 | 11.8 | CLABSI | 5.5 |
| 3 | 2 | 4 | 5 | 1 | 1 | 37.5 | 11.2 | CLABSI | 5.5 |
| 3 | 2 | 5 | 6 | 1 | 1 | 37.4 | 11.5 | CLABSI | 5.5 |

Table 3: Example of landmark data structure including selected features

### Supplementary material 2 - Variable descriptions

Baseline variables are invariant for a catheter episode and are known at the start of the catheter episode. Time-varying variables represent variables that can vary from one landmark to another.

**Continuous variables**: Whenever multiple measurements are available during a time window (typically 24 hours) for continuous variables these are aggregated into a single landmark value. The aggregation rule (e.g.: maximum, minimum) with the most clinical significance is chosen (e.g.: maximum temperature in last 24 hours). Whenever no measurements are taken in the time window, the variable value is represented as a missing value.

**Binary variables** are coded for presence or absence of specific clinical events that are recorded in the EHR only when present, e.g.: antibiotics administration.

**Categorical variables** (e.g.: catheter type) that might occur simultaneously in the aggregation window are coded as binary variables (0/1) for all values recorded (e.g.: if a patient has two catheters: CVC and port catheter, two binary variables are kept for CVC and port catheter). Whenever categorical variables (with two or more categories) are expected to be recorded regardless of the category (e.g.: admission source), and no value is present, the variable value is represented as a missing value.

Values outside the possible range have been deleted before variable aggregation, for the following variables and ranges (min – max): temperature, [30, 45]; systolic and diastolic blood pressure, [30, 370]; respiratory rate, [0, 900]; heart rate, [0, 500]; oxygen saturation, [0, 100]; CVP (central venous pressure), [-5, 20]; weight, [0.05, 250]; length, [0.05, 250]; glycemia, [0, 2000]. These deletions might result in missing values to be imputed later.

Table 4 presents the variables description, as well as their inclusion in the developed models. The variables in the “complete set” are used for missing data imputation and for the development of one RF and one XGB model; The variables in the “limited set” are used for the development in the landmark cause-specific supermodel.

| Variable | Description | Type | Baseline / time-varying | Complete set | Limited set |
| --- | --- | --- | --- | --- | --- |
| functioneelDossierNr | Admission ID. A 'functional' admission can contain more 'administrative' admissions (e.g.: a patient that has an emergency admission and then is hospitalised in the cardiology medical unit will have only one functional admission, because they are logically linked together. There might still be administrative admissions not linked together for billing/insurance reasons, e.g. for foreign patients, but these should be extremely rare, if any. | NA | NA | NO | NO |
| CAT_catheter_episode | The ID of the catheter epsiode. First catheter episode is 1, second is 2, and so on… A catheter episode groups together overlapping catheters and catheters separated apart by less than 48 hours (between the end date of a catheter and start date of any another catheter). | cont | BASE | YES | NO |
| LM | Landmark ID. LM 0 constitutes the baseline, which will contain only information at the exact time of (first) catheter access/observation or information from the past 24h before the time of (first) catheter access/observation | cont | TV | YES | YES |
| eventtime | Time of event: CLABSI / Death / Discharge (or catheter removal / contact with palliative care) | cont | NA | NO | NO |
| type | Type of event: CLABSI / Death / Discharge (where Discharge means discharge, contact with palliative care or catheter removal) | categ | NA | NO | NO |
| ADM_admission_to_catheter | Time since admission to catheter epsiode start (in days). | cont | BASE | YES | YES |
| ADM_admission_type_binary_all_Emergency | Admission reason (emergency or other) | binary | BASE | YES | NO |
| ADM_nr_adm_past_180_days | Number of admissions in the past 180 days for the patient | cont | BASE | YES | YES |
| ADM_nr_emergency_adm_past_180_days | Number of emergency admissions in the past 180 days for the patient | cont | BASE | YES | NO |
| ADM_unplanned_readmission | Is the admission unplanned? | binary | BASE | YES | NO |
| CARE_ICU_ECMO | Has the patient been planned to be on ECMO in the timeframe since previous LM? (only recorded in ICU, considered 0 when not recorded) | binary | TV | YES | NO |
| CARE_ISO_binary_all_protective_isolation | Has the patient been in protective isolation since previous LM? | binary | TV | YES | NO |
| CARE_ISO_binary_all_source_isolation | Has the patient been in source isolation since previous LM? | binary | TV | YES | YES |
| CARE_NEU_GCS_score_last | GCS (Glasgow Coma Scale) total score. | cont | TV | YES | YES |
| CARE_PHY_drain | Was a drain monitored for the patient since last LM? | binary | TV | YES | YES |
| CARE_SAF_freedom_restriction_categorical_last | The last freedom restriction measure recorded since last LM (partial/complete). Partial = Dag en nacht; Complete = Tijdens dag, Tijdens avond, Tijdens ochtend, Tijdens ochtend, Tijdens namiddag or Tijdens nacht (missing whenever not recorded during 2 LMs) | categ | TV | YES | NO |
| CARE_SAF_mobility_assistance_binary_all_no_help | Did the patient move independently without mobility assistance (for either transfer, move or bed installation) since previous LM? | binary | TV | YES | NO |
| CARE_SYM_pruritus_binary_all_YES | Pruritus evaluation since last LM (missing when not recorded). | binary | TV | YES | NO |
| CARE_SYM_RASS_max | Maximum value of RASS (Richmond Agitation-Sedation Scale) since last LM (missing when not recorded) | cont | TV | YES | NO |
| CARE_VS_breathing_aid | Did the patient receive breathing aid since last LM? | binary | TV | YES | NO |
| CARE_VS_CVP_last | Last value of central venous pressure since previous landmark. For baseline (LM 0) the last value from the previous 24 hours is used. | cont | TV | YES | NO |
| CARE_VS_CVP_measured | Has central venous pressure been measured during the time interval between current LM and previous LM. CVP is meassued using the central line, therefore it is a catheter manipulation that potentially be associated with higher risk of CLABSI. (This is the opposite of a missing data indicator). | binary | TV | YES | NO |
| CARE_VS_diastolic_BP_last | Last value of diastolic blood pressure since previous landmark. For baseline (LM 0) the last value from the previous 24 hours is used. | cont | TV | YES | NO |
| CARE_VS_heart_rate_max | Maximum value of heart rate since previous landmark. For baseline (LM 0) the last value from the previous 24 hours is used. | cont | TV | YES | YES |
| CARE_VS_MV | Is the patient on mechanical ventilation (MV) since previous landmark? A patient is considered on MV if at least one value of PEEP or FiO2 are recorded between 2 landmarks. Only valid for ICU patients | binary | TV | YES | NO |
| CARE_VS_oxygen_saturation_last | Last Oxygen saturation value (%) since previous LM. | cont | TV | YES | NO |
| CARE_VS_respiratory_rate_last | Last value of respiratory rate since previous landmark. For baseline (LM 0) the last value from the previous 24 hours is used. | cont | TV | YES | YES |
| CARE_VS_systolic_BP_last | Last value of systolic blood pressure since previous landmark. For baseline (LM 0) the last value from the previous 24 hours is used. | cont | TV | YES | NO |
| CARE_VS_temperature_max | Maximum value of temperature since previous landmark. For baseline (LM 0) the last value from the previous 24 hours is used. Only temperatures in the range (30 °C, 45 °C) are kept, the others are deleted. Maximum value is used to correct for very low temperatures measured by devices in ICU, when the temperature falls closer to the room temperature. | cont | TV | YES | YES |
| CARE_WND_wound_type_binary_all_closed_wound | Is there a closed wound present since previous LM? | binary | TV | YES | NO |
| CARE_WND_wound_type_binary_all_open_wound | Is there an open wound present since previous LM? | binary | TV | YES | NO |
| CARE_WND_wound_type_binary_all_post_suture | Is there a post suture wound present since previous LM? | binary | TV | YES | NO |
| CARE_WND_wound_type_binary_all_suture | Is there a suture wound present since previous LM? | binary | TV | YES | NO |
| CAT_bandage_change | Indicator if at least one bandage change (for any central line) has happened since previous LM. The value will be 1 for LM 0, as the bandage has been placed. | binary | TV | YES | NO |
| CAT_bandage_observation_binary_all_Bloody_or_Moist | Has there been a bandage (or insert point) observation of type Bloody or Moist since previous LM? _binary_all means that all values encountered in the specified timeframe are binarized (not on last value) | binary | TV | YES | NO |
| CAT_bandage_observation_binary_all_Normal | Has there been a bandage (or insert point) observation of type Normal since previous LM? _binary_all means that all values encountered in the specified timeframe are binarized (not on last value) | binary | TV | YES | NO |
| CAT_bandage_observation_binary_all_Other_Hema_Pus_Loose_Necro | Has there been a bandage (or insert point) observation of other type (hematome, pus, loose, necrosis) since previous LM? _binary_all means that all values encountered in the specified timeframe are binarized (not on last value) | binary | TV | YES | NO |
| CAT_bandage_observation_binary_all_Red | Has there been a bandage (or insert point) observation of type Red since previous LM? _binary_all means that all values encountered in the specified timeframe are binarized (not on last value) | binary | TV | YES | NO |
| CAT_bandage_type_binary_last_gauze | Bandage of type Gauze (gaasverband) or Silicone registerd since previous LM. Silicone is like gauze wrt stickiness (needs more changes, closer risk) and like polyutheane wrt transparency. _binary_last means that only the last value encountered in the specified timeframe is taken. | binary | TV | YES | NO |
| CAT_catheter_location_binary_all_Arm | Was there a catheter connected at location Arm since previous LM? | binary | TV | YES | NO |
| CAT_catheter_location_binary_all_Collarbone | Was there a catheter connected at location Collarbone since previous LM? | binary | TV | YES | NO |
| CAT_catheter_location_binary_all_Groin | Was there a catheter connected at location Groin since previous LM? | binary | TV | YES | NO |
| CAT_catheter_location_binary_all_Navel | Was there a catheter connected at location Navel since previous LM? | binary | TV | YES | NO |
| CAT_catheter_location_binary_all_Neck | Was there a catheter connected at location Neck since previous LM? | binary | TV | YES | NO |
| CAT_catheter_location_binary_all_Other | Was there a catheter connected at location other than Neck, Collarbone, Navel, Arm, Groin since previous LM? | binary | TV | YES | NO |
| CAT_catheter_placement_binary_all_bedside | Indicator if any catheter during the catheter episode (and before current LM) has been placed in VE (at bed side). Whenever not recorded it is considered 0 (no missing values) | binary | TV | YES | NO |
| CAT_catheter_placement_binary_all_OR | Indicator if any catheter during the catheter episode (and before current LM) has been placed in OR. Whenever not recorded it is considered 0 (no missing values). | binary | TV | YES | NO |
| CAT_consecutive_current_days_CVC | Total number of days a CVC was present in current CVC episode. If a CVC is connected and disconnected, this will be reflected. E.g.: If CVC days are 0 1 1 1 0 0 1 1, the number of days will be 0 1 2 3 0 0 1 2 | cont | TV | YES | YES |
| CAT_consecutive_current_days_Dialysis_CVC | Total number of days a Dialysis CVC was present in current Dialysis_CVC episode. If a Dialysis_CVC is connected and disconnected, this will be reflected. E.g.: If Dialysis_CVC days are 0 1 1 1 0 0 1 1, the number of days will be 0 1 2 3 0 0 1 2 | cont | TV | YES | YES |
| CAT_consecutive_current_days_PICC | Total number of days a PICC was present in current PICC episode. If a PICC is connected and disconnected, this will be reflected. E.g.: If PICC days are 0 1 1 1 0 0 1 1, the number of days will be 0 1 2 3 0 0 1 2 | cont | TV | YES | YES |
| CAT_consecutive_current_days_Port_a_cath | Total number of days a Port_a_cath was present in current Port_a_cath episode. If a Port_a_cath is connected and disconnected, this will be reflected. E.g.: If Port_a_cath days are 0 1 1 1 0 0 1 1, the number of days will be 0 1 2 3 0 0 1 2 | cont | TV | YES | YES |
| CAT_consecutive_current_days_Tunneled_CVC | Total number of days a Tunneled CVC was present in current Tunneled CVC episode. If a CVC is connected and disconnected, this will be reflected. E.g.: If Tunneled CVC days are 0 1 1 1 0 0 1 1, the number of days will be 0 1 2 3 0 0 1 2 | cont | TV | YES | YES |
| CAT_days_since_last_bandage_changed | Number of days since last bandage change was recorded (for any central line). It is considered that at the time of first catheter placement (start of catheter episode) the bandage is placed (equivalent to changed) so the maximum value can be number of days since placement. This variable encodes missingness. | cont | TV | YES | NO |
| CAT_days_since_last_bandage_obs | Number of days since last catheter observation was recorded. It is considered that at the time of first catheter access/observation (start of catheter episode) the bandage is observed, so the maximum value can be number of days since placement. This variable encodes missingness. | cont | TV | YES | YES |
| CAT_days_since_last_tube_change | Number of days since last tube change was recorded. It is considered that at the time of first catheter access/observation (start of catheter episode) the tube is placed (equivalent to changed), so the maximum value can be number of days since placement. This variable encodes missingness. | cont | TV | YES | YES |
| CAT_lumens_flushed | Distinct number of lumens flushed at least once since previous LM. | cont | TV | YES | YES |
| CAT_needle_length_max | Needle length (typically 16, 19, 25, …). In case of more catheters the maximum needle length is taken (if for example a catheter has a needle length of 19 and another one has needle length unknown/missing, the value of 19 will be used. This is different than the lumens calculation.) Not applicable to dialysis catheters. | cont | TV | YES | YES |
| CAT_nr_bandage_obersations | Number of catheter observations recorded since previous LM. 0 will encode missingness. | cont | TV | YES | YES |
| CAT_number_central_lines | Number of central lines connected since previous LM | cont | TV | YES | NO |
| CAT_number_peripheral_catheters | Number of peripheral catheters connected since previous LM. | cont | TV | YES | NO |
| CAT_tube_change | Indicator that a tube change took placesince previous LM. 1 = tube change, 0 = no tube change registered in the system. On LM 0 it is always 1 (tube placed is considered tube change). Not applicable to dialysis catheters. | binary | TV | YES | NO |
| CLABSI_history | Did the patient experience a CLABSI event in the past 3 months since LM time? | binary | TV | YES | NO |
| COM_arterial_occlusive_disease_before_LM | Has arterial occlusive disease been registered as a comorbidity before current LM time? | binary | TV | YES | NO |
| COM_COPD_before_LM | Has COPD been registered as a comorbidity before current LM time? | binary | TV | YES | NO |
| COM_heart_failure_before_LM | Has heart failure been registered as a comorbidity before current LM time? | binary | TV | YES | NO |
| COM_HIV_before_LM | Has HIV been registered as a comorbidity before current LM time? | binary | TV | YES | NO |
| COM_lymphoma_before_LM | Has lymphoma been registered as a comorbidity before current LM time? | binary | TV | YES | NO |
| COM_ORG_heart_and_circulatory_system_before_LM | Has a comrbidity for organ heart and circulatory system been registered before current LM time? | binary | TV | YES | NO |
| GEN_LM_month_categ | Month of the landmark time point, categorized as months (January - December) | categ | TV | YES | NO |
| HC_101_Coronary_atherosclerosis_and_other_heart_disease | Whether the patient had any coronary atherosclerosis and other heart disease (ccs) at admission | binary | BASE | YES | NO |
| HC_106_Cardiac_dysrhythmias | Whether the patient had cardiac dysrhythmias (ccs) at admission | binary | BASE | YES | NO |
| HC_117_Other_circulatory_disease | Whether the patient had any other circulatory disease (ccs) at admission | binary | BASE | YES | NO |
| HC_155_Other_gastrointestinal_disorders | Whether the patient had any other gastrointestinal disorders (ccs) at admission | binary | BASE | YES | NO |
| HC_158_Chronic_kidney_disease | Whether the patient had any chronic kidney disease (ccs) at admission | binary | BASE | YES | NO |
| HC_238_Complications_of_surgical_procedures_or_medical_care | Whether the patient had any complications of surgical procedures or medical care (ccs) at admission | binary | BASE | YES | NO |
| HC_253_Allergic_reactions | Whether the patient had any allergic reactions (ccs) at admission | binary | BASE | YES | NO |
| HC_2617_Adverse_effects_of_medical_drugs | Whether the patient had any adverse effects of medical drugs (ccs) at admission | binary | BASE | YES | NO |
| HC_3_Bacterial_infection_unspecified_site | Whether the patient had any bacterial infection at unspecified site (ccs) at admission | binary | BASE | YES | NO |
| HC_42_Secondary_malignancies | Whether the patient any secondary malignancies (ccs) at admission | binary | BASE | YES | NO |
| HC_45_Maintenance_chemotherapy_radiotherapy | Whether the patient had any maintenance chemotherapy radiotherapy (ccs) at admission | binary | BASE | YES | NO |
| HC_53_Disorders_of_lipid_metabolism | Whether the patient had disorders of lipid metabolism (ccs) at admission | binary | BASE | YES | NO |
| HC_55_Fluid_and_electrolyte_disorders | Whether the patient had suffered from fluid and electrolyte disorders (ccs) at admission | binary | BASE | YES | NO |
| HC_58_Other_nutritional_endocrine_and_metabolic_disorders | Whether the patient had any other nutritional endocrine and metabolic disorders (ccs) at admission | binary | BASE | YES | NO |
| HC_59_Deficiency_and_other_anemia | Whether the patient had any deficiency and other anemia (ccs) at admission | binary | BASE | YES | NO |
| HC_96_Heart_valve_disorders | Whether the patient had any heart valve disorders (ccs) at admission | binary | BASE | YES | NO |
| HC_98_Essential_hypertension | Whether the patient had suffered from essential hypertension (ccs) at admission | binary | BASE | YES | NO |
| LAB_ALT_last | ALT, last value since previous LM. Unit: U/L | cont | TV | YES | NO |
| LAB_APTT_last | APTT (activated partial thromboplastin time), last value since previous LM. Unit: s | cont | TV | YES | NO |
| LAB_aspergillus_ag_last | Aspergillus antigen, last value since previous LM. Unit: none | cont | TV | YES | YES |
| LAB_AST_last | AST, last value since previous LM. Unit: U/L | cont | TV | YES | NO |
| LAB_bilirubin_last | Bilirubin, last value since previous LM. Unit: mg/dL | cont | TV | YES | YES |
| LAB_CK_last | Creatine kinase (CK), last value since previous LM. Unit: U/L | cont | TV | YES | YES |
| LAB_creatinine_clearance_last | Creatinine clearance, last value since previous LM. Unit: U/L | cont | TV | YES | YES |
| LAB_creatinine_last | Creatinine, last value since previous LM. Unit: mg/dL | cont | TV | YES | YES |
| LAB_CRP_last | CRP, last value since previous LM. Unit: mg/L | cont | TV | YES | YES |
| LAB_D_dimer_last | D-dimer, last value since previous LM. Unit: µg/L | cont | TV | YES | YES |
| LAB_ferritin_last | Ferritin, last value since previous LM. Unit: µg/L | cont | TV | YES | YES |
| LAB_fibrinogen_last | Fibrinogen, last value since previous LM. Unit: g/L | cont | TV | YES | NO |
| LAB_glucose_arterial_last | Arterial glucose, last value since previous LM. Unit: mg/dL | cont | TV | YES | YES |
| LAB_glucose_last | Glucose, last value since previous LM. Unit: mg/dL | cont | TV | YES | YES |
| LAB_haematocrit_last | Haematocrit, last value since previous LM. Unit: none | cont | TV | YES | NO |
| LAB_Hemoglobine_last | Hemoglobine, last value since previous LM. Unit: g/dL | cont | TV | YES | NO |
| LAB_is_neutropenia | Is neutropenia detected since previous LM? Neutropenia is defined as in Sciensano 2019: absolute value of neutrophils or WBC count <500/mm3. | binary | TV | YES | NO |
| LAB_LDH_last | LDH, last value since previous LM. Unit: U/L | cont | TV | YES | YES |
| LAB_natrium_last | Natirum, last value since previous LM. Unit: mmol/L | cont | TV | YES | YES |
| LAB_NT_proBNP_last | NT-proBNP, last value since previous LM. Unit: ng/L | cont | TV | YES | NO |
| LAB_O2_saturation_last | O2 saturation, last value since previous LM. Unit: % | cont | TV | YES | YES |
| LAB_pH_last | pH, last value since previous LM. Unit: none | cont | TV | YES | YES |
| LAB_Platelet_count_last | Platelet count (Bloedplaatjes telling), last value since previous LM. Unit: 10**9/L | cont | TV | YES | YES |
| LAB_pO2_last | pO2, last value since previous LM. Unit: mm Hg | cont | TV | YES | YES |
| LAB_potassium_last | Potassium, last value since previous LM. Unit: mmol/L | cont | TV | YES | NO |
| LAB_PT_INR_last | Prothrombin time, last value since previous LM. Unit: INR (International normalized ratio) | cont | TV | YES | NO |
| LAB_PT_percent_last | Prothrombin time, last value since previous LM. Unit: percent | cont | TV | YES | NO |
| LAB_PT_sec_last | Prothrombin time, last value since previous LM. Unit: seconds | cont | TV | YES | NO |
| LAB_RBC_count_last | Red blood cell count, last value since previous LM. Unit: 10**12/L | cont | TV | YES | YES |
| LAB_SPE_albumin_alpha_1_globulin_last | Albumin alpha 1 globulin, last value since previous LM. Unit: g/L (SPE = Serum protein electrophoresis) | cont | TV | YES | YES |
| LAB_troponine_T_last | Troponine T, last value since previous LM. Unit: µg/L | cont | TV | YES | NO |
| LAB_TSH_last | TSH (thyroid stimulating hormone), last value since previous LM. Unit: mIU/L | cont | TV | YES | YES |
| LAB_urea_last | Urea, last value since previous LM. Unit: mg/dL | cont | TV | YES | YES |
| LAB_vancomycine_last | Vancomycine, last value since previous LM. Unit: mg/L | cont | TV | YES | YES |
| LAB_WBC_count_last | WBC count, last value since previous LM. Unit: 10**9/L | cont | TV | YES | YES |
| LAB_WBC_Monocytes_last | Monocytes amount, last value since previous LM. Unit: 10**9/L | cont | TV | YES | NO |
| LAB_WBC_Neutrophils_last | Neutrophils amount, last value since previous LM. Unit: 10**9/L | cont | TV | YES | YES |
| MB_infection_time_window_binary_all_blood | Has there been a positive culture, for blood sample type, in the last 17 days (time window used for secondary BSIs). The validation time of the sample is used (as opposed to the CLABSI calculation, where the date foreseen for the sample collection is used) | binary | TV | YES | NO |
| MB_infection_time_window_binary_all_catheter | Has there been a positive culture, for catheter sample type, in the last 17 days (time window used for secondary BSIs). The validation time of the sample is used (as opposed to the CLABSI calculation, where the date foreseen for the sample collection is used) | binary | TV | YES | NO |
| MB_infection_time_window_binary_all_deep_tissue | Has there been a positive culture, for deep tissue sample type, in the last 17 days (time window used for secondary BSIs). The validation time of the sample is used (as opposed to the CLABSI calculation, where the date foreseen for the sample collection is used) | binary | TV | YES | NO |
| MB_infection_time_window_binary_all_drain | Has there been a positive culture, for drain sample type, in the last 17 days (time window used for secondary BSIs). The validation time of the sample is used (as opposed to the CLABSI calculation, where the date foreseen for the sample collection is used) | binary | TV | YES | NO |
| MB_infection_time_window_binary_all_GI | Has there been a positive culture, for GI (gastrointestinal) sample type, in the last 17 days (time window used for secondary BSIs). The validation time of the sample is used (as opposed to the CLABSI calculation, where the date foreseen for the sample collection is used) | binary | TV | YES | NO |
| MB_infection_time_window_binary_all_lung | Has there been a positive culture, for lung sample type, in the last 17 days (time window used for secondary BSIs). The validation time of the sample is used (as opposed to the CLABSI calculation, where the date foreseen for the sample collection is used) | binary | TV | YES | NO |
| MB_infection_time_window_binary_all_skin | Has there been a positive culture, for skin sample type, in the last 17 days (time window used for secondary BSIs). The validation time of the sample is used (as opposed to the CLABSI calculation, where the date foreseen for the sample collection is used) | binary | TV | YES | NO |
| MB_infection_time_window_binary_all_sputum | Has there been a positive culture, for sputum sample type, in the last 17 days (time window used for secondary BSIs). The validation time of the sample is used (as opposed to the CLABSI calculation, where the date foreseen for the sample collection is used) | binary | TV | YES | NO |
| MB_infection_time_window_binary_all_urogen | Has there been a positive culture, for urogen sample type, in the last 17 days (time window used for secondary BSIs). The validation time of the sample is used (as opposed to the CLABSI calculation, where the date foreseen for the sample collection is used) | binary | TV | YES | NO |
| MB_number_contaminants_in_blood | The number of samples with a germ of type contaminant during the time interval between current LM and previous LM. The validation time of the sample is used (as opposed to the CLABSI calculation, where the date foreseen for the sample collection is used) | cont | TV | YES | NO |
| MB_other_infection_than_BSI_during_window | Has there been a positive culture, of any other type than blood, in the last 17 days (time window used for secondary BSIs). The validation time of the sample is used (as opposed to the CLABSI calculation, where the date foreseen for the sample collection is used) | binary | TV | YES | YES |
| MED_7d_CITRA_LOCK | Has product CITRA-LOCK been ordered for the patient in the previous 7 days from LM time | binary | TV | YES | NO |
| MED_7d_immunoglobulins | Have immunoglobulins (J06, L01F, L04AA03, L04AA04, V09HA01, J06BB02, J07AM52, V09IX03) been ordered for the patient in the previous 7 days from LM time | binary | TV | YES | NO |
| MED_7d_number_of_IV_med | Number of distinct medication (distinct ATC codes) ordered for intravenous administration (IV administration route) in the previous 7 days from LM time. | cont | TV | YES | YES |
| MED_7d_number_of_ORAL_med | Number of distinct medication (distinct ATC codes) ordered for oral administration (P.O. administration route) in the previous 7 days from LM time. | cont | TV | YES | YES |
| MED_7d_VANCO_CEFTA_LOCK | Has product VANCOLOCK (MAGISTRALE BEREIDING) or CEFTAZIDIM LOCK been ordered for the patient in the previous 7 days from LM time | binary | TV | YES | NO |
| MED_L2_7d_H02_CORTICOSTEROIDS_FOR_SYSTEMIC_USE | Have any drugs in ATC group (level 2) H02 (CORTICOSTEROIDS) been ordered for the patient in the previous 3 days from LM time | binary | TV | YES | NO |
| MED_L2_7d_J01_ANTIBACTERIALS_FOR_SYSTEMIC_USE | Have any drugs in ATC group (level 2) J01 (ANTIBACTERIALS FOR SYSTEMIC USE) been ordered for the patient in the previous 7 days from LM time | binary | TV | YES | NO |
| MED_L2_7d_J02_ANTIMYCOTICS_FOR_SYSTEMIC_USE | Have any drugs in ATC group (level 2) J02 (ANTIMYCOTICS FOR SYSTEMIC USE) been ordered for the patient in the previous 7 days from LM time | binary | TV | YES | YES |
| MED_L2_7d_J04_ANTIMYCOBACTERIALS | Have any drugs in ATC group (level 2) J04 (ANTIMYCOBACTERIALS) been ordered for the patient in the previous 7 days from LM time | binary | TV | YES | NO |
| MED_L2_7d_J05_ANTIVIRALS_FOR_SYSTEMIC_USE | Have any drugs in ATC group (level 2) J05 (ANTIVIRALS FOR SYSTEMIC USE) been ordered for the patient in the previous 3 days from LM time | binary | TV | YES | NO |
| MED_L2_7d_J06_IMMUNE_SERA_AND_IMMUNOGLOBULINS | Have any drugs in ATC group (level 2) J06 (IMMUNE SERA AND IMMUNOGLOBULINS) been ordered for the patient in the previous 7 days from LM time | binary | TV | YES | NO |
| MED_L2_7d_L01_ANTINEOPLASTIC_AGENTS | Have any drugs in ATC group (level 2) L01 (ANTINEOPLASTIC AGENTS) been ordered for the patient in the previous 7 days from LM time | binary | TV | YES | YES |
| MED_L2_7d_L04_IMMUNOSUPPRESSANTS | Have any drugs in ATC group (level 2) L04 (IMMUNOSUPPRESSANTS) been ordered for the patient in the previous 7 days from LM time | binary | TV | YES | NO |
| MED_L5_7d_B05BA01_amino_acids | Have amino acids (TPN) been ordered for the patient in the previous 7 days from LM time | binary | TV | YES | NO |
| MED_L5_7d_B05BA02_fat_emulsions | Have fat emulsions (TPN) been ordered for the patient in the previous 7 days from LM time | binary | TV | YES | NO |
| MED_L5_7d_B05BA03_carbohydrates | Have carbohydrates (TPN) been ordered for the patient in the previous 7 days from LM time | binary | TV | YES | NO |
| MED_L5_7d_B05BA10_combinations | Have TPN combinations (TPN) been ordered for the patient in the previous 7 days from LM time | binary | TV | YES | YES |
| MS_alternative_flag | Is the patient located in a nursing ward which is not common to his/her own medical speciality at landmark time | binary | TV | YES | NO |
| MS_ICU_time_before_catheter | Time spent in ICU since admission to catheter placement. | cont | BASE | YES | YES |
| MS_is_24h_net_OR_unit | Is the patient in net OR within the recent 24h (from last LM to the current LM)? | binary | TV | YES | NO |
| MS_is_ICU_unit | Is the patient now (at the exact second of the current LM) in ICU? | binary | TV | YES | NO |
| MS_medical_specialty | The medical specialty that the patient's doctor registered at each landmark time point | categ | TV | YES | NO |
| MS_net_OR_time_before_catheter | Time spent in operating theatre since admission to catheter placement. | cont | BASE | YES | YES |
| MS_net_OR_time_before_LM | Time spent in operating theatre before landmark time. | cont | TV | YES | YES |
| PAT_age | Patient age at admission | cont | BASE | YES | YES |
| PAT_gender_M | Patient gender | binary | BASE | YES | NO |
| MS_physical_ward_base | Admission ward | categ | BASE | YES | NO |
| CAT_lumens_total | Total number of lumens | cont | TV | YES | NO |
| MS_total_ICU_time_before_LM | Time spent in ICU during the catheter episode (and before landmark time) | cont | TV | YES | NO |
| LM_squared | The square number of LM | cont | TV | NO | YES |
| MED_L5_7d_B05BA10_combinations_LM | The interaction effect between MED_L5_7d_B05BA10_combinations and LM | cont | TV | NO | YES |
| CARE_NEU_GCS_score_last_LM | The interaction effect between CARE_NEU_GCS_score_last and LM | cont | TV | NO | YES |
| LAB_RBC_count_last_LM | The interaction effect between LAB_RBC_count_last and LM | cont | TV | NO | YES |
| MS_medical_specialty_bin_drop_Burns | The specific level of Variable MS_medical_specialty: Burns | binary | TV | NO | YES |
| MS_medical_specialty_bin_drop_Cardiac | The specific level of Variable MS_medical_specialty: Cardiac | binary | TV | NO | YES |
| MS_medical_specialty_bin_drop_ICU | The specific level of Variable MS_medical_specialty: ICU | binary | TV | NO | YES |
| MS_medical_specialty_bin_drop_Pediatrics | The specific level of Variable MS_medical_specialty: Pediatrics | binary | TV | NO | YES |
| MS_medical_specialty_bin_drop_Thoracic_Surgery | The specific level of Variable MS_medical_specialty: Thoracic Surgery | binary | TV | NO | YES |
| MS_medical_specialty_bin_drop_Traumatology | The specific level of Variable MS_medical_specialty: Traumatology | binary | TV | NO | YES |

Table 4: Variables included in the models; _binary_all means that all variable values encountered in the aggregation window are kept as binary values (0/1) for categorical variables

| Variable | Statistic | Train | Test | Total |
| --- | --- | --- | --- | --- |
|  |  | n = 61628 | n = 44544 | n = 106172 |
| ADM_admission_to_catheter | Median (IQR) | 0.9 (0.2 to 3.2) | 0.7 (0.1 to 2.9) | 0.8 (0.2 to 3.1) |
|  | Range (min, max) | (0 to 631.5) | (0 to 372.9) | (0 to 631.5) |
| ADM_admission_type_binary_all_Emergency | n(%) | 243153 (44.9) | 191362 (48.9) | 434515 (46.6) |
| ADM_nr_adm_past_180_days | Median (IQR) | 0.0 (0.0 to 2.0) | 0.0 (0.0 to 2.0) | 0.0 (0.0 to 2.0) |
|  | Range (min, max) | (0 to 25) | (0 to 25) | (0 to 25) |
| ADM_nr_emergency_adm_past_180_days | Median (IQR) | 0.0 (0.0 to 1.0) | 0.0 (0.0 to 1.0) | 0.0 (0.0 to 1.0) |
|  | Range (min, max) | (0 to 11) | (0 to 11) | (0 to 11) |
| ADM_unplanned_readmission | n(%) | 91600 (16.9) | 77293 (19.8) | 168893 (18.1) |
| CARE_ICU_ECMO | n(%) | 1426 (0.3) | 2080 (0.5) | 3506 (0.4) |
| CARE_ISO_binary_all_protective_isolation | n(%) | 29876 (5.5) | 18103 (4.6) | 47979 (5.1) |
| CARE_ISO_binary_all_source_isolation | n(%) | 51307 (9.5) | 25407 (6.5) | 76714 (8.2) |
| CARE_NEU_GCS_score_last | Median (IQR) | 14.5 (13.8 to 14.7) | 14.5 (13.7 to 14.7) | 14.5 (13.7 to 14.7) |
|  | Range (min, max) | (0 to 15) | (0 to 54.3) | (0 to 54.3) |
| CARE_PHY_drain | n(%) | 104069 (19.2) | 66765 (17.1) | 170834 (18.3) |
| CARE_SAF_freedom_restriction_categorical_last | complete, n(%) | 173090 (31.9) | 122449 (31.3) | 295539 (31.7) |
|  | partial, n(%) | 368725 (68.1) | 268904 (68.7) | 637629 (68.3) |
| CARE_SAF_mobility_assistance_binary_all_no_help | n(%) | 130573 (24.1) | 131250 (33.5) | 261823 (28.1) |
| CARE_SYM_RASS_max | Median (IQR) | 0.1 (0.0 to 0.2) | 0.1 (0.0 to 0.2) | 0.1 (0.0 to 0.2) |
|  | Range (min, max) | (-5 to 4) | (-5 to 4) | (-5 to 4) |
| CARE_SYM_pruritus_binary_all_YES | n(%) | 11317 (2.1) | 6935 (1.8) | 18252 (2.0) |
| CARE_VS_CVP_last | Median (IQR) | 7.2 (6.2 to 10.8) | 7.5 (6.3 to 11.1) | 7.3 (6.2 to 11.0) |
|  | Range (min, max) | (1 to 88) | (0.3 to 99) | (0.3 to 99) |
| CARE_VS_CVP_measured | n(%) | 79304 (14.6) | 60815 (15.5) | 140119 (15.0) |
| CARE_VS_MV | n(%) | 97377 (18.0) | 72723 (18.6) | 170100 (18.2) |
| CARE_VS_breathing_aid | n(%) | 107638 (19.9) | 82074 (21.0) | 189712 (20.3) |
| CARE_VS_diastolic_BP_last | Median (IQR) | 69.0 (60.0 to 77.0) | 69.0 (60.0 to 78.0) | 69.0 (60.0 to 77.7) |
|  | Range (min, max) | (31 to 195) | (31 to 206) | (31 to 206) |
| CARE_VS_heart_rate_max | Median (IQR) | 89.0 (79.0 to 104.0) | 90.0 (79.0 to 105.0) | 90.0 (79.0 to 104.0) |
|  | Range (min, max) | (2 to 344) | (1 to 328) | (1 to 344) |
| CARE_VS_oxygen_saturation_last | Median (IQR) | 96.6 (95.0 to 97.4) | 96.4 (95.0 to 97.4) | 96.5 (95.0 to 97.4) |
|  | Range (min, max) | (1 to 99.6) | (1 to 99) | (1 to 99.6) |
| CARE_VS_respiratory_rate_last | Median (IQR) | 17.3 (15.8 to 19.9) | 16.7 (15.7 to 20.0) | 17.0 (15.8 to 19.9) |
|  | Range (min, max) | (1 to 573) | (1 to 418) | (1 to 573) |
| CARE_VS_systolic_BP_last | Median (IQR) | 123.0 (108.0 to 137.0) | 122.0 (107.0 to 136.0) | 123.0 (108.0 to 137.0) |
|  | Range (min, max) | (31 to 344) | (31 to 344) | (31 to 344) |
| CARE_VS_temperature_max | Median (IQR) | 37.1 (36.7 to 37.6) | 37.1 (36.8 to 37.6) | 37.1 (36.7 to 37.6) |
|  | Range (min, max) | (30.1 to 43.1) | (30.7 to 43) | (30.1 to 43.1) |
| CARE_WND_wound_type_binary_all_closed_wound | n(%) | 27459 (5.1) | 23673 (6.0) | 51132 (5.5) |
| CARE_WND_wound_type_binary_all_open_wound | n(%) | 252570 (46.6) | 185772 (47.5) | 438342 (47.0) |
| CARE_WND_wound_type_binary_all_post_suture | n(%) | 5220 (1.0) | 3472 (0.9) | 8692 (0.9) |
| CARE_WND_wound_type_binary_all_suture | n(%) | 317587 (58.6) | 224294 (57.3) | 541881 (58.1) |
| CAT_bandage_change | n(%) | 199850 (36.9) | 136802 (35.0) | 336652 (36.1) |
| CAT_bandage_observation_binary_all_Bloody_or_Moist | n(%) | 17700 (3.3) | 10742 (2.7) | 28442 (3.0) |
| CAT_bandage_observation_binary_all_Normal | n(%) | 540035 (99.7) | 390498 (99.8) | 930533 (99.7) |
| CAT_bandage_observation_binary_all_Other_Hema_Pus_Loose_Necro | n(%) | 14946 (2.8) | 7650 (2.0) | 22596 (2.4) |
| CAT_bandage_observation_binary_all_Red | n(%) | 7499 (1.4) | 4400 (1.1) | 11899 (1.3) |
| CAT_bandage_type_binary_last_gauze | n(%) | 85407 (15.8) | 52368 (13.4) | 137775 (14.8) |
| CAT_catheter_location_binary_all_Arm | n(%) | 96298 (17.8) | 87526 (22.4) | 183824 (19.7) |
| CAT_catheter_location_binary_all_Collarbone | n(%) | 232546 (42.9) | 158328 (40.5) | 390874 (41.9) |
| CAT_catheter_location_binary_all_Groin | n(%) | 20226 (3.7) | 14214 (3.6) | 34440 (3.7) |
| CAT_catheter_location_binary_all_Navel | n(%) | 9326 (1.7) | 6812 (1.7) | 16138 (1.7) |
| CAT_catheter_location_binary_all_Neck | n(%) | 214504 (39.6) | 140417 (35.9) | 354921 (38.0) |
| CAT_catheter_location_binary_all_Other | n(%) | 12786 (2.4) | 18852 (4.8) | 31638 (3.4) |
| CAT_catheter_placement_binary_all_OR | n(%) | 202688 (37.4) | 150002 (38.3) | 352690 (37.8) |
| CAT_catheter_placement_binary_all_bedside | n(%) | 29050 (5.4) | 15546 (4.0) | 44596 (4.8) |
| CAT_consecutive_current_days_CVC | Median (IQR) | 0.0 (0.0 to 6.0) | 0.0 (0.0 to 5.0) | 0.0 (0.0 to 5.0) |
|  | Range (min, max) | (0 to 279) | (0 to 172) | (0 to 279) |
| CAT_consecutive_current_days_Dialysis_CVC | Median (IQR) | 0.0 (0.0 to 0.0) | 0.0 (0.0 to 0.0) | 0.0 (0.0 to 0.0) |
|  | Range (min, max) | (0 to 202) | (0 to 143) | (0 to 202) |
| CAT_consecutive_current_days_PICC | Median (IQR) | 0.0 (0.0 to 0.0) | 0.0 (0.0 to 0.0) | 0.0 (0.0 to 0.0) |
|  | Range (min, max) | (0 to 220) | (0 to 227) | (0 to 227) |
| CAT_consecutive_current_days_Port_a_cath | Median (IQR) | 0.0 (0.0 to 1.0) | 0.0 (0.0 to 2.0) | 0.0 (0.0 to 2.0) |
|  | Range (min, max) | (0 to 204) | (0 to 135) | (0 to 204) |
| CAT_consecutive_current_days_Tunneled_CVC | Median (IQR) | 0.0 (0.0 to 0.0) | 0.0 (0.0 to 0.0) | 0.0 (0.0 to 0.0) |
|  | Range (min, max) | (0 to 299) | (0 to 182) | (0 to 299) |
| CAT_days_since_last_bandage_changed | Median (IQR) | 1.0 (0.0 to 3.0) | 1.0 (0.0 to 3.0) | 1.0 (0.0 to 3.0) |
|  | Range (min, max) | (0 to 78) | (0 to 38) | (0 to 78) |
| CAT_days_since_last_bandage_obs | Median (IQR) | 0.0 (0.0 to 0.0) | 0.0 (0.0 to 0.0) | 0.0 (0.0 to 0.0) |
|  | Range (min, max) | (0 to 31) | (0 to 32) | (0 to 32) |
| CAT_days_since_last_tube_change | Median (IQR) | 1.0 (0.0 to 2.0) | 1.0 (0.0 to 3.0) | 1.0 (0.0 to 3.0) |
|  | Range (min, max) | (0 to 135) | (0 to 98) | (0 to 135) |
| CAT_lumens_flushed | Median (IQR) | 1.2 (1.0 to 1.8) | 1.3 (1.0 to 1.8) | 1.3 (1.0 to 1.8) |
|  | Range (min, max) | (1 to 6) | (1 to 7) | (1 to 7) |
| CAT_lumens_total | Median (IQR) | 2.0 (1.0 to 2.0) | 2.0 (1.0 to 3.0) | 2.0 (1.0 to 3.0) |
|  | Range (min, max) | (0 to 14) | (0 to 13) | (0 to 14) |
| CAT_needle_length_max | Median (IQR) | 16.9 (16.3 to 19.0) | 16.7 (16.1 to 17.5) | 16.8 (16.2 to 18.0) |
|  | Range (min, max) | (0 to 38) | (12 to 38) | (0 to 38) |
| CAT_nr_bandage_obersations | Median (IQR) | 3.0 (2.0 to 3.0) | 3.0 (2.0 to 4.0) | 3.0 (2.0 to 3.0) |
|  | Range (min, max) | (0 to 36) | (0 to 18) | (0 to 36) |
| CAT_number_central_lines | Median (IQR) | 1.0 (1.0 to 1.0) | 1.0 (1.0 to 1.0) | 1.0 (1.0 to 1.0) |
|  | Range (min, max) | (1 to 8) | (1 to 7) | (1 to 8) |
| CAT_number_peripheral_catheters | Median (IQR) | 0.0 (0.0 to 1.0) | 0.0 (0.0 to 2.0) | 0.0 (0.0 to 1.0) |
|  | Range (min, max) | (0 to 11) | (0 to 12) | (0 to 12) |
| CAT_tube_change | n(%) | 210517 (38.9) | 151350 (38.7) | 361867 (38.8) |
| CLABSI_history | n(%) | 11843 (2.2) | 6538 (1.7) | 18381 (2.0) |
| COM_COPD_before_LM | n(%) | 11530 (2.1) | 9920 (2.5) | 21450 (2.3) |
| COM_HIV_before_LM | n(%) | 1145 (0.2) | 1130 (0.3) | 2275 (0.2) |
| COM_ORG_heart_and_circulatory_system_before_LM | n(%) | 90421 (16.7) | 81985 (20.9) | 172406 (18.5) |
| COM_arterial_occlusive_disease_before_LM | n(%) | 10331 (1.9) | 12908 (3.3) | 23239 (2.5) |
| COM_heart_failure_before_LM | n(%) | 10894 (2.0) | 9768 (2.5) | 20662 (2.2) |
| COM_lymphoma_before_LM | n(%) | 27886 (5.1) | 20484 (5.2) | 48370 (5.2) |
| GEN_LM_month_categ | April, n(%) | 44904 (8.3) | 31505 (8.1) | 76409 (8.2) |
|  | August, n(%) | 44542 (8.2) | 32053 (8.2) | 76595 (8.2) |
|  | December, n(%) | 45753 (8.4) | 29500 (7.5) | 75253 (8.1) |
|  | February, n(%) | 43001 (7.9) | 32072 (8.2) | 75073 (8.0) |
|  | January, n(%) | 46137 (8.5) | 31508 (8.1) | 77645 (8.3) |
|  | July, n(%) | 43304 (8.0) | 33259 (8.5) | 76563 (8.2) |
|  | June, n(%) | 44362 (8.2) | 32939 (8.4) | 77301 (8.3) |
|  | March, n(%) | 47991 (8.9) | 35399 (9.0) | 83390 (8.9) |
|  | May, n(%) | 44914 (8.3) | 32568 (8.3) | 77482 (8.3) |
|  | November, n(%) | 44816 (8.3) | 31349 (8.0) | 76165 (8.2) |
|  | October, n(%) | 47215 (8.7) | 35206 (9.0) | 82421 (8.8) |
|  | September, n(%) | 44876 (8.3) | 33995 (8.7) | 78871 (8.5) |
| HC_101_Coronary_atherosclerosis_and_other_heart_disease | n(%) | 58694 (10.8) | 51136 (13.1) | 109830 (11.8) |
| HC_106_Cardiac_dysrhythmias | n(%) | 84234 (15.5) | 80550 (20.6) | 164784 (17.7) |
| HC_117_Other_circulatory_disease | n(%) | 88685 (16.4) | 88685 (22.7) | 177370 (19.0) |
| HC_155_Other_gastrointestinal_disorders | n(%) | 153825 (28.4) | 143418 (36.6) | 297243 (31.9) |
| HC_158_Chronic_kidney_disease | n(%) | 74147 (13.7) | 68097 (17.4) | 142244 (15.2) |
| HC_238_Complications_of_surgical_procedures_or_medical_care | n(%) | 144985 (26.8) | 135709 (34.7) | 280694 (30.1) |
| HC_253_Allergic_reactions | n(%) | 100604 (18.6) | 90343 (23.1) | 190947 (20.5) |
| HC_2617_Adverse_effects_of_medical_drugs | n(%) | 151656 (28.0) | 131027 (33.5) | 282683 (30.3) |
| HC_3_Bacterial_infection_unspecified_site | n(%) | 131189 (24.2) | 112813 (28.8) | 244002 (26.1) |
| HC_42_Secondary_malignancies | n(%) | 104705 (19.3) | 88445 (22.6) | 193150 (20.7) |
| HC_45_Maintenance_chemotherapy_radiotherapy | n(%) | 133618 (24.7) | 111971 (28.6) | 245589 (26.3) |
| HC_53_Disorders_of_lipid_metabolism | n(%) | 100804 (18.6) | 86137 (22.0) | 186941 (20.0) |
| HC_55_Fluid_and_electrolyte_disorders | n(%) | 153474 (28.3) | 141355 (36.1) | 294829 (31.6) |
| HC_58_Other_nutritional_endocrine_and_metabolic_disorders | n(%) | 229732 (42.4) | 219260 (56.0) | 448992 (48.1) |
| HC_59_Deficiency_and_other_anemia | n(%) | 141022 (26.0) | 118697 (30.3) | 259719 (27.8) |
| HC_96_Heart_valve_disorders | n(%) | 48058 (8.9) | 48358 (12.4) | 96416 (10.3) |
| HC_98_Essential_hypertension | n(%) | 118916 (21.9) | 96131 (24.6) | 215047 (23.0) |
| LAB_ALT_last | Median (IQR) | 30.7 (23.2 to 42.0) | 30.4 (22.1 to 41.8) | 30.6 (23.0 to 42.0) |
|  | Range (min, max) | (5 to 18526) | (5 to 21204) | (5 to 21204) |
| LAB_APTT_last | Median (IQR) | 31.6 (30.3 to 34.5) | 31.7 (30.2 to 35.4) | 31.6 (30.2 to 35.0) |
|  | Range (min, max) | (16.1 to 180) | (16 to 180) | (16 to 180) |
| LAB_AST_last | Median (IQR) | 30.1 (24.5 to 41.0) | 30.0 (24.5 to 41.7) | 30.1 (24.5 to 41.3) |
|  | Range (min, max) | (5 to 68420) | (5 to 33732) | (5 to 68420) |
| LAB_CK_last | Median (IQR) | 146.8 (74.7 to 341.8) | 188.9 (77.2 to 404.7) | 162.3 (75.7 to 370.4) |
|  | Range (min, max) | (7 to 490100) | (7 to 264800) | (7 to 490100) |
| LAB_CRP_last | Median (IQR) | 41.3 (21.4 to 78.6) | 40.9 (21.1 to 76.2) | 41.1 (21.3 to 77.6) |
|  | Range (min, max) | (0.3 to 3731) | (0.3 to 685.3) | (0.3 to 3731) |
| LAB_D_dimer_last | Median (IQR) | 2622.4 (2187.2 to 3367.2) | 2689.8 (2246.1 to 3404.7) | 2650.7 (2212.0 to 3384.4) |
|  | Range (min, max) | (215 to 7650) | (215 to 284780) | (215 to 284780) |
| LAB_Hemoglobine_last | Median (IQR) | 9.9 (9.1 to 10.6) | 9.8 (8.9 to 10.5) | 9.9 (9.0 to 10.5) |
|  | Range (min, max) | (0.1 to 26.1) | (0.3 to 26.6) | (0.1 to 26.6) |
| LAB_LDH_last | Median (IQR) | 229.0 (203.0 to 294.9) | 236.0 (206.1 to 311.0) | 231.8 (204.4 to 301.2) |
|  | Range (min, max) | (10 to 114000) | (57 to 51420) | (10 to 114000) |
| LAB_NT_proBNP_last | Median (IQR) | 4531.5 (2840.2 to 7992.4) | 5110.4 (3249.6 to 9042.8) | 4774.5 (3003.8 to 8416.1) |
|  | Range (min, max) | (9 to 308160) | (10 to 227675) | (9 to 308160) |
| LAB_O2_saturation_last | Median (IQR) | 96.2 (94.5 to 96.7) | 96.0 (94.4 to 96.7) | 96.1 (94.4 to 96.7) |
|  | Range (min, max) | (2.6 to 100) | (4 to 100) | (2.6 to 100) |
| LAB_PT_INR_last | Median (IQR) | 1.2 (1.1 to 1.3) | 1.2 (1.1 to 1.3) | 1.2 (1.1 to 1.3) |
|  | Range (min, max) | (0.7 to 15) | (0.8 to 15) | (0.7 to 15) |
| LAB_PT_percent_last | Median (IQR) | 79.0 (71.9 to 86.6) | 79.6 (71.7 to 86.4) | 79.2 (71.8 to 86.5) |
|  | Range (min, max) | (15 to 150) | (8 to 150) | (8 to 150) |
| LAB_PT_sec_last | Median (IQR) | 13.4 (12.6 to 14.6) | 13.5 (12.6 to 14.8) | 13.4 (12.6 to 14.6) |
|  | Range (min, max) | (8 to 120) | (8 to 120) | (8 to 120) |
| LAB_Platelet_count_last | Median (IQR) | 252.0 (185.0 to 320.0) | 246.0 (179.0 to 310.1) | 249.3 (182.1 to 316.0) |
|  | Range (min, max) | (1 to 2829) | (1 to 2268) | (1 to 2829) |
| LAB_RBC_count_last | Median (IQR) | 3.4 (3.1 to 3.6) | 3.3 (3.0 to 3.6) | 3.4 (3.0 to 3.6) |
|  | Range (min, max) | (0 to 9) | (0.3 to 8.6) | (0 to 9) |
| LAB_SPE_albumin_alpha_1_globulin_last | Median (IQR) | 4.4 (4.0 to 4.9) | 4.5 (4.1 to 4.9) | 4.4 (4.0 to 4.9) |
|  | Range (min, max) | (0.9 to 15) | (0.4 to 12.3) | (0.4 to 15) |
| LAB_TSH_last | Median (IQR) | 3.2 (2.7 to 3.9) | 3.3 (2.8 to 4.1) | 3.3 (2.8 to 4.0) |
|  | Range (min, max) | (0 to 183.6) | (0 to 197.5) | (0 to 197.5) |
| LAB_WBC_Monocytes_last | Median (IQR) | 0.7 (0.5 to 0.9) | 0.7 (0.5 to 0.9) | 0.7 (0.5 to 0.9) |
|  | Range (min, max) | (0 to 166.5) | (0 to 59.8) | (0 to 166.5) |
| LAB_WBC_Neutrophils_last | Median (IQR) | 5.3 (3.8 to 7.7) | 5.3 (3.9 to 7.7) | 5.3 (3.8 to 7.7) |
|  | Range (min, max) | (0 to 150) | (0 to 170.5) | (0 to 170.5) |
| LAB_WBC_count_last | Median (IQR) | 7.9 (5.9 to 10.5) | 8.0 (6.0 to 10.7) | 7.9 (6.0 to 10.6) |
|  | Range (min, max) | (0.1 to 445.1) | (0.1 to 301.7) | (0.1 to 445.1) |
| LAB_aspergillus_ag_last | Median (IQR) | 0.1 (0.1 to 0.2) | 0.1 (0.1 to 0.2) | 0.1 (0.1 to 0.2) |
|  | Range (min, max) | (0 to 8.1) | (0 to 7.7) | (0 to 8.1) |
| LAB_bilirubin_last | Median (IQR) | 0.5 (0.4 to 0.8) | 0.5 (0.4 to 0.8) | 0.5 (0.4 to 0.8) |
|  | Range (min, max) | (0.2 to 90.8) | (0.2 to 63.3) | (0.2 to 90.8) |
| LAB_creatinine_clearance_last | Median (IQR) | 68.9 (56.2 to 83.7) | 66.9 (53.0 to 81.6) | 68.1 (54.8 to 82.8) |
|  | Range (min, max) | (0 to 1008.7) | (0 to 633.1) | (0 to 1008.7) |
| LAB_creatinine_last | Median (IQR) | 0.8 (0.7 to 1.1) | 0.9 (0.7 to 1.1) | 0.9 (0.7 to 1.1) |
|  | Range (min, max) | (0.1 to 75.9) | (0.1 to 27.9) | (0.1 to 75.9) |
| LAB_ferritin_last | Median (IQR) | 753.5 (529.7 to 1397.0) | 803.0 (548.6 to 1613.2) | 773.5 (537.1 to 1486.2) |
|  | Range (min, max) | (4 to 1e+05) | (1 to 422600) | (1 to 422600) |
| LAB_fibrinogen_last | Median (IQR) | 3.6 (3.1 to 4.0) | 3.6 (3.1 to 4.0) | 3.6 (3.1 to 4.0) |
|  | Range (min, max) | (0.3 to 12) | (0.3 to 12) | (0.3 to 12) |
| LAB_glucose_arterial_last | Median (IQR) | 121.2 (110.2 to 125.7) | 122.4 (116.0 to 127.5) | 121.7 (112.4 to 126.4) |
|  | Range (min, max) | (9 to 820) | (10 to 815) | (9 to 820) |
| LAB_glucose_last | Median (IQR) | 109.7 (102.8 to 120.2) | 111.0 (103.3 to 122.0) | 110.1 (103.0 to 121.0) |
|  | Range (min, max) | (2 to 1500) | (2 to 1500) | (2 to 1500) |
| LAB_haematocrit_last | Median (IQR) | 0.3 (0.3 to 0.3) | 0.3 (0.3 to 0.3) | 0.3 (0.3 to 0.3) |
|  | Range (min, max) | (0 to 0.8) | (0 to 0.8) | (0 to 0.8) |
| LAB_is_neutropenia | n(%) | 16862 (3.1) | 12849 (3.3) | 29711 (3.2) |
| LAB_natrium_last | Median (IQR) | 138.7 (137.6 to 139.9) | 138.7 (137.6 to 139.9) | 138.7 (137.6 to 139.9) |
|  | Range (min, max) | (90.9 to 194) | (80 to 209.2) | (80 to 209.2) |
| LAB_pH_last | Median (IQR) | 7.5 (7.4 to 7.5) | 7.5 (7.4 to 7.5) | 7.5 (7.4 to 7.5) |
|  | Range (min, max) | (6.4 to 7.8) | (6.6 to 7.7) | (6.4 to 7.8) |
| LAB_pO2_last | Median (IQR) | 88.3 (78.4 to 93.6) | 87.8 (78.6 to 94.0) | 88.1 (78.5 to 93.7) |
|  | Range (min, max) | (10 to 656) | (10.1 to 632) | (10 to 656) |
| LAB_potassium_last | Median (IQR) | 3.9 (3.7 to 4.2) | 4.0 (3.7 to 4.2) | 3.9 (3.7 to 4.2) |
|  | Range (min, max) | (1.5 to 36.9) | (1.8 to 72.6) | (1.5 to 72.6) |
| LAB_troponine_T_last | Median (IQR) | 0.1 (0.1 to 0.3) | 0.1 (0.1 to 0.3) | 0.1 (0.1 to 0.3) |
|  | Range (min, max) | (0 to 174) | (0 to 65.8) | (0 to 174) |
| LAB_urea_last | Median (IQR) | 31.0 (25.2 to 44.7) | 33.0 (26.4 to 48.5) | 32.0 (25.9 to 46.0) |
|  | Range (min, max) | (3 to 608) | (3 to 589) | (3 to 608) |
| LAB_vancomycine_last | Median (IQR) | 13.7 (12.7 to 17.2) | 14.1 (12.9 to 17.8) | 13.8 (12.8 to 17.5) |
|  | Range (min, max) | (1.7 to 417) | (4 to 738) | (1.7 to 738) |
| LM | Median (IQR) | 5.0 (2.0 to 13.0) | 5.0 (2.0 to 13.0) | 5.0 (2.0 to 13.0) |
|  | Range (min, max) | (0 to 298) | (0 to 254) | (0 to 298) |
| MB_infection_time_window_binary_all_GI | n(%) | 19090 (3.5) | 15215 (3.9) | 34305 (3.7) |
| MB_infection_time_window_binary_all_blood | n(%) | 55086 (10.2) | 40199 (10.3) | 95285 (10.2) |
| MB_infection_time_window_binary_all_catheter | n(%) | 8301 (1.5) | 2772 (0.7) | 11073 (1.2) |
| MB_infection_time_window_binary_all_deep_tissue | n(%) | 25578 (4.7) | 18972 (4.8) | 44550 (4.8) |
| MB_infection_time_window_binary_all_drain | n(%) | 9632 (1.8) | 5810 (1.5) | 15442 (1.7) |
| MB_infection_time_window_binary_all_lung | n(%) | 53291 (9.8) | 42911 (11.0) | 96202 (10.3) |
| MB_infection_time_window_binary_all_skin | n(%) | 61859 (11.4) | 38617 (9.9) | 100476 (10.8) |
| MB_infection_time_window_binary_all_sputum | n(%) | 35028 (6.5) | 21143 (5.4) | 56171 (6.0) |
| MB_infection_time_window_binary_all_urogen | n(%) | 76304 (14.1) | 55751 (14.2) | 132055 (14.2) |
| MB_number_contaminants_in_blood | Median (IQR) | 0.0 (0.0 to 0.0) | 0.0 (0.0 to 0.0) | 0.0 (0.0 to 0.0) |
|  | Range (min, max) | (0 to 15) | (0 to 15) | (0 to 15) |
| MB_other_infection_than_BSI_during_window | n(%) | 197498 (36.5) | 145096 (37.1) | 342594 (36.7) |
| MED_7d_CITRA_LOCK | n(%) | 8037 (1.5) | 6649 (1.7) | 14686 (1.6) |
| MED_7d_VANCO_CEFTA_LOCK | n(%) | 2828 (0.5) | 2154 (0.6) | 4982 (0.5) |
| MED_7d_immunoglobulins | n(%) | 17437 (3.2) | 11812 (3.0) | 29249 (3.1) |
| MED_7d_number_of_IV_med | Median (IQR) | 9.0 (5.0 to 14.0) | 8.0 (4.0 to 14.0) | 8.0 (5.0 to 14.0) |
|  | Range (min, max) | (0 to 51) | (0 to 47) | (0 to 51) |
| MED_7d_number_of_ORAL_med | Median (IQR) | 6.0 (3.0 to 9.0) | 6.0 (3.0 to 10.0) | 6.0 (3.0 to 9.0) |
|  | Range (min, max) | (0 to 28) | (0 to 29) | (0 to 29) |
| MED_L2_7d_H02_CORTICOSTEROIDS_FOR_SYSTEMIC_USE | n(%) | 221610 (40.9) | 182958 (46.8) | 404568 (43.4) |
| MED_L2_7d_J01_ANTIBACTERIALS_FOR_SYSTEMIC_USE | n(%) | 415342 (76.7) | 300312 (76.7) | 715654 (76.7) |
| MED_L2_7d_J02_ANTIMYCOTICS_FOR_SYSTEMIC_USE | n(%) | 77146 (14.2) | 57169 (14.6) | 134315 (14.4) |
| MED_L2_7d_J04_ANTIMYCOBACTERIALS | n(%) | 8837 (1.6) | 9455 (2.4) | 18292 (2.0) |
| MED_L2_7d_J05_ANTIVIRALS_FOR_SYSTEMIC_USE | n(%) | 45289 (8.4) | 39903 (10.2) | 85192 (9.1) |
| MED_L2_7d_J06_IMMUNE_SERA_AND_IMMUNOGLOBULINS | n(%) | 10475 (1.9) | 5239 (1.3) | 15714 (1.7) |
| MED_L2_7d_L01_ANTINEOPLASTIC_AGENTS | n(%) | 60513 (11.2) | 44942 (11.5) | 105455 (11.3) |
| MED_L2_7d_L04_IMMUNOSUPPRESSANTS | n(%) | 54061 (10.0) | 44752 (11.4) | 98813 (10.6) |
| MED_L5_7d_B05BA01_amino_acids | n(%) | 14581 (2.7) | 4448 (1.1) | 19029 (2.0) |
| MED_L5_7d_B05BA02_fat_emulsions | n(%) | 21858 (4.0) | 16690 (4.3) | 38548 (4.1) |
| MED_L5_7d_B05BA03_carbohydrates | n(%) | 297037 (54.8) | 200441 (51.2) | 497478 (53.3) |
| MED_L5_7d_B05BA10_combinations | n(%) | 117315 (21.7) | 72271 (18.5) | 189586 (20.3) |
| MS_ICU_time_before_catheter | Median (IQR) | 0.0 (0.0 to 0.0) | 0.0 (0.0 to 0.0) | 0.0 (0.0 to 0.0) |
|  | Range (min, max) | (0 to 281) | (0 to 357.2) | (0 to 357.2) |
| MS_alternative_flag | n(%) | 15596 (2.9) | 14234 (3.6) | 29830 (3.2) |
| MS_is_24h_net_OR_unit | n(%) | 49596 (9.2) | 40945 (10.5) | 90541 (9.7) |
| MS_is_ICU_unit | n(%) | 113716 (21.0) | 81912 (20.9) | 195628 (21.0) |
| MS_medical_specialty | Abdomen, n(%) | 98463 (18.2) | 63110 (16.1) | 161573 (17.3) |
|  | Burns, n(%) | 3761 (0.7) | 2727 (0.7) | 6488 (0.7) |
|  | Cardiac, n(%) | 37359 (6.9) | 28970 (7.4) | 66329 (7.1) |
|  | Endocrinology, n(%) | 2828 (0.5) | 2576 (0.7) | 5404 (0.6) |
|  | Geriatrics, n(%) | 15507 (2.9) | 11716 (3.0) | 27223 (2.9) |
|  | Gynecology, n(%) | 22026 (4.1) | 18576 (4.7) | 40602 (4.4) |
|  | Hematology, n(%) | 41590 (7.7) | 33539 (8.6) | 75129 (8.1) |
|  | ICU, n(%) | 61254 (11.3) | 42009 (10.7) | 103263 (11.1) |
|  | Internal Medicine, n(%) | 25908 (4.8) | 21680 (5.5) | 47588 (5.1) |
|  | Neonatology, n(%) | 17071 (3.2) | 12650 (3.2) | 29721 (3.2) |
|  | Nephrology, n(%) | 15034 (2.8) | 11527 (2.9) | 26561 (2.8) |
|  | Neuro, n(%) | 9999 (1.8) | 7312 (1.9) | 17311 (1.9) |
|  | ORL, n(%) | 3979 (0.7) | 3380 (0.9) | 7359 (0.8) |
|  | Oncology, n(%) | 38719 (7.1) | 27405 (7.0) | 66124 (7.1) |
|  | Operation, n(%) | 2310 (0.4) | 1199 (0.3) | 3509 (0.4) |
|  | Other, n(%) | 7817 (1.4) | 7082 (1.8) | 14899 (1.6) |
|  | Pediatrics, n(%) | 30046 (5.5) | 21556 (5.5) | 51602 (5.5) |
|  | Pneumology, n(%) | 40102 (7.4) | 33481 (8.6) | 73583 (7.9) |
|  | Thoracic Surgery, n(%) | 20274 (3.7) | 10978 (2.8) | 31252 (3.3) |
|  | Transplant, n(%) | 3970 (0.7) | 2697 (0.7) | 6667 (0.7) |
|  | Traumatology, n(%) | 35296 (6.5) | 22366 (5.7) | 57662 (6.2) |
|  | Urology, n(%) | 8502 (1.6) | 4817 (1.2) | 13319 (1.4) |
| MS_net_OR_time_before_LM | Median (IQR) | 0.0 (0.0 to 0.0) | 0.0 (0.0 to 0.0) | 0.0 (0.0 to 0.0) |
|  | Range (min, max) | (0 to 1.7) | (0 to 1.1) | (0 to 1.7) |
| MS_net_OR_time_before_catheter | Median (IQR) | 0.0 (0.0 to 0.1) | 0.0 (0.0 to 0.1) | 0.0 (0.0 to 0.1) |
|  | Range (min, max) | (0 to 4.4) | (0 to 6.6) | (0 to 6.6) |
| MS_physical_ward_base | Abdomen, n(%) | 79282 (14.6) | 49285 (12.6) | 128567 (13.8) |
|  | Cardiac, n(%) | 21607 (4.0) | 24229 (6.2) | 45836 (4.9) |
|  | Emergency, n(%) | 19843 (3.7) | 38601 (9.9) | 58444 (6.3) |
|  | Endocrinology, n(%) | 6033 (1.1) | 3761 (1.0) | 9794 (1.0) |
|  | Geriatrics, n(%) | 8843 (1.6) | 7766 (2.0) | 16609 (1.8) |
|  | Gynecology, n(%) | 17754 (3.3) | 13429 (3.4) | 31183 (3.3) |
|  | Hematology, n(%) | 32057 (5.9) | 26074 (6.7) | 58131 (6.2) |
|  | ICU, n(%) | 122657 (22.6) | 58298 (14.9) | 180955 (19.4) |
|  | Internal Medicine, n(%) | 6476 (1.2) | 5309 (1.4) | 11785 (1.3) |
|  | Neonatology, n(%) | 18039 (3.3) | 13994 (3.6) | 32033 (3.4) |
|  | Nephrology, n(%) | 4129 (0.8) | 3165 (0.8) | 7294 (0.8) |
|  | Neuro, n(%) | 9362 (1.7) | 7496 (1.9) | 16858 (1.8) |
|  | ORL, n(%) | 4907 (0.9) | 4512 (1.2) | 9419 (1.0) |
|  | Oncology, n(%) | 54241 (10.0) | 36293 (9.3) | 90534 (9.7) |
|  | Other, n(%) | 473 (0.1) | 441 (0.1) | 914 (0.1) |
|  | Pediatrics, n(%) | 30334 (5.6) | 7452 (1.9) | 37786 (4.0) |
|  | Pneumology, n(%) | 31441 (5.8) | 25685 (6.6) | 57126 (6.1) |
|  | Thoracic Surgery, n(%) | 20549 (3.8) | 12952 (3.3) | 33501 (3.6) |
|  | Transplant, n(%) | 10145 (1.9) | 7766 (2.0) | 17911 (1.9) |
|  | Traumatology, n(%) | 33993 (6.3) | 20996 (5.4) | 54989 (5.9) |
|  | UNKNOWN_VALUE, n(%) | 0 (0.0) | 18612 (4.8) | 18612 (2.0) |
|  | Urology, n(%) | 9650 (1.8) | 5237 (1.3) | 14887 (1.6) |
| MS_total_ICU_time_before_LM | Median (IQR) | 0.0 (0.0 to 2.0) | 0.0 (0.0 to 2.1) | 0.0 (0.0 to 2.0) |
|  | Range (min, max) | (0 to 277.4) | (0 to 178.9) | (0 to 277.4) |
| PAT_age | Median (IQR) | 61.0 (46.0 to 71.0) | 62.0 (46.0 to 72.0) | 61.0 (46.0 to 71.0) |
|  | Range (min, max) | (0 to 101) | (0 to 100) | (0 to 101) |
| PAT_gender_M | n(%) | 290545 (53.6) | 208855 (53.4) | 499400 (53.5) |
| eventtime | Median (IQR) | 13.2 (6.7 to 26.7) | 13.9 (7.0 to 27.8) | 13.5 (6.8 to 27.1) |
|  | Range (min, max) | (0 to 298) | (0 to 255) | (0 to 298) |

Table 5: Descriptive table of all variables included in the models

### Supplementary material 3 - Missing data imputation

Table 6 presents the missing data information in the training and test sets. Specific laboratory test results (AST, ALT, WBC and glucose), scores (RASS) and respiratory indicators (oxygen saturation and respiratory rate) show improved registration during the test period (2018 - 2020), most probably partly attributable to closer patient monitoring during the COVID-19 pandemic.

| Feature | Number present | | Percentage missing | | Difference percent |
| --- | --- | --- | --- | --- | --- |
|  | Training | Test | Training | Test |  |
| PAT_age | 541797 | 391345 | 0% | 0% | 0% |
| MS_alternative_flag | 541326 | 390436 | 0.1% | 0.2% | -0.1% |
| MS_medical_specialty | 541326 | 390436 | 0.1% | 0.2% | -0.1% |
| MS_physical_ward_base | 538504 | 387700 | 0.6% | 0.9% | -0.3% |
| ADM_admission_type_binary_all_Emergency | 535997 | 385232 | 1.1% | 1.6% | -0.5% |
| CARE_VS_temperature_max | 515385 | 374487 | 4.9% | 4.3% | 0.6% |
| CARE_VS_heart_rate_max | 507080 | 371987 | 6.4% | 4.9% | 1.5% |
| CARE_VS_systolic_BP_last | 497423 | 367386 | 8.2% | 6.1% | 2.1% |
| CARE_VS_diastolic_BP_last | 496673 | 366927 | 8.3% | 6.2% | 2.1% |
| CAT_bandage_type_binary_last_gauze | 461416 | 332895 | 14.8% | 14.9% | -0.1% |
| CAT_bandage_observation_binary_all_Bloody_or_Moist | 454474 | 328980 | 16.1% | 15.9% | 0.2% |
| CAT_bandage_observation_binary_all_Normal | 454474 | 328980 | 16.1% | 15.9% | 0.2% |
| CAT_bandage_observation_binary_all_Other_Hema_Pus_Loose_Necro | 454474 | 328980 | 16.1% | 15.9% | 0.2% |
| CAT_bandage_observation_binary_all_Red | 454474 | 328980 | 16.1% | 15.9% | 0.2% |
| CARE_SAF_mobility_assistance_binary_all_no_help | 414549 | 316437 | 23.5% | 19.1% | 4.3% |
| LAB_potassium_last | 326279 | 237722 | 39.8% | 39.3% | 0.5% |
| LAB_natrium_last | 326065 | 237424 | 39.8% | 39.3% | 0.5% |
| LAB_is_neutropenia | 324511 | 237641 | 40.1% | 39.3% | 0.8% |
| LAB_creatinine_last | 324508 | 236327 | 40.1% | 39.6% | 0.5% |
| LAB_Hemoglobine_last | 323921 | 236849 | 40.2% | 39.5% | 0.7% |
| LAB_urea_last | 323464 | 234947 | 40.3% | 40% | 0.3% |
| LAB_WBC_count_last | 322755 | 235969 | 40.4% | 39.7% | 0.7% |
| LAB_CRP_last | 319967 | 234284 | 40.9% | 40.1% | 0.8% |
| LAB_Platelet_count_last | 319555 | 234402 | 41% | 40.1% | 0.9% |
| LAB_RBC_count_last | 319547 | 231744 | 41% | 40.8% | 0.2% |
| LAB_haematocrit_last | 318844 | 231718 | 41.2% | 40.8% | 0.4% |
| CARE_VS_oxygen_saturation_last | 283323 | 280818 | 47.7% | 28.2% | 19.5% |
| CARE_WND_wound_type_binary_all_closed_wound | 277270 | 199036 | 48.8% | 49.1% | -0.3% |
| CARE_WND_wound_type_binary_all_open_wound | 277270 | 199036 | 48.8% | 49.1% | -0.3% |
| CARE_WND_wound_type_binary_all_post_suture | 277270 | 199036 | 48.8% | 49.1% | -0.3% |
| CARE_WND_wound_type_binary_all_suture | 277270 | 199036 | 48.8% | 49.1% | -0.3% |
| LAB_bilirubin_last | 234732 | 183100 | 56.7% | 53.2% | 3.5% |
| LAB_AST_last | 198719 | 167022 | 63.3% | 57.3% | 6% |
| LAB_ALT_last | 198579 | 167154 | 63.3% | 57.3% | 6.1% |
| LAB_LDH_last | 184383 | 149222 | 66% | 61.9% | 4.1% |
| LAB_PT_sec_last | 180314 | 128197 | 66.7% | 67.2% | -0.5% |
| LAB_PT_INR_last | 180131 | 128055 | 66.8% | 67.3% | -0.5% |
| LAB_PT_percent_last | 180131 | 128057 | 66.8% | 67.3% | -0.5% |
| CARE_VS_respiratory_rate_last | 167202 | 235251 | 69.1% | 39.9% | 29.3% |
| LAB_WBC_Neutrophils_last | 162954 | 158881 | 69.9% | 59.4% | 10.5% |
| LAB_WBC_Monocytes_last | 162685 | 158842 | 70% | 59.4% | 10.6% |
| CAT_lumens_flushed | 151870 | 121740 | 72% | 68.9% | 3.1% |
| CARE_PHY_weight_mean | 151176 | 130177 | 72.1% | 66.7% | 5.4% |
| LAB_APTT_last | 150516 | 107421 | 72.2% | 72.6% | -0.3% |
| CAT_needle_length_max | 148847 | 111266 | 72.5% | 71.6% | 1% |
| CARE_SYM_RASS_max | 127908 | 148663 | 76.4% | 62% | 14.4% |
| LAB_pH_last | 117417 | 90000 | 78.3% | 77% | 1.3% |
| LAB_pO2_last | 117351 | 89966 | 78.3% | 77% | 1.3% |
| LAB_O2_saturation_last | 117341 | 89988 | 78.3% | 77% | 1.3% |
| LAB_glucose_arterial_last | 116316 | 89420 | 78.5% | 77.2% | 1.4% |
| LAB_glucose_last | 109182 | 101478 | 79.8% | 74.1% | 5.8% |
| CARE_NEU_GCS_score_last | 104919 | 85541 | 80.6% | 78.1% | 2.5% |
| CARE_VS_CVP_last | 79304 | 60815 | 85.4% | 84.5% | 0.9% |
| CARE_SAF_freedom_restriction_categorical_last | 68853 | 36750 | 87.3% | 90.6% | -3.3% |
| LAB_CK_last | 56597 | 44447 | 89.6% | 88.6% | 0.9% |
| LAB_troponine_T_last | 28853 | 25715 | 94.7% | 93.4% | 1.2% |
| LAB_vancomycine_last | 25678 | 16886 | 95.3% | 95.7% | -0.4% |
| CARE_PHY_length_mean | 18403 | 16165 | 96.6% | 95.9% | 0.7% |
| LAB_fibrinogen_last | 16733 | 19156 | 96.9% | 95.1% | 1.8% |
| LAB_aspergillus_ag_last | 15550 | 15058 | 97.1% | 96.2% | 1% |
| LAB_TSH_last | 10478 | 8319 | 98.1% | 97.9% | 0.2% |
| LAB_ferritin_last | 9369 | 11538 | 98.3% | 97.1% | 1.2% |
| LAB_creatinine_clearance_last | 7721 | 5053 | 98.6% | 98.7% | -0.1% |
| LAB_D_dimer_last | 4354 | 7963 | 99.2% | 98% | 1.2% |
| LAB_SPE_albumin_alpha_1_globulin_last | 3440 | 2315 | 99.4% | 99.4% | 0% |
| LAB_NT_proBNP_last | 1287 | 1498 | 99.8% | 99.6% | 0.1% |

Table 6: Missing values in training and test sets

The number of catheter lumens were imputed during the data preparation with typical (normal) values for each catheter type based on clinical knowledge and the total number of lumens (for all catheters) was calculated and kept as a variable. Missing data in other variables were subsequently imputed using missForestPredict^14^^,^^15^ on the landmark dataset using the complete set of variables. The missing values were first initialized using mean/mode at baseline (start of the catheter episode), followed by last observation carried forward (LOCF) imputation and then iteratively imputed using missForestPredict with default hyperparameter values, except maximum tree depth which was set to 10 for computational speed. The outcome was not included in the imputation process. The test set is initialized with mean/mode at baseline and LOCF imputation and imputed using missForestPredict models learned on the training set.

The out-of-bag (OOB) normalized mean square errors (NMSE) for all variables (grouped per variable category) are presented in Figure 3, Figure 4 and Figure 5. The OOB NMSE is an indication of imputation quality for each variable.

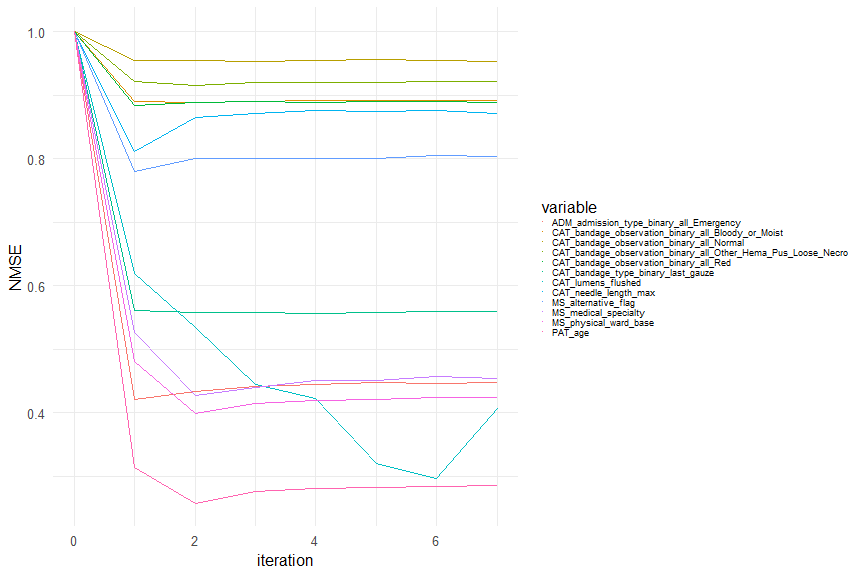

Figure 3: Out-of-bag (OOB) normalized mean square errors (NMSE) for patient, admission, medical specialty and catheter variables

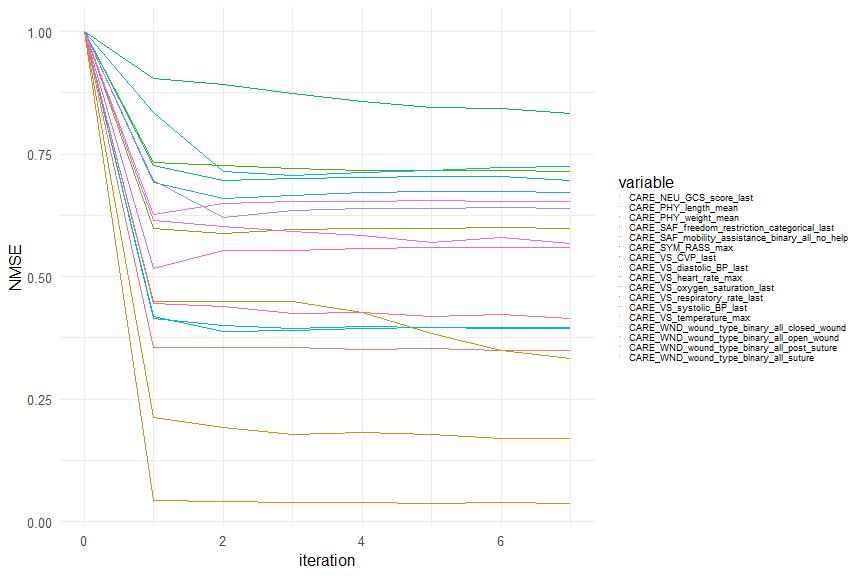

Figure 4: Out-of-bag (OOB) normalized mean square errors (NMSE) for care observations variables

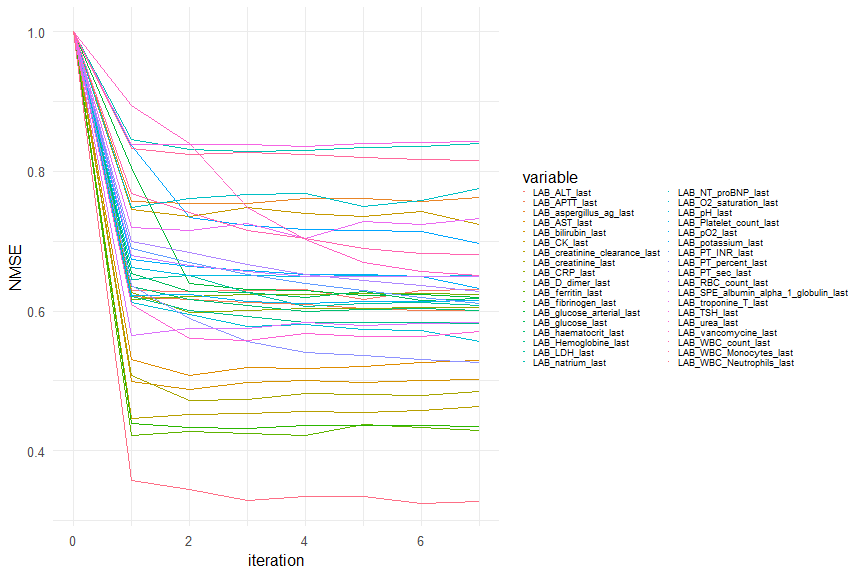

Figure 5: Out-of-bag (OOB) normalized mean square errors (NMSE) for laboratory results variables

### Supplementary material 4 - Selection of the limited set of 62 variables

The limited set of variables were selected based on the lasso regression results on the playground data from 2012 to 2013. We conducted variable selection based on 100 repeated train-test splits on the playground data. For each split, we used a random sample of two-thirds of hospital admissions from the landmark dataset, which was imputed using missForestPredict, as the training set. Each hospital admission’s landmarks and catheter episodes were entirely included in the training set or the test set. In the variable selection process we utilized only the training data. We performed cross-validation (split by hospital admission identifier) to identify the optimal lambda for the Lasso model, subsequently fitting the model with this lambda. Candidate predictors include not only the original predictors in the dataset, but also lagged and differentiated values, as well as missing value indicators (grouped based on panel, for example, some lab tests which all belong to liver panel like bilirubin and lactate dehydrogenase (LDH) share same missing indicator). Moreover, variable *LM* is forced into the Lasso model. From the fitted model, we extracted the coefficients from the fitted Lasso model and computed the standard deviation of each predictor variable, then obtained the standardized coefficients by multiplying the coefficients by the corresponding standard deviations of the predictor variables. Then we assessed the relative importance of the predictors and selected the top 20 important variables. Finally, we summarized the frequency of each variable selected by LASSO across all training sets to identify consistently important variables. We firstly performed the selection using all original predictors, lagged variables, differentiated variables as well as missing indicators, and take the top 10 most frequently selected variables out of 100 train-test splits, then added their time interaction term with *LM* as well as an interaction effect of age with gender, finally performed a Lasso selection again adding the above interactions.

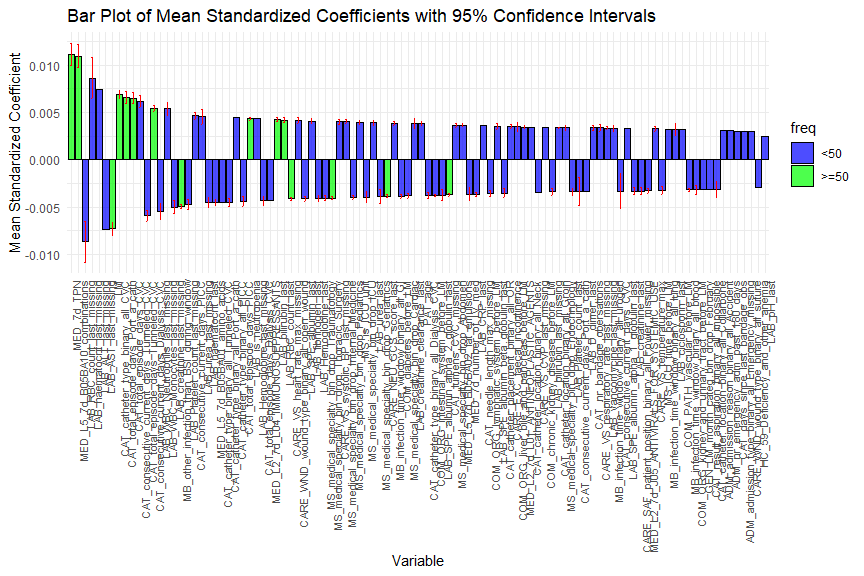

Figure 6: The mean standardized coefficients of lasso regression results

Figure 6 presents the mean standard coefficients of the lasso regression results. We sorted them based on the absolute value of the mean standard coefficients, and marked those which were selected in more than half of the 100 runs in green, otherwise in blue. Their confidence intervals across these splits were shown in red as error bars. According to the Lasso regression results, we selected those which were selected in more than half of the 100 runs into our limited set of predictors.

Moreover, we selected the top 50 variables from the multinomial RF model built on playground data and binarized the categorical variables (e.g., MS_medical_specialty), which resulted in a list of 78 variables. We used the pmsampsize package in R to estimate the minimum sample size required for developing either a logistic regression or Cox proportional hazard model. According to the previous systematic review of CLABSI prediction models, the mean of optimism-corrected AUC is 0.75 in the studies with similar EHR setting. Assuming we can obtain a c-statistic of 0.75 with 62 parameters and a prevalence of 1.31% for the 7-day CLABSI, a minimum of 49,070 and 49,260 unique catheter episodes are required to develop logistic and Cox models, respectively. Hence, our sample size of 61,628 is sufficiently large for static models if applying a 5-fold cross-validation. Given the utilization of the landmark approach for dynamic models, where information from adjacent landmarks is leveraged, we conclude that the same sample size rationale is applicable for dynamic models as well. Thus, to further reduce the list of variables, we performed Lasso regression again (forcing *LM* in the model), combining the list of 78 variables with the previously selected variables from Figure 6, and selected the top 5 ranked levels from variable MS_medical_specialty. According to the domain knowledge we collected from our expert panel, we also kept an individual level *Burns* from MS_medical_specialty. Eventually, we obtained a limited set of 62 predictors to fit our landmark cause-specific supermodel.

### Supplementary material 5 - Modelling details

#### Landmark cause-specific supermodel

The landmarking approach for dynamic prediction of survival was first described by van Houwelingen^30^. In brief, at a given landmark time $s$ from which a prediction is to be made, the data are restricted to individuals who have not yet had the event (in this case, CLABSI) or been censored. Values of predictor variables available up to the landmark time $s$, namely $Z\left( s \right)$, are used as covariates in a model for the probability of survival up to some time horizon $w$, conditional on survival to the landmark time $s$. In this case, the focus is on the predicted risk up to a single time horizon ($w$), that is, 7 days after the landmark time ($w$=7). To create the landmark model, risk prediction times of interest are first partitioned into different landmarks $\{s_{0},\ldots,s_{L}\}$. The sliding landmark datasets are created for each landmark s, using only the data of individuals still at risk at $s$, and applying artificial censoring at $s+w$ to these individuals.

In the previous study, we found that ignoring other competing risks would overestimate the risk of CLABSI^12^. Thus, we use the cause-specific model to predict the risk of CLABSI up to 7 days after each landmark time.

In this case, we assume the baseline hazard depends on s and this can be modelled by $\lambda_{0}\left( t│s \right)=\lambda_{0}\left( t \right)exp\left( \gamma\left( s \right) \right)$, where $\gamma\left( s \right)=\gamma_{1}\left( s \right)+\gamma_{2}\left( s \right)^{2}$. The landmark cause-specific supermodel can be fitted by applying the cause-specific Cox model for event J (J=1,…, j) to the stacked dataset and consider the baseline hazards $\lambda_{j0}\left( t \right)$ from each of the cause-specific Cox models as $\lambda_{j0}^{cs}\left( t│s \right)=\lambda_{j0}^{cs}\left( t \right)exp\left( \gamma_{j}\left( s \right) \right)$ and $\gamma_{j}\left( s \right)=\gamma_{j1}s+\gamma_{j2}s^{2}$. Thus, the cause-specific hazards of supermodel for event J from a landmark time $s\in\left[ s_{0},s_{L} \right]$ for time t ($s\leq t\leq s+w$) is:

$$\lambda_{j}^{cs}\left( t│Z\left( s \right),s \right)=\lambda_{j0}^{cs}\left( t│s \right)exp\left( \beta_{j}\left( s \right)Z\left( s \right) \right)=\lambda_{j0}^{cs}\left( t \right)exp\left( \gamma_{j}\left( s \right)+\beta_{j}\left( s \right)Z\left( s \right) \right)$$

Then w-day event free survival at any time point s in the window $\left[ s_{0},s_{L} \right]$ can be estimated with the exponential approximation:

$$S\left( s+w\mid Z\left( s \right),s \right)=exp\left( -\int_{s}^{s+w} \sum_{j=1}^{J} \lambda_{j}^{cs}\left( t\mid Z\left( s \right),s \right),dt \right)=exp\left( -\sum_{s\leq t_{i}\leq s+w} \sum_{j=1}^{J} \lambda_{j}^{cs}\left( t_{i}\mid Z\left( s \right),s \right) \right)$$

The cause-specific cumulative incidence for event J at any time point s in the window $\left[ s_{0},s_{L} \right]$ can be obtained with

$$F_{j}\left( s+w\mid Z\left( s \right),s \right)=\int_{s}^{s+w} \lambda_{j}^{cs}\left( t*\mid Z\left( s \right),s \right)S\left( t\mid Z\left( s \right),s \right) dt=\sum_{s\leq t_{i}\leq s+w} \lambda_{j}^{cs}\left( t_{i}\mid Z\left( s \right),s \right)S\left( t_{i}\mid Z\left( s \right),s \right)$$

where $t_{i}$ are event times in the training dataset used to fitting the landmark cause-specific supermodel.

#### Random forest (RF) and gradient boosting (XGB) models

The RF and XGB models treated each landmark within each catheter episode as an independent observation. The landmark number was incorporated into the dynamic models as a predictor, enabling models to account for time effects and interactions with time. Model based optimization was used for hyperparameter tuning, using the mlrMBO R package^31^ with 30 warmup iterations and 70 optimization iterations for RF models and 20 warmup and 30 optimization iterations for XGB models. Hyperparameters were tuned using the out-of-bag (OOB) binary logloss for 7-days CLABSI risk for the RF model and cross-validation multinomial logloss for 7-days events for XGB. We used nested 5-fold cross-validation for XGB tuning. Sampling for the in-bag and out-of-bag subsamples for RF and for the CV folds for XGB was based on the admission identifier, such that an admission falls completely in the training subsample or in the validation subsample. The hyperparameters used for the RF and XGB models are detailed in Table 7. For the RF model on the complete set we attempted variable selection by tuning the number of variables included in the model, as an additional hyperparameter. We first built a multinomial RF model using the randomForestSRC R package^32^ using deep trees (nodesize = 2) and sampling a third of the variables at each split of the tree. We then used the minimal depth of the maximal subtree^33^ to rank the variables from most important (on which splits in trees are done closer to the root node, having lower minimal depth) to least important (on which splits are done further down the tree, having higher minimal depth). At each tuning iteration in the mlrMBO framework, a combination of hyperparameters (including “number of variables”) was evaluated. First, the top “number of variables” were selected (as ranked by their minimal depth). Then, an RF model was built on this subset of variables using the other hyperparameters to be evaluated at this tuning iteration and using the ranger R package^28^. No explicit variable selection was performed for the XGB model, but regularization hyperparameters that might select out variables were included in the tuning procedure. Finally, the best hyperparameters resulting from the tuning procedure were used to build models on the outer CV “folds-in” (10 combinations of 9 “folds-in”, where one fold is kept out). The models are then tuned again entire training set using 5-fold CV for XGB and subsamples for RF. The hyperparameter values used to build the final models, as well as variable importance were saved.

| Model | Hyperparameter | Complete set | Limited set |
| --- | --- | --- | --- |
| RF | Number of trees | fixed, 1000 | fixed, 1000 |
|  | Subsampling type | fixed, sampling without replacement | fixed, sampling without replacement |
|  | Subsample size | tuned, 30% to 80% of the number of observations | tuned, 30% to 80% of the number of observations |
|  | Number of variables to split at each node (mtry) | tuned, 2% to 90% of the total number of variables) | tuned, 2 to 75% of the total number of variables) |
|  | Minimum node size | tuned, 10 to 4000 | tuned, 10 to 4000 |
|  | Number of variables | tuned, 15 to 120 | fixed, 58 |
| XGB | Learning rate (eta) | tuned, 0.001 to 0.01 | tuned, 0.001 to 0.01 |
|  | Minimum loss reduction to split (gamma) | tuned, 0.5 to 5 | tuned, 0.5 to 5 |
|  | Maximum tree depth (max_depth) | tuned, 1 to 18 | tuned, 1 to 18 |
|  | Minimum node size (min_child_weight) | tuned, 60 to 200 | tuned, 60 to 200 |
|  | Subsample ratio (subsample) | tuned, 0.3 to 1 | tuned, 0.3 to 1 |
|  | Fraction of variables to split at each tree (colsample_bytree) | tuned, 0.2 to 1 | tuned, 0.2 to 1 |
|  | L1 regularization term (alpha) | tuned, 0.5 to 10 | tuned, 0.5 to 10 |
|  | The number of iterations (nrounds) | based on early stopping on a maximum of 50000 iterations; stopping if performance does not increase in 30 iterations | based on early stopping on a maximum of 50000 iterations; stopping if performance does not increase in 30 iterations |

Table 7: Fixed and tuned hyperparameters for the RF and XGB models. Hyperparameters not mentioned in the table are left on their default value. Whenever a hyperparameter is tuned, we report the lower and upper bound values provided to the model-based optimization tuning procedure.

#### Superlearner (SL) model

To build the SL model, we fitted the five individual models 10 times using the outer CV folds (Figure 7), making each time predictions on the held-out fold. The CV folds were created based on the admission identifier, such that an admission (with all catheter episodes and landmarks) falls completely into one fold. We imputed the missing data and learn imputation models using missForestPredict on the folds that constitute the training data (9 folds) and applied the imputation models on the fold that constitutes the validation data. The held-out predictions were then “pooled” together and used to train the SL model.^34^. Non-negative least squares (NNLS) meta-learner was used.

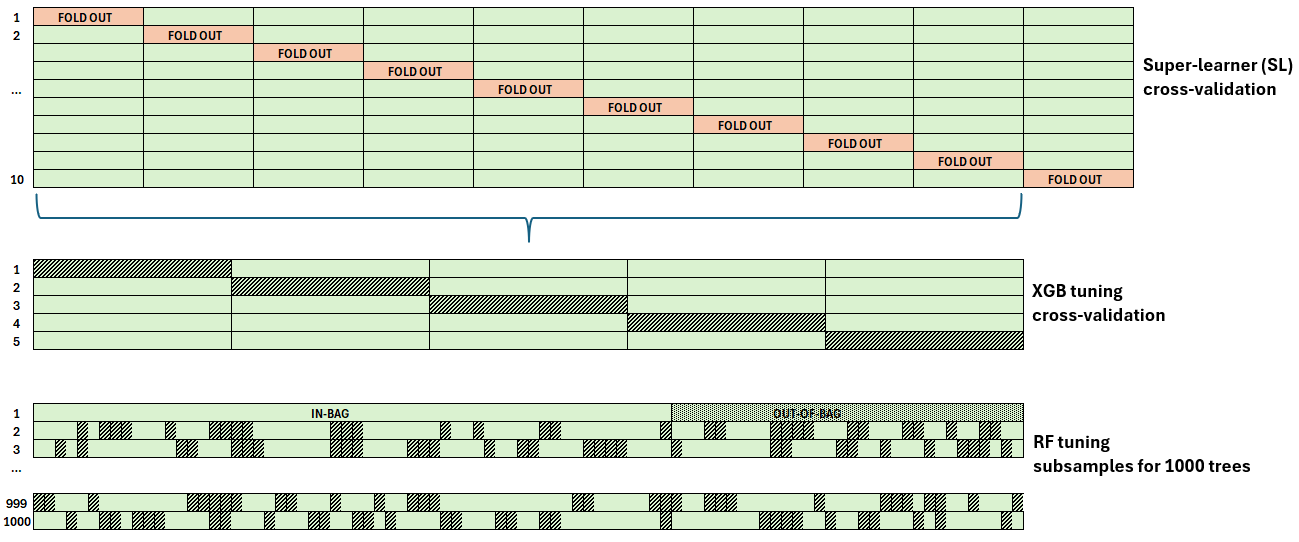

Figure 7: Nested cross-validation for tuning the Superlearner and the RF and XGB models

#### Evaluation metrics

| Metric | Explanation | Type |
| --- | --- | --- |
| AUPRC (Area Under the Precision Recall Curve) | Summary metric for the Precision-Recall Curve. Approximately, the average precision (positive predictive value) over a set of recall (sensitivity) values or over all possible thresholds. | Discrimination |
| AUROC (Area Under the ROC curve) | Summary metric for the Receiver Operating Characteristic (ROC) curve. Given a randomly chosen landmark with and without CLABSI (within 7 days), the probability that the model predictions are higher for the CLABSI landmark. | Discrimination |
| BSS (Brier Skill Score) | The percentage reduction in Brier Score compared to a null model that predicts the event prevalence for all catheter-days. Has a maximum value of 1. The higher the Brier Skill Score is, the better the predictions are discriminated and calibrated^35^. Negative values indicate a model worse than the null model. | Overall performance |
| E:O ratio (Expected-Observed ratio) | The mean of predicted risks divided by the proportion of landmarks with CLABSI (within 7 days). Target value is 1. The E:O ratio > 1 suggests that the model tends to overestimate the risk. | Calibration |
| Calibration slope | Calculated by regressing the true binary outcome on the logit of the predicted risks^36^. Target value is 1. A slope < 1 suggests that estimated risks are too extreme, i.e., too high for patients who are at high-risk and too low for patients who are at low risk. A slope > 1 suggests the opposite, i.e., that risk estimates are too moderate. | Calibration |
| ECI (Estimated Calibration Index) | The mean squared difference between the predicted probabilities and the predicted probabilities obtained with a loess fit of the observed outcome on the predicted risks, multiplied by 100^37^. Target value is 0. The larger the ECI, the more miscalibrated the model. | Calibration |
| Sensitivity | True positive rate; the proportion of landmarks labelled as positive-class (CLABSI within 7 days) that the model correctly identifies as positive. | Threshold dependent |
| Specificity | True negative rate; the proportion of landmarks labelled as negative-class (no CLABSI within 7 days) that the model correctly identifies as negative. | Threshold dependent |
| PPV (Positive Predictive Value) | The proportion of positive predictions (CLABSI within 7 days) that are truly positive (or the proportion of correct alerts). | Threshold dependent |
| NPV (Negative Predictive Value) | The proportion of negative predictions (no CLABSI within 7 days) that are truly negative. | Threshold dependent |
| NB (Net Benefit) | A weighted combination of true and false positives, where the weight reflects the harm of a false positive and the benefit of a true positive (compared to a false negative).^19^ | Threshold dependent |
| Alert rate | The proportion of predictions higher than the evaluation threshold. | Threshold dependent |

Table 8: Metrics evaluated

### Supplementary material 6 - Outcome

Figure 8 presents the cumulative incidence function for all events (CLABSI, death and discharge) in the training and test datasets and the 7 days event proportions over time for the first 20 landmarks in the training and test sets (after landmark 20 there are less than 100 events per landmark in the test set).

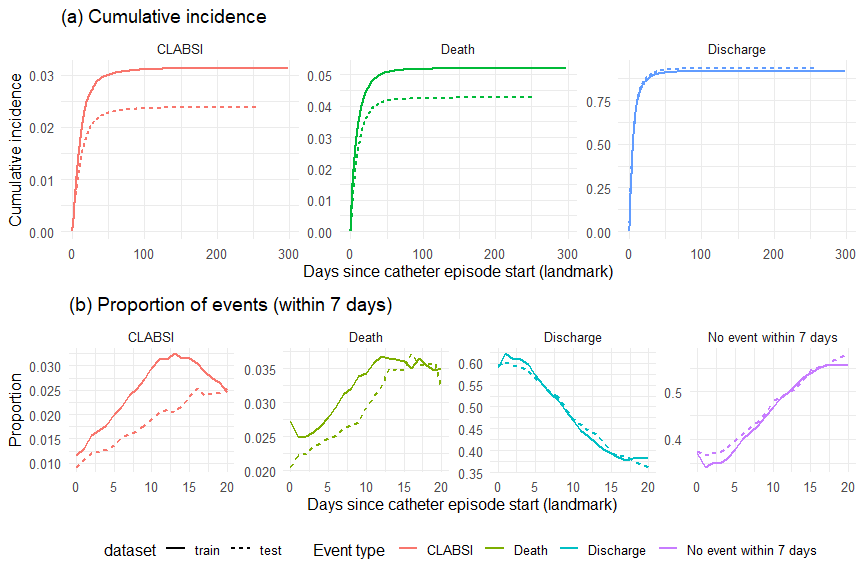

Figure 8: CLABSI, death and discharge events incidence and prevalence (a) Cumulative incidence function curves for all events (training and test datasets) over the followup period (maximum catheter episode length, which is 298 days on the training set). (b) Proportion of events (within 7 days) over time (landmark) for all event types (limited to the first 20 landmarks).

### Supplementary material 7 - Additional evaluation

#### Number and proportion of CLABSI events per landmark

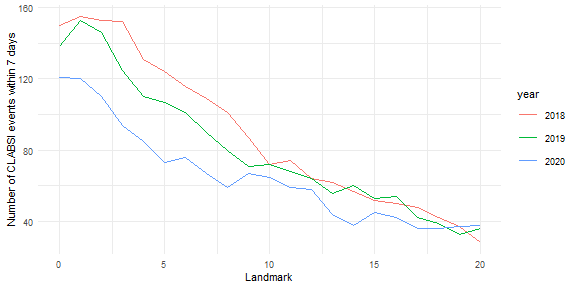

Figure 9: Number of CLABSI events within 7 days per year and landmark in th test set

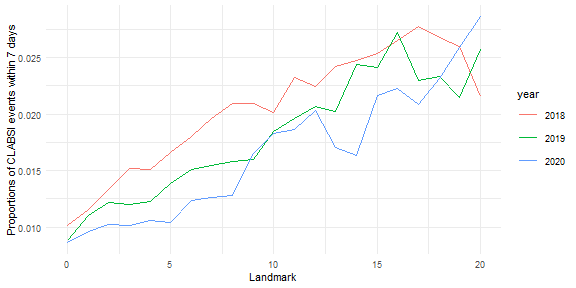

Figure 10: Proportion of CLABSI events within 7 days per year and landmark in the test set

#### Internal evaluation

The peak discrimination performance for all individual models occurred at landmarks 1 to 3 for median AUROC (e.g.: 0.770 to 0.774 for the XGB-ALL model) and at landmarks 11 to 13 for AUPRC and BSS (e.g.: median BSS between 0.026 and 0.030 for the XGB-ALL model). The XGB and cause-specific models produced E:O ratios and calibration slopes close to one, while RF models deviated from one. The ECI deteriorated with landmark for all models: the cause-specific and the XGB-LIM models displayed the similar curves, the XGB-ALL model had the highest ECI (0.053 at landmark 16) and RF models showed slightly less deterioration. The ECI reflected the miscalibration of high predicted risks: XBG and cause-specific models produced high predictions (on cross-validation on all ten folds, the number CLABSI risks higher than 20% were: 490 for cause-specific, 676 for XGB-ALL and 348 for XGB-LIM), while RF models produced lower predictions (54 CLABSI risks higher than 20% for RF-LIM and 1 for RF-ALL).

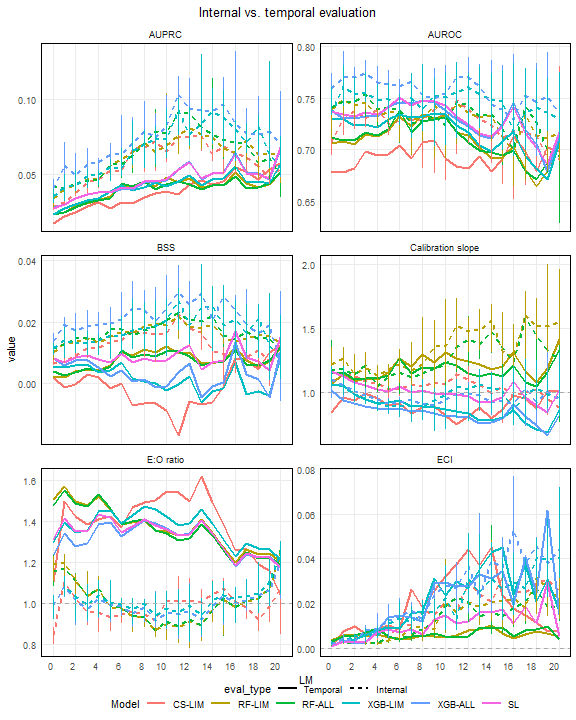

Figure 11: Prediction performance at each landmark on the test set for internal and temporal evaluation. The internal performance of different models is compared with the temporal performance evaluated on the entire test set. AUPRC = Area Under the Precision Recall Curve; AUROC = Area Under the ROC curve; BSS = Brier Skill Score; E:O = Expected:Observed; ECI = Estimated Calibration Index. Median The bars represent the Q1 to Q3 range based on the cross-validation metrics (internal evaluation only).

#### Internal vs. temporal evaluation per year

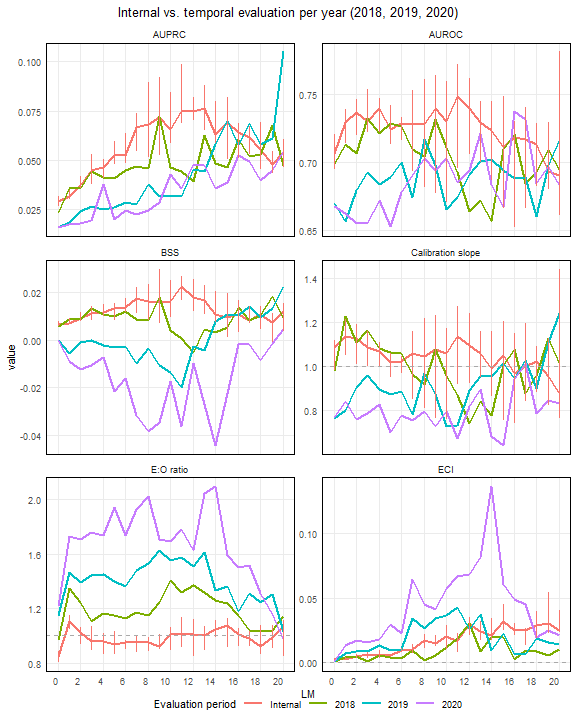

Figure 12: Prediction performance at each landmark on the test set for internal and temporal evaluation for the cause-specific (CS) model. The internal performance is compared to the temporal performance evaluated on each year of the test set. AUPRC = Area Under the Precision Recall Curve; AUROC = Area Under the ROC curve; BSS = Brier Skill Score; E:O = Expected:Observed; ECI = Estimated Calibration Index

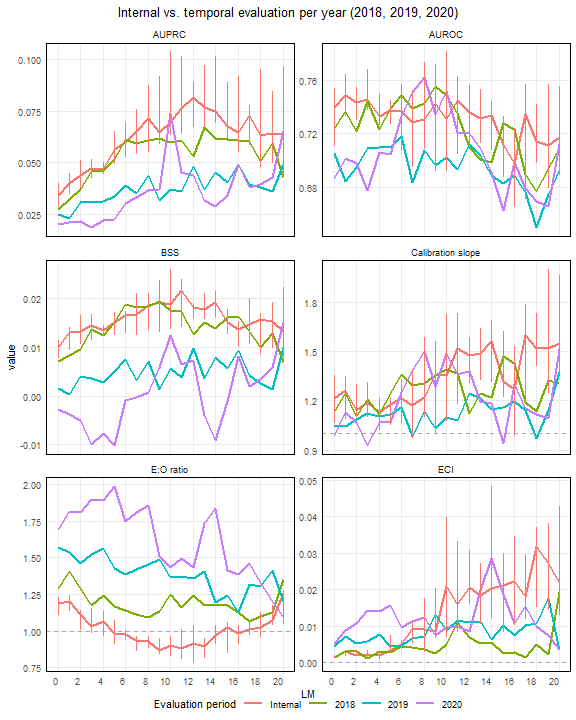

Figure 13: Prediction performance at each landmark on the test set for internal and temporal evaluation for the RF model built on the limited set (RF-LIM). The internal performance is compared to the temporal performance evaluated on each year of the test set. AUPRC = Area Under the Precision Recall Curve; AUROC = Area Under the ROC curve; BSS = Brier Skill Score; E:O = Expected:Observed; ECI = Estimated Calibration Index

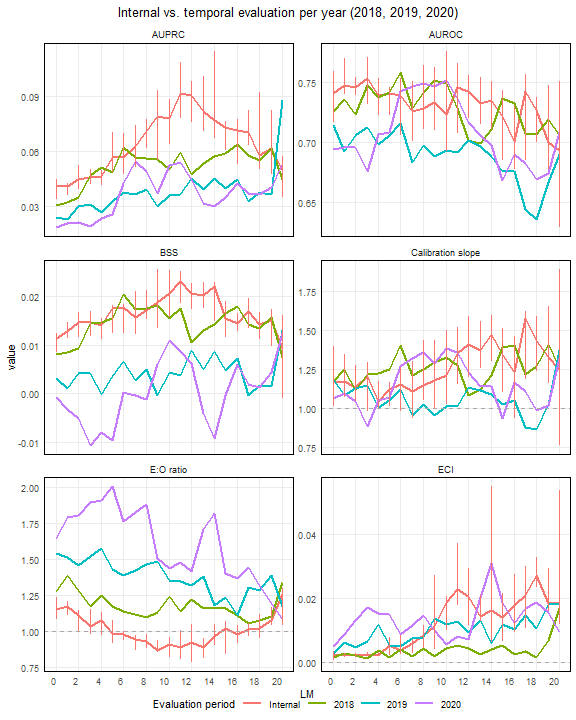

Figure 14: Prediction performance at each landmark for internal and temporal evaluation per year for the RF model built on the complete variables set (RF-ALL). The internal performance is compared to the temporal performance evaluated on each year of the test set. AUPRC = Area Under the Precision Recall Curve; AUROC = Area Under the ROC curve; BSS = Brier Skill Score; E:O = Expected:Observed; ECI = Estimated Calibration Index. The bars represent the Q1 to Q3 range based for the cross-validation metrics (internal evaluation only).

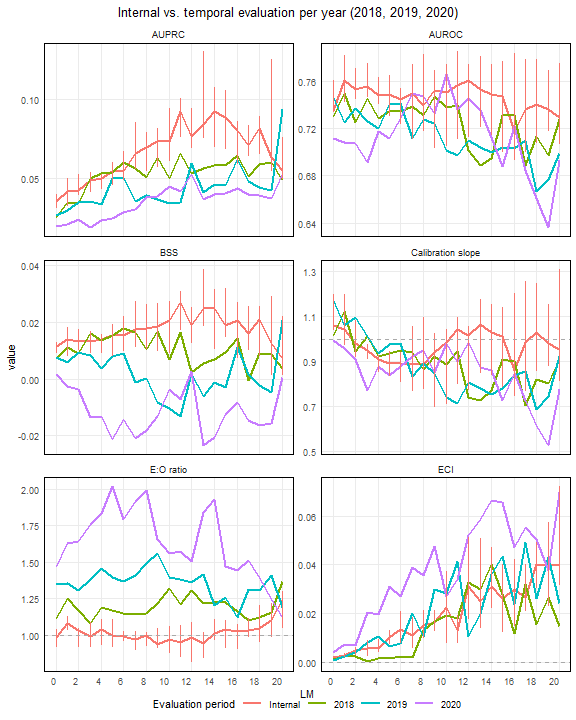

Figure 15: Prediction performance at each landmark on the test set for internal and temporal evaluation for the XGB model built on the limited set (XGB-LIM). The internal performance is compared to the temporal performance evaluated on each year of the test set. AUPRC = Area Under the Precision Recall Curve; AUROC = Area Under the ROC curve; BSS = Brier Skill Score; E:O = Expected:Observed; ECI = Estimated Calibration Index

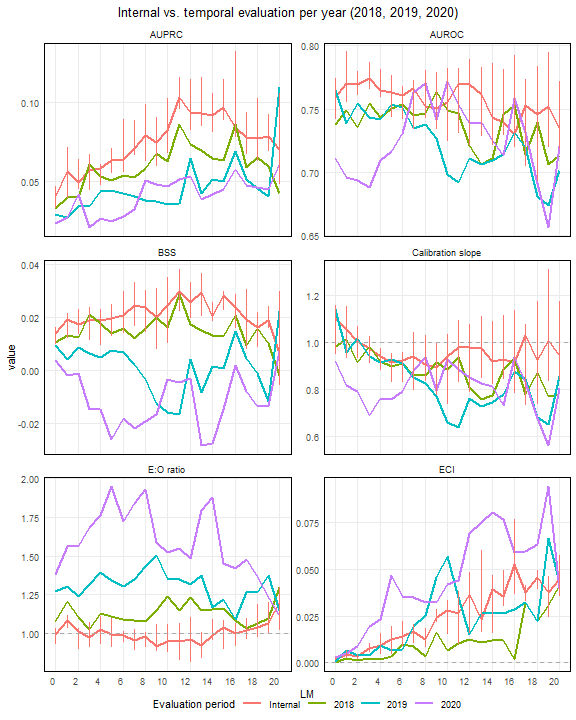

Figure 16: Prediction performance at each landmark on the test set for internal and temporal evaluation for the XGB model built on the complete set (XGB-ALL). The internal performance is compared to the temporal performance evaluated on each year of the test set. AUPRC = Area Under the Precision Recall Curve; AUROC = Area Under the ROC curve; BSS = Brier Skill Score; E:O = Expected:Observed; ECI = Estimated Calibration Index

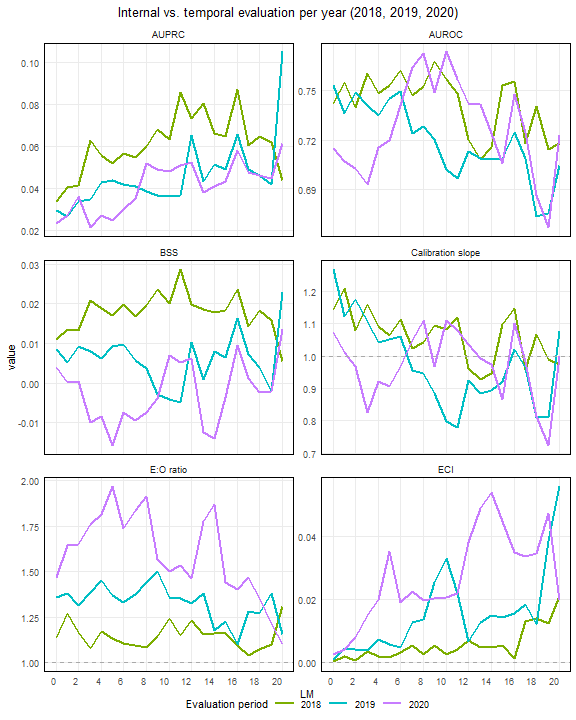

Figure 17: Prediction performance at each landmark on the test set for temporal evaluation for the SL model (superlearner). The temporal performance evaluated on each year of the test set. AUPRC = Area Under the Precision Recall Curve; AUROC = Area Under the ROC curve; BSS = Brier Skill Score; E:O = Expected:Observed; ECI = Estimated Calibration Index

#### Calibration curves per landmark

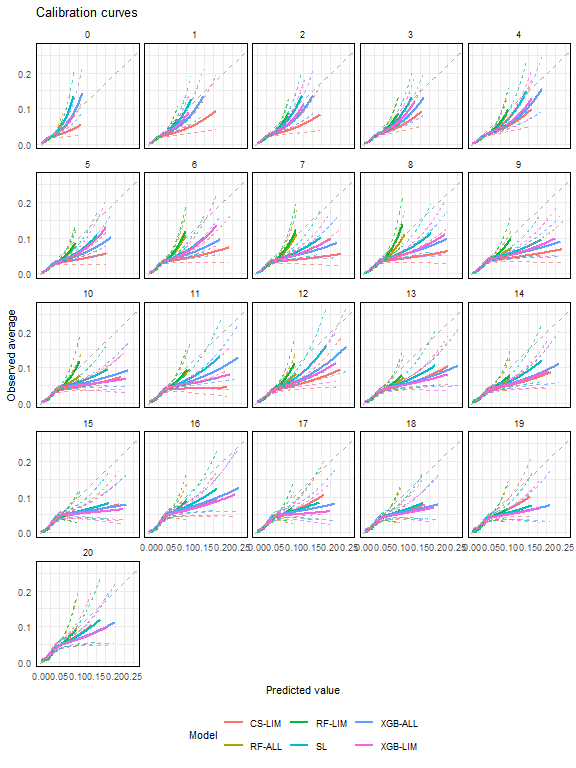

Figure 18: Calibration curves per landmark using restricted cubic splines for each landmark on all predictions in the test set (2018-2020). The dashed line represents the identity function (perfect calibration). The dashed grey line represents the identity function (perfect calibration). The dashed coloured lines represent confidence intervals.

#### Model predictions

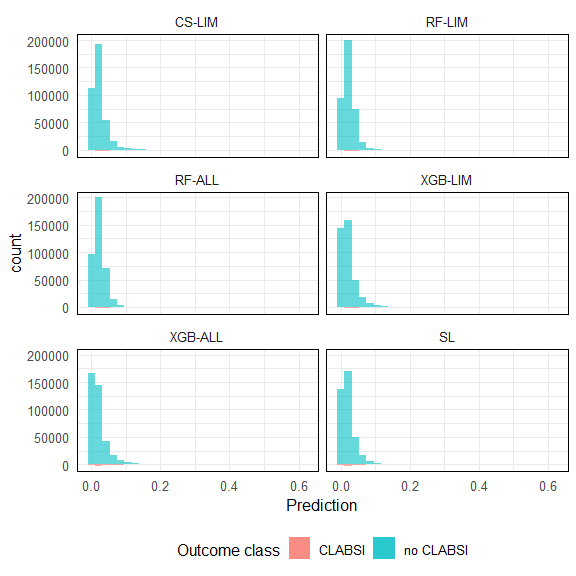

Figure 19: Prediction histograms (counts) for all models (test set) per outcome class (CLABSI vs. no CLABSI). The CLABSI histogram in shown below the x axis.

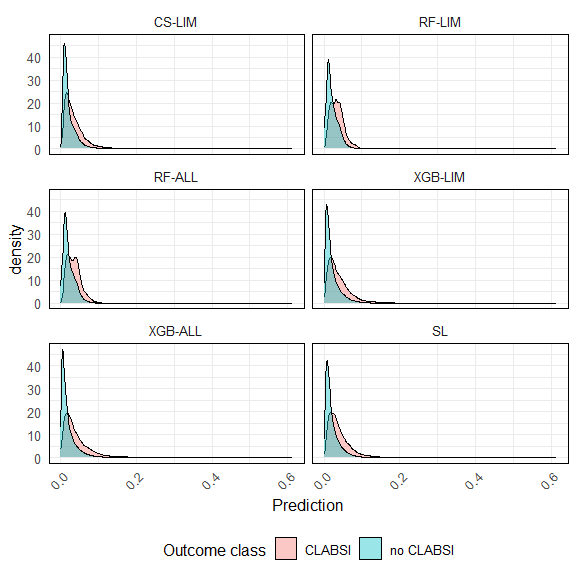

Figure 20: Prediction density plots for all models (test set) per outcome class (CLABSI vs. no CLABSI).

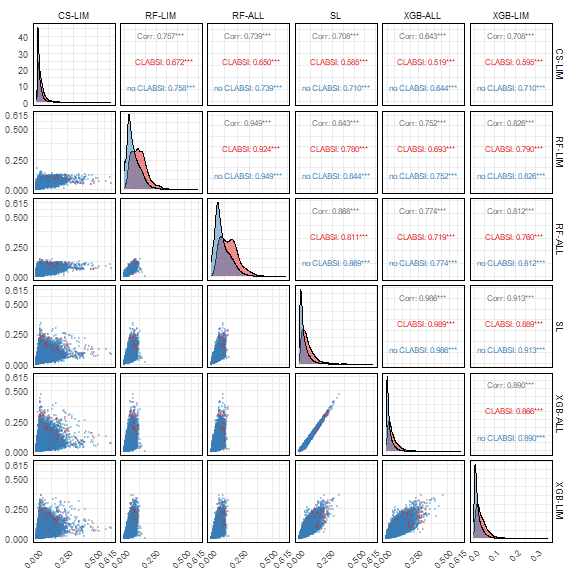

Figure 21: Comparison of predictions by model. The lower plots under the diagonal show the predictions of two models plotted against each other. The diagonal contains the the prediction density curves by class (CLABSI vs. no CLABSI). The upper plots above the diagonal show the correlation (Pearson correlation coefficient) for the predictions of the two models, as well as the correlation within in each class.

#### Threshold independent metric by ward

| Ward | Model | metric | value |
| --- | --- | --- | --- |
| Abdomen | CS-LIM | AUPRC | 0.0269 |
| Abdomen | RF-ALL | AUPRC | 0.0320 |
| Abdomen | RF-LIM | AUPRC | 0.0317 |
| Abdomen | SL | AUPRC | 0.0340 |
| Abdomen | XGB-ALL | AUPRC | 0.0341 |
| Abdomen | XGB-LIM | AUPRC | 0.0329 |
| Cardiac | CS-LIM | AUPRC | 0.0290 |
| Cardiac | RF-ALL | AUPRC | 0.0256 |
| Cardiac | RF-LIM | AUPRC | 0.0193 |
| Cardiac | SL | AUPRC | 0.0275 |
| Cardiac | XGB-ALL | AUPRC | 0.0266 |
| Cardiac | XGB-LIM | AUPRC | 0.0190 |
| Emergency | CS-LIM | AUPRC | 0.0169 |
| Emergency | RF-ALL | AUPRC | 0.0112 |
| Emergency | RF-LIM | AUPRC | 0.0127 |
| Emergency | SL | AUPRC | 0.0174 |
| Emergency | XGB-ALL | AUPRC | 0.0197 |
| Emergency | XGB-LIM | AUPRC | 0.0153 |
| Endocrinology | CS-LIM | AUPRC | 0.0247 |
| Endocrinology | RF-ALL | AUPRC | 0.0242 |
| Endocrinology | RF-LIM | AUPRC | 0.0328 |
| Endocrinology | SL | AUPRC | 0.0282 |
| Endocrinology | XGB-ALL | AUPRC | 0.0295 |
| Endocrinology | XGB-LIM | AUPRC | 0.0339 |
| Geriatrics | CS-LIM | AUPRC | 0.0623 |
| Geriatrics | RF-ALL | AUPRC | 0.1255 |
| Geriatrics | RF-LIM | AUPRC | 0.0897 |
| Geriatrics | SL | AUPRC | 0.0844 |
| Geriatrics | XGB-ALL | AUPRC | 0.0787 |
| Geriatrics | XGB-LIM | AUPRC | 0.0785 |
| Gynecology | CS-LIM | AUPRC | 0.0298 |
| Gynecology | RF-ALL | AUPRC | 0.0404 |
| Gynecology | RF-LIM | AUPRC | 0.0335 |
| Gynecology | SL | AUPRC | 0.0637 |
| Gynecology | XGB-ALL | AUPRC | 0.0615 |
| Gynecology | XGB-LIM | AUPRC | 0.0324 |
| Hematology | CS-LIM | AUPRC | 0.0471 |
| Hematology | RF-ALL | AUPRC | 0.0517 |
| Hematology | RF-LIM | AUPRC | 0.0554 |
| Hematology | SL | AUPRC | 0.0638 |
| Hematology | XGB-ALL | AUPRC | 0.0643 |
| Hematology | XGB-LIM | AUPRC | 0.0555 |
| ICU | CS-LIM | AUPRC | 0.0413 |
| ICU | RF-ALL | AUPRC | 0.0462 |
| ICU | RF-LIM | AUPRC | 0.0456 |
| ICU | SL | AUPRC | 0.0484 |
| ICU | XGB-ALL | AUPRC | 0.0473 |
| ICU | XGB-LIM | AUPRC | 0.0431 |
| Internal Medicine | CS-LIM | AUPRC | 0.0344 |
| Internal Medicine | RF-ALL | AUPRC | 0.0474 |
| Internal Medicine | RF-LIM | AUPRC | 0.0300 |
| Internal Medicine | SL | AUPRC | 0.0428 |
| Internal Medicine | XGB-ALL | AUPRC | 0.0420 |
| Internal Medicine | XGB-LIM | AUPRC | 0.0378 |
| Neonatology | CS-LIM | AUPRC | 0.0317 |
| Neonatology | RF-ALL | AUPRC | 0.0748 |
| Neonatology | RF-LIM | AUPRC | 0.0618 |
| Neonatology | SL | AUPRC | 0.0802 |
| Neonatology | XGB-ALL | AUPRC | 0.0782 |
| Neonatology | XGB-LIM | AUPRC | 0.0661 |
| Nephrology | CS-LIM | AUPRC | 0.0168 |
| Nephrology | RF-ALL | AUPRC | 0.0164 |
| Nephrology | RF-LIM | AUPRC | 0.0148 |
| Nephrology | SL | AUPRC | 0.0152 |
| Nephrology | XGB-ALL | AUPRC | 0.0151 |
| Nephrology | XGB-LIM | AUPRC | 0.0203 |
| Neuro | CS-LIM | AUPRC | 0.0399 |
| Neuro | RF-ALL | AUPRC | 0.0430 |
| Neuro | RF-LIM | AUPRC | 0.0386 |
| Neuro | SL | AUPRC | 0.0739 |
| Neuro | XGB-ALL | AUPRC | 0.0695 |
| Neuro | XGB-LIM | AUPRC | 0.0598 |
| ORL | CS-LIM | AUPRC | 0.0450 |
| ORL | RF-ALL | AUPRC | 0.0361 |
| ORL | RF-LIM | AUPRC | 0.0300 |
| ORL | SL | AUPRC | 0.0442 |
| ORL | XGB-ALL | AUPRC | 0.0446 |
| ORL | XGB-LIM | AUPRC | 0.0407 |
| Oncology | CS-LIM | AUPRC | 0.0316 |
| Oncology | RF-ALL | AUPRC | 0.0329 |
| Oncology | RF-LIM | AUPRC | 0.0321 |
| Oncology | SL | AUPRC | 0.0326 |
| Oncology | XGB-ALL | AUPRC | 0.0309 |
| Oncology | XGB-LIM | AUPRC | 0.0357 |
| Other | CS-LIM | AUPRC | 0.0067 |
| Other | RF-ALL | AUPRC | 0.0161 |
| Other | RF-LIM | AUPRC | 0.0149 |
| Other | SL | AUPRC | 0.0073 |
| Other | XGB-ALL | AUPRC | 0.0054 |
| Other | XGB-LIM | AUPRC | 0.0084 |
| Pediatrics | CS-LIM | AUPRC | 0.0121 |
| Pediatrics | RF-ALL | AUPRC | 0.0143 |
| Pediatrics | RF-LIM | AUPRC | 0.1108 |
| Pediatrics | SL | AUPRC | 0.0156 |
| Pediatrics | XGB-ALL | AUPRC | 0.0144 |
| Pediatrics | XGB-LIM | AUPRC | 0.0109 |
| Pneumology | CS-LIM | AUPRC | 0.0176 |
| Pneumology | RF-ALL | AUPRC | 0.0208 |
| Pneumology | RF-LIM | AUPRC | 0.0176 |
| Pneumology | SL | AUPRC | 0.0199 |
| Pneumology | XGB-ALL | AUPRC | 0.0195 |
| Pneumology | XGB-LIM | AUPRC | 0.0170 |
| Thoracic Surgery | CS-LIM | AUPRC | 0.0349 |
| Thoracic Surgery | RF-ALL | AUPRC | 0.0483 |
| Thoracic Surgery | RF-LIM | AUPRC | 0.0453 |
| Thoracic Surgery | SL | AUPRC | 0.0535 |
| Thoracic Surgery | XGB-ALL | AUPRC | 0.0524 |
| Thoracic Surgery | XGB-LIM | AUPRC | 0.0435 |
| Transplant | CS-LIM | AUPRC | 0.0429 |
| Transplant | RF-ALL | AUPRC | 0.0249 |
| Transplant | RF-LIM | AUPRC | 0.0227 |
| Transplant | SL | AUPRC | 0.0343 |
| Transplant | XGB-ALL | AUPRC | 0.0353 |
| Transplant | XGB-LIM | AUPRC | 0.0301 |
| Traumatology | CS-LIM | AUPRC | 0.0288 |
| Traumatology | RF-ALL | AUPRC | 0.0320 |
| Traumatology | RF-LIM | AUPRC | 0.0269 |
| Traumatology | SL | AUPRC | 0.0379 |
| Traumatology | XGB-ALL | AUPRC | 0.0331 |
| Traumatology | XGB-LIM | AUPRC | 0.0260 |
| UNKNOWN_VALUE | CS-LIM | AUPRC | 0.0185 |
| UNKNOWN_VALUE | RF-ALL | AUPRC | 0.0235 |
| UNKNOWN_VALUE | RF-LIM | AUPRC | 0.0246 |
| UNKNOWN_VALUE | SL | AUPRC | 0.0317 |
| UNKNOWN_VALUE | XGB-ALL | AUPRC | 0.0319 |
| UNKNOWN_VALUE | XGB-LIM | AUPRC | 0.0303 |
| Urology | CS-LIM | AUPRC | 0.0573 |
| Urology | RF-ALL | AUPRC | 0.0755 |
| Urology | RF-LIM | AUPRC | 0.0648 |
| Urology | SL | AUPRC | 0.0649 |
| Urology | XGB-ALL | AUPRC | 0.0596 |
| Urology | XGB-LIM | AUPRC | 0.0623 |
| NA | CS-LIM | AUPRC | NA |
| NA | RF-ALL | AUPRC | NA |
| NA | RF-LIM | AUPRC | NA |
| NA | SL | AUPRC | NA |
| NA | XGB-ALL | AUPRC | NA |
| NA | XGB-LIM | AUPRC | NA |
| Abdomen | CS-LIM | AUROC | 0.6166 |
| Abdomen | RF-ALL | AUROC | 0.6397 |
| Abdomen | RF-LIM | AUROC | 0.6418 |
| Abdomen | SL | AUROC | 0.6686 |
| Abdomen | XGB-ALL | AUROC | 0.6722 |
| Abdomen | XGB-LIM | AUROC | 0.6608 |
| Cardiac | CS-LIM | AUROC | 0.7365 |
| Cardiac | RF-ALL | AUROC | 0.7879 |
| Cardiac | RF-LIM | AUROC | 0.7645 |
| Cardiac | SL | AUROC | 0.8081 |
| Cardiac | XGB-ALL | AUROC | 0.8023 |
| Cardiac | XGB-LIM | AUROC | 0.7773 |
| Emergency | CS-LIM | AUROC | 0.5177 |
| Emergency | RF-ALL | AUROC | 0.5435 |
| Emergency | RF-LIM | AUROC | 0.4610 |
| Emergency | SL | AUROC | 0.6062 |
| Emergency | XGB-ALL | AUROC | 0.6252 |
| Emergency | XGB-LIM | AUROC | 0.6079 |
| Endocrinology | CS-LIM | AUROC | 0.6781 |
| Endocrinology | RF-ALL | AUROC | 0.6723 |
| Endocrinology | RF-LIM | AUROC | 0.7205 |
| Endocrinology | SL | AUROC | 0.7384 |
| Endocrinology | XGB-ALL | AUROC | 0.7486 |
| Endocrinology | XGB-LIM | AUROC | 0.7509 |
| Geriatrics | CS-LIM | AUROC | 0.7036 |
| Geriatrics | RF-ALL | AUROC | 0.7630 |
| Geriatrics | RF-LIM | AUROC | 0.7580 |
| Geriatrics | SL | AUROC | 0.7612 |
| Geriatrics | XGB-ALL | AUROC | 0.7462 |
| Geriatrics | XGB-LIM | AUROC | 0.7589 |
| Gynecology | CS-LIM | AUROC | 0.7511 |
| Gynecology | RF-ALL | AUROC | 0.8331 |
| Gynecology | RF-LIM | AUROC | 0.8212 |
| Gynecology | SL | AUROC | 0.8453 |
| Gynecology | XGB-ALL | AUROC | 0.8448 |
| Gynecology | XGB-LIM | AUROC | 0.8348 |
| Hematology | CS-LIM | AUROC | 0.6165 |
| Hematology | RF-ALL | AUROC | 0.6626 |
| Hematology | RF-LIM | AUROC | 0.6671 |
| Hematology | SL | AUROC | 0.6697 |
| Hematology | XGB-ALL | AUROC | 0.6626 |
| Hematology | XGB-LIM | AUROC | 0.6482 |
| ICU | CS-LIM | AUROC | 0.6433 |
| ICU | RF-ALL | AUROC | 0.6669 |
| ICU | RF-LIM | AUROC | 0.6607 |
| ICU | SL | AUROC | 0.6864 |
| ICU | XGB-ALL | AUROC | 0.6822 |
| ICU | XGB-LIM | AUROC | 0.6667 |
| Internal Medicine | CS-LIM | AUROC | 0.6519 |
| Internal Medicine | RF-ALL | AUROC | 0.6243 |
| Internal Medicine | RF-LIM | AUROC | 0.6355 |
| Internal Medicine | SL | AUROC | 0.6570 |
| Internal Medicine | XGB-ALL | AUROC | 0.6566 |
| Internal Medicine | XGB-LIM | AUROC | 0.6353 |
| Neonatology | CS-LIM | AUROC | 0.6271 |
| Neonatology | RF-ALL | AUROC | 0.7359 |
| Neonatology | RF-LIM | AUROC | 0.7248 |
| Neonatology | SL | AUROC | 0.7539 |
| Neonatology | XGB-ALL | AUROC | 0.7539 |
| Neonatology | XGB-LIM | AUROC | 0.7321 |
| Nephrology | CS-LIM | AUROC | 0.6355 |
| Nephrology | RF-ALL | AUROC | 0.6177 |
| Nephrology | RF-LIM | AUROC | 0.6176 |
| Nephrology | SL | AUROC | 0.6100 |
| Nephrology | XGB-ALL | AUROC | 0.6065 |
| Nephrology | XGB-LIM | AUROC | 0.6250 |
| Neuro | CS-LIM | AUROC | 0.7290 |
| Neuro | RF-ALL | AUROC | 0.7641 |
| Neuro | RF-LIM | AUROC | 0.7442 |
| Neuro | SL | AUROC | 0.7687 |
| Neuro | XGB-ALL | AUROC | 0.7618 |
| Neuro | XGB-LIM | AUROC | 0.7418 |
| ORL | CS-LIM | AUROC | 0.7760 |
| ORL | RF-ALL | AUROC | 0.7756 |
| ORL | RF-LIM | AUROC | 0.7741 |
| ORL | SL | AUROC | 0.8163 |
| ORL | XGB-ALL | AUROC | 0.8103 |
| ORL | XGB-LIM | AUROC | 0.7974 |
| Oncology | CS-LIM | AUROC | 0.7100 |
| Oncology | RF-ALL | AUROC | 0.7084 |
| Oncology | RF-LIM | AUROC | 0.7234 |
| Oncology | SL | AUROC | 0.7422 |
| Oncology | XGB-ALL | AUROC | 0.7423 |
| Oncology | XGB-LIM | AUROC | 0.7504 |
| Other | CS-LIM | AUROC | 0.4957 |
| Other | RF-ALL | AUROC | 0.7704 |
| Other | RF-LIM | AUROC | 0.7644 |
| Other | SL | AUROC | 0.5359 |
| Other | XGB-ALL | AUROC | 0.6255 |
| Other | XGB-LIM | AUROC | 0.5863 |
| Pediatrics | CS-LIM | AUROC | 0.7385 |
| Pediatrics | RF-ALL | AUROC | 0.7610 |
| Pediatrics | RF-LIM | AUROC | 0.7828 |
| Pediatrics | SL | AUROC | 0.7807 |
| Pediatrics | XGB-ALL | AUROC | 0.7702 |
| Pediatrics | XGB-LIM | AUROC | 0.7444 |
| Pneumology | CS-LIM | AUROC | 0.6233 |
| Pneumology | RF-ALL | AUROC | 0.6842 |
| Pneumology | RF-LIM | AUROC | 0.6846 |
| Pneumology | SL | AUROC | 0.7021 |
| Pneumology | XGB-ALL | AUROC | 0.7025 |
| Pneumology | XGB-LIM | AUROC | 0.7137 |
| Thoracic Surgery | CS-LIM | AUROC | 0.7845 |
| Thoracic Surgery | RF-ALL | AUROC | 0.8142 |
| Thoracic Surgery | RF-LIM | AUROC | 0.7903 |
| Thoracic Surgery | SL | AUROC | 0.8221 |
| Thoracic Surgery | XGB-ALL | AUROC | 0.8179 |
| Thoracic Surgery | XGB-LIM | AUROC | 0.7894 |
| Transplant | CS-LIM | AUROC | 0.6775 |
| Transplant | RF-ALL | AUROC | 0.7793 |
| Transplant | RF-LIM | AUROC | 0.7546 |
| Transplant | SL | AUROC | 0.7998 |
| Transplant | XGB-ALL | AUROC | 0.8090 |
| Transplant | XGB-LIM | AUROC | 0.8039 |
| Traumatology | CS-LIM | AUROC | 0.6466 |
| Traumatology | RF-ALL | AUROC | 0.7212 |
| Traumatology | RF-LIM | AUROC | 0.7010 |
| Traumatology | SL | AUROC | 0.7349 |
| Traumatology | XGB-ALL | AUROC | 0.7141 |
| Traumatology | XGB-LIM | AUROC | 0.7117 |
| UNKNOWN_VALUE | CS-LIM | AUROC | 0.7020 |
| UNKNOWN_VALUE | RF-ALL | AUROC | 0.7526 |
| UNKNOWN_VALUE | RF-LIM | AUROC | 0.7583 |
| UNKNOWN_VALUE | SL | AUROC | 0.8068 |
| UNKNOWN_VALUE | XGB-ALL | AUROC | 0.8095 |
| UNKNOWN_VALUE | XGB-LIM | AUROC | 0.7968 |
| Urology | CS-LIM | AUROC | 0.7521 |
| Urology | RF-ALL | AUROC | 0.7420 |
| Urology | RF-LIM | AUROC | 0.7484 |
| Urology | SL | AUROC | 0.7575 |
| Urology | XGB-ALL | AUROC | 0.7482 |
| Urology | XGB-LIM | AUROC | 0.7647 |
| NA | CS-LIM | AUROC | NA |
| NA | RF-ALL | AUROC | NA |
| NA | RF-LIM | AUROC | NA |
| NA | SL | AUROC | NA |
| NA | XGB-ALL | AUROC | NA |
| NA | XGB-LIM | AUROC | NA |
| Abdomen | CS-LIM | BSS | -0.0046 |
| Abdomen | RF-ALL | BSS | 0.0029 |
| Abdomen | RF-LIM | BSS | 0.0023 |
| Abdomen | SL | BSS | 0.0016 |
| Abdomen | XGB-ALL | BSS | -0.0033 |
| Abdomen | XGB-LIM | BSS | -0.0007 |
| Cardiac | CS-LIM | BSS | 0.0080 |
| Cardiac | RF-ALL | BSS | 0.0066 |
| Cardiac | RF-LIM | BSS | 0.0013 |
| Cardiac | SL | BSS | 0.0091 |
| Cardiac | XGB-ALL | BSS | 0.0077 |
| Cardiac | XGB-LIM | BSS | 0.0044 |
| Emergency | CS-LIM | BSS | -0.0055 |
| Emergency | RF-ALL | BSS | -0.0094 |
| Emergency | RF-LIM | BSS | -0.0095 |
| Emergency | SL | BSS | -0.0038 |
| Emergency | XGB-ALL | BSS | -0.0036 |
| Emergency | XGB-LIM | BSS | -0.0068 |
| Endocrinology | CS-LIM | BSS | 0.0015 |
| Endocrinology | RF-ALL | BSS | 0.0031 |
| Endocrinology | RF-LIM | BSS | 0.0077 |
| Endocrinology | SL | BSS | 0.0060 |
| Endocrinology | XGB-ALL | BSS | 0.0041 |
| Endocrinology | XGB-LIM | BSS | 0.0075 |
| Geriatrics | CS-LIM | BSS | 0.0170 |
| Geriatrics | RF-ALL | BSS | 0.0160 |
| Geriatrics | RF-LIM | BSS | 0.0158 |
| Geriatrics | SL | BSS | 0.0258 |
| Geriatrics | XGB-ALL | BSS | 0.0273 |
| Geriatrics | XGB-LIM | BSS | 0.0235 |
| Gynecology | CS-LIM | BSS | 0.0030 |
| Gynecology | RF-ALL | BSS | 0.0097 |
| Gynecology | RF-LIM | BSS | 0.0038 |
| Gynecology | SL | BSS | 0.0178 |
| Gynecology | XGB-ALL | BSS | 0.0203 |
| Gynecology | XGB-LIM | BSS | 0.0111 |
| Hematology | CS-LIM | BSS | -0.0027 |
| Hematology | RF-ALL | BSS | 0.0089 |
| Hematology | RF-LIM | BSS | 0.0099 |
| Hematology | SL | BSS | 0.0124 |
| Hematology | XGB-ALL | BSS | 0.0088 |
| Hematology | XGB-LIM | BSS | 0.0043 |
| ICU | CS-LIM | BSS | -0.0160 |
| ICU | RF-ALL | BSS | -0.0030 |
| ICU | RF-LIM | BSS | 0.0006 |
| ICU | SL | BSS | -0.0051 |
| ICU | XGB-ALL | BSS | -0.0167 |
| ICU | XGB-LIM | BSS | -0.0170 |
| Internal Medicine | CS-LIM | BSS | 0.0030 |
| Internal Medicine | RF-ALL | BSS | 0.0052 |
| Internal Medicine | RF-LIM | BSS | 0.0033 |
| Internal Medicine | SL | BSS | 0.0075 |
| Internal Medicine | XGB-ALL | BSS | 0.0024 |
| Internal Medicine | XGB-LIM | BSS | 0.0058 |
| Neonatology | CS-LIM | BSS | -0.0170 |
| Neonatology | RF-ALL | BSS | 0.0141 |
| Neonatology | RF-LIM | BSS | 0.0100 |
| Neonatology | SL | BSS | 0.0244 |
| Neonatology | XGB-ALL | BSS | 0.0258 |
| Neonatology | XGB-LIM | BSS | 0.0175 |
| Nephrology | CS-LIM | BSS | -0.0134 |
| Nephrology | RF-ALL | BSS | -0.0020 |
| Nephrology | RF-LIM | BSS | -0.0071 |
| Nephrology | SL | BSS | -0.0086 |
| Nephrology | XGB-ALL | BSS | -0.0151 |
| Nephrology | XGB-LIM | BSS | -0.0089 |
| Neuro | CS-LIM | BSS | 0.0099 |
| Neuro | RF-ALL | BSS | 0.0091 |
| Neuro | RF-LIM | BSS | 0.0097 |
| Neuro | SL | BSS | 0.0136 |
| Neuro | XGB-ALL | BSS | 0.0149 |
| Neuro | XGB-LIM | BSS | 0.0149 |
| ORL | CS-LIM | BSS | 0.0150 |
| ORL | RF-ALL | BSS | 0.0131 |
| ORL | RF-LIM | BSS | 0.0106 |
| ORL | SL | BSS | 0.0161 |
| ORL | XGB-ALL | BSS | 0.0153 |
| ORL | XGB-LIM | BSS | 0.0152 |
| Oncology | CS-LIM | BSS | 0.0072 |
| Oncology | RF-ALL | BSS | 0.0089 |
| Oncology | RF-LIM | BSS | 0.0089 |
| Oncology | SL | BSS | 0.0098 |
| Oncology | XGB-ALL | BSS | 0.0078 |
| Oncology | XGB-LIM | BSS | 0.0112 |
| Other | CS-LIM | BSS | -0.0190 |
| Other | RF-ALL | BSS | -0.0069 |
| Other | RF-LIM | BSS | -0.0084 |
| Other | SL | BSS | -0.0177 |
| Other | XGB-ALL | BSS | -0.0259 |
| Other | XGB-LIM | BSS | -0.0318 |
| Pediatrics | CS-LIM | BSS | -0.0080 |
| Pediatrics | RF-ALL | BSS | -0.0103 |
| Pediatrics | RF-LIM | BSS | -0.0117 |
| Pediatrics | SL | BSS | -0.0108 |
| Pediatrics | XGB-ALL | BSS | -0.0158 |
| Pediatrics | XGB-LIM | BSS | -0.0230 |
| Pneumology | CS-LIM | BSS | -0.0071 |
| Pneumology | RF-ALL | BSS | -0.0012 |
| Pneumology | RF-LIM | BSS | -0.0038 |
| Pneumology | SL | BSS | 0.0014 |
| Pneumology | XGB-ALL | BSS | -0.0005 |
| Pneumology | XGB-LIM | BSS | -0.0081 |
| Thoracic Surgery | CS-LIM | BSS | 0.0127 |
| Thoracic Surgery | RF-ALL | BSS | 0.0168 |
| Thoracic Surgery | RF-LIM | BSS | 0.0151 |
| Thoracic Surgery | SL | BSS | 0.0212 |
| Thoracic Surgery | XGB-ALL | BSS | 0.0215 |
| Thoracic Surgery | XGB-LIM | BSS | 0.0169 |
| Transplant | CS-LIM | BSS | 0.0011 |
| Transplant | RF-ALL | BSS | 0.0056 |
| Transplant | RF-LIM | BSS | -0.0009 |
| Transplant | SL | BSS | 0.0099 |
| Transplant | XGB-ALL | BSS | 0.0096 |
| Transplant | XGB-LIM | BSS | 0.0059 |
| Traumatology | CS-LIM | BSS | 0.0076 |
| Traumatology | RF-ALL | BSS | 0.0041 |
| Traumatology | RF-LIM | BSS | 0.0040 |
| Traumatology | SL | BSS | 0.0092 |
| Traumatology | XGB-ALL | BSS | 0.0092 |
| Traumatology | XGB-LIM | BSS | 0.0068 |
| UNKNOWN_VALUE | CS-LIM | BSS | -0.0072 |
| UNKNOWN_VALUE | RF-ALL | BSS | -0.0029 |
| UNKNOWN_VALUE | RF-LIM | BSS | -0.0129 |
| UNKNOWN_VALUE | SL | BSS | 0.0055 |
| UNKNOWN_VALUE | XGB-ALL | BSS | 0.0072 |
| UNKNOWN_VALUE | XGB-LIM | BSS | 0.0027 |
| Urology | CS-LIM | BSS | 0.0164 |
| Urology | RF-ALL | BSS | 0.0174 |
| Urology | RF-LIM | BSS | 0.0190 |
| Urology | SL | BSS | 0.0200 |
| Urology | XGB-ALL | BSS | 0.0191 |
| Urology | XGB-LIM | BSS | 0.0209 |
| NA | CS-LIM | BSS | NA |
| NA | RF-ALL | BSS | NA |
| NA | RF-LIM | BSS | NA |
| NA | SL | BSS | NA |
| NA | XGB-ALL | BSS | NA |
| NA | XGB-LIM | BSS | NA |
| Abdomen | CS-LIM | Calibration slope | 0.5290 |
| Abdomen | RF-ALL | Calibration slope | 0.8750 |
| Abdomen | RF-LIM | Calibration slope | 0.7960 |
| Abdomen | SL | Calibration slope | 0.8544 |
| Abdomen | XGB-ALL | Calibration slope | 0.7502 |
| Abdomen | XGB-LIM | Calibration slope | 0.7561 |
| Cardiac | CS-LIM | Calibration slope | 1.4848 |
| Cardiac | RF-ALL | Calibration slope | 1.6648 |
| Cardiac | RF-LIM | Calibration slope | 1.3347 |
| Cardiac | SL | Calibration slope | 1.3531 |
| Cardiac | XGB-ALL | Calibration slope | 1.1119 |
| Cardiac | XGB-LIM | Calibration slope | 1.1279 |
| Emergency | CS-LIM | Calibration slope | 0.2589 |
| Emergency | RF-ALL | Calibration slope | 0.3148 |
| Emergency | RF-LIM | Calibration slope | 0.3084 |
| Emergency | SL | Calibration slope | 0.7531 |
| Emergency | XGB-ALL | Calibration slope | 0.7198 |
| Emergency | XGB-LIM | Calibration slope | 0.5957 |
| Endocrinology | CS-LIM | Calibration slope | 1.0572 |
| Endocrinology | RF-ALL | Calibration slope | 1.3173 |
| Endocrinology | RF-LIM | Calibration slope | 1.6125 |
| Endocrinology | SL | Calibration slope | 1.1528 |
| Endocrinology | XGB-ALL | Calibration slope | 0.9385 |
| Endocrinology | XGB-LIM | Calibration slope | 1.0632 |
| Geriatrics | CS-LIM | Calibration slope | 1.4839 |
| Geriatrics | RF-ALL | Calibration slope | 3.5274 |
| Geriatrics | RF-LIM | Calibration slope | 2.7058 |
| Geriatrics | SL | Calibration slope | 1.5812 |
| Geriatrics | XGB-ALL | Calibration slope | 1.1860 |
| Geriatrics | XGB-LIM | Calibration slope | 1.4434 |
| Gynecology | CS-LIM | Calibration slope | 1.6510 |
| Gynecology | RF-ALL | Calibration slope | 2.2223 |
| Gynecology | RF-LIM | Calibration slope | 2.0912 |
| Gynecology | SL | Calibration slope | 1.9605 |
| Gynecology | XGB-ALL | Calibration slope | 1.6081 |
| Gynecology | XGB-LIM | Calibration slope | 1.6746 |
| Hematology | CS-LIM | Calibration slope | 0.5661 |
| Hematology | RF-ALL | Calibration slope | 1.2653 |
| Hematology | RF-LIM | Calibration slope | 1.1383 |
| Hematology | SL | Calibration slope | 0.9187 |
| Hematology | XGB-ALL | Calibration slope | 0.7037 |
| Hematology | XGB-LIM | Calibration slope | 0.6544 |
| ICU | CS-LIM | Calibration slope | 0.6642 |
| ICU | RF-ALL | Calibration slope | 1.2453 |
| ICU | RF-LIM | Calibration slope | 1.1941 |
| ICU | SL | Calibration slope | 0.9762 |
| ICU | XGB-ALL | Calibration slope | 0.7782 |
| ICU | XGB-LIM | Calibration slope | 0.7903 |
| Internal Medicine | CS-LIM | Calibration slope | 1.0012 |
| Internal Medicine | RF-ALL | Calibration slope | 1.4384 |
| Internal Medicine | RF-LIM | Calibration slope | 1.1375 |
| Internal Medicine | SL | Calibration slope | 1.0015 |
| Internal Medicine | XGB-ALL | Calibration slope | 0.7813 |
| Internal Medicine | XGB-LIM | Calibration slope | 0.8021 |
| Neonatology | CS-LIM | Calibration slope | 0.9984 |
| Neonatology | RF-ALL | Calibration slope | 2.0272 |
| Neonatology | RF-LIM | Calibration slope | 2.0170 |
| Neonatology | SL | Calibration slope | 1.4454 |
| Neonatology | XGB-ALL | Calibration slope | 1.1361 |
| Neonatology | XGB-LIM | Calibration slope | 1.0359 |
| Nephrology | CS-LIM | Calibration slope | 0.6665 |
| Nephrology | RF-ALL | Calibration slope | 0.6842 |
| Nephrology | RF-LIM | Calibration slope | 0.6358 |
| Nephrology | SL | Calibration slope | 0.5000 |
| Nephrology | XGB-ALL | Calibration slope | 0.3946 |
| Nephrology | XGB-LIM | Calibration slope | 0.5521 |
| Neuro | CS-LIM | Calibration slope | 1.1185 |
| Neuro | RF-ALL | Calibration slope | 1.4283 |
| Neuro | RF-LIM | Calibration slope | 1.1841 |
| Neuro | SL | Calibration slope | 1.3449 |
| Neuro | XGB-ALL | Calibration slope | 1.1556 |
| Neuro | XGB-LIM | Calibration slope | 1.0666 |
| ORL | CS-LIM | Calibration slope | 1.5839 |
| ORL | RF-ALL | Calibration slope | 1.6304 |
| ORL | RF-LIM | Calibration slope | 1.5608 |
| ORL | SL | Calibration slope | 1.3997 |
| ORL | XGB-ALL | Calibration slope | 1.1616 |
| ORL | XGB-LIM | Calibration slope | 1.1951 |
| Oncology | CS-LIM | Calibration slope | 1.3011 |
| Oncology | RF-ALL | Calibration slope | 1.3514 |
| Oncology | RF-LIM | Calibration slope | 1.2537 |
| Oncology | SL | Calibration slope | 1.1638 |
| Oncology | XGB-ALL | Calibration slope | 0.9871 |
| Oncology | XGB-LIM | Calibration slope | 1.1592 |
| Other | CS-LIM | Calibration slope | 0.0382 |
| Other | RF-ALL | Calibration slope | 1.8778 |
| Other | RF-LIM | Calibration slope | 1.4833 |
| Other | SL | Calibration slope | 0.1358 |
| Other | XGB-ALL | Calibration slope | -0.5013 |
| Other | XGB-LIM | Calibration slope | 0.2305 |
| Pediatrics | CS-LIM | Calibration slope | 1.0158 |
| Pediatrics | RF-ALL | Calibration slope | 1.3468 |
| Pediatrics | RF-LIM | Calibration slope | 1.6254 |
| Pediatrics | SL | Calibration slope | 1.1590 |
| Pediatrics | XGB-ALL | Calibration slope | 0.9531 |
| Pediatrics | XGB-LIM | Calibration slope | 0.8378 |
| Pneumology | CS-LIM | Calibration slope | 0.8991 |
| Pneumology | RF-ALL | Calibration slope | 1.2746 |
| Pneumology | RF-LIM | Calibration slope | 1.1420 |
| Pneumology | SL | Calibration slope | 0.9693 |
| Pneumology | XGB-ALL | Calibration slope | 0.7682 |
| Pneumology | XGB-LIM | Calibration slope | 0.8322 |
| Thoracic Surgery | CS-LIM | Calibration slope | 1.5132 |
| Thoracic Surgery | RF-ALL | Calibration slope | 2.1124 |
| Thoracic Surgery | RF-LIM | Calibration slope | 1.9545 |
| Thoracic Surgery | SL | Calibration slope | 1.6108 |
| Thoracic Surgery | XGB-ALL | Calibration slope | 1.3425 |
| Thoracic Surgery | XGB-LIM | Calibration slope | 1.3570 |
| Transplant | CS-LIM | Calibration slope | 1.3457 |
| Transplant | RF-ALL | Calibration slope | 1.6938 |
| Transplant | RF-LIM | Calibration slope | 1.5457 |
| Transplant | SL | Calibration slope | 1.4633 |
| Transplant | XGB-ALL | Calibration slope | 1.2706 |
| Transplant | XGB-LIM | Calibration slope | 1.4228 |
| Traumatology | CS-LIM | Calibration slope | 1.4668 |
| Traumatology | RF-ALL | Calibration slope | 1.9871 |
| Traumatology | RF-LIM | Calibration slope | 1.7446 |
| Traumatology | SL | Calibration slope | 1.6168 |
| Traumatology | XGB-ALL | Calibration slope | 1.2208 |
| Traumatology | XGB-LIM | Calibration slope | 1.4353 |
| UNKNOWN_VALUE | CS-LIM | Calibration slope | 1.0508 |
| UNKNOWN_VALUE | RF-ALL | Calibration slope | 2.1035 |
| UNKNOWN_VALUE | RF-LIM | Calibration slope | 2.0674 |
| UNKNOWN_VALUE | SL | Calibration slope | 1.7538 |
| UNKNOWN_VALUE | XGB-ALL | Calibration slope | 1.4493 |
| UNKNOWN_VALUE | XGB-LIM | Calibration slope | 1.4287 |
| Urology | CS-LIM | Calibration slope | 1.4556 |
| Urology | RF-ALL | Calibration slope | 1.7055 |
| Urology | RF-LIM | Calibration slope | 1.6312 |
| Urology | SL | Calibration slope | 1.3894 |
| Urology | XGB-ALL | Calibration slope | 1.1662 |
| Urology | XGB-LIM | Calibration slope | 1.2959 |
| NA | CS-LIM | Calibration slope | NA |
| NA | RF-ALL | Calibration slope | NA |
| NA | RF-LIM | Calibration slope | NA |
| NA | SL | Calibration slope | NA |
| NA | XGB-ALL | Calibration slope | NA |
| NA | XGB-LIM | Calibration slope | NA |
| Abdomen | CS-LIM | E:O ratio | 1.3040 |
| Abdomen | RF-ALL | E:O ratio | 1.2296 |
| Abdomen | RF-LIM | E:O ratio | 1.2452 |
| Abdomen | SL | E:O ratio | 1.3099 |
| Abdomen | XGB-ALL | E:O ratio | 1.3493 |
| Abdomen | XGB-LIM | E:O ratio | 1.3265 |
| Cardiac | CS-LIM | E:O ratio | 1.5351 |
| Cardiac | RF-ALL | E:O ratio | 1.7004 |
| Cardiac | RF-LIM | E:O ratio | 1.9003 |
| Cardiac | SL | E:O ratio | 1.2777 |
| Cardiac | XGB-ALL | E:O ratio | 1.0373 |
| Cardiac | XGB-LIM | E:O ratio | 1.2829 |
| Emergency | CS-LIM | E:O ratio | 1.3234 |
| Emergency | RF-ALL | E:O ratio | 1.6891 |
| Emergency | RF-LIM | E:O ratio | 1.6703 |
| Emergency | SL | E:O ratio | 1.5648 |
| Emergency | XGB-ALL | E:O ratio | 1.5111 |
| Emergency | XGB-LIM | E:O ratio | 1.5994 |
| Endocrinology | CS-LIM | E:O ratio | 1.4102 |
| Endocrinology | RF-ALL | E:O ratio | 1.3656 |
| Endocrinology | RF-LIM | E:O ratio | 1.3448 |
| Endocrinology | SL | E:O ratio | 1.1848 |
| Endocrinology | XGB-ALL | E:O ratio | 1.0882 |
| Endocrinology | XGB-LIM | E:O ratio | 1.1579 |
| Geriatrics | CS-LIM | E:O ratio | 1.0063 |
| Geriatrics | RF-ALL | E:O ratio | 1.0863 |
| Geriatrics | RF-LIM | E:O ratio | 1.0124 |
| Geriatrics | SL | E:O ratio | 0.9438 |
| Geriatrics | XGB-ALL | E:O ratio | 0.8764 |
| Geriatrics | XGB-LIM | E:O ratio | 0.8696 |
| Gynecology | CS-LIM | E:O ratio | 2.0494 |
| Gynecology | RF-ALL | E:O ratio | 1.7682 |
| Gynecology | RF-LIM | E:O ratio | 2.1337 |
| Gynecology | SL | E:O ratio | 1.4985 |
| Gynecology | XGB-ALL | E:O ratio | 1.3113 |
| Gynecology | XGB-LIM | E:O ratio | 1.7095 |
| Hematology | CS-LIM | E:O ratio | 1.1024 |
| Hematology | RF-ALL | E:O ratio | 1.0400 |
| Hematology | RF-LIM | E:O ratio | 1.0141 |
| Hematology | SL | E:O ratio | 1.0676 |
| Hematology | XGB-ALL | E:O ratio | 1.0828 |
| Hematology | XGB-LIM | E:O ratio | 1.0868 |
| ICU | CS-LIM | E:O ratio | 1.5169 |
| ICU | RF-ALL | E:O ratio | 1.6953 |
| ICU | RF-LIM | E:O ratio | 1.5726 |
| ICU | SL | E:O ratio | 1.6718 |
| ICU | XGB-ALL | E:O ratio | 1.6759 |
| ICU | XGB-LIM | E:O ratio | 1.7065 |
| Internal Medicine | CS-LIM | E:O ratio | 1.3809 |
| Internal Medicine | RF-ALL | E:O ratio | 1.3570 |
| Internal Medicine | RF-LIM | E:O ratio | 1.2903 |
| Internal Medicine | SL | E:O ratio | 1.3584 |
| Internal Medicine | XGB-ALL | E:O ratio | 1.3645 |
| Internal Medicine | XGB-LIM | E:O ratio | 1.1929 |
| Neonatology | CS-LIM | E:O ratio | 1.8267 |
| Neonatology | RF-ALL | E:O ratio | 1.4759 |
| Neonatology | RF-LIM | E:O ratio | 1.4695 |
| Neonatology | SL | E:O ratio | 1.3489 |
| Neonatology | XGB-ALL | E:O ratio | 1.2703 |
| Neonatology | XGB-LIM | E:O ratio | 1.3339 |
| Nephrology | CS-LIM | E:O ratio | 1.6838 |
| Nephrology | RF-ALL | E:O ratio | 1.3330 |
| Nephrology | RF-LIM | E:O ratio | 1.5270 |
| Nephrology | SL | E:O ratio | 1.2768 |
| Nephrology | XGB-ALL | E:O ratio | 1.2179 |
| Nephrology | XGB-LIM | E:O ratio | 1.4648 |
| Neuro | CS-LIM | E:O ratio | 0.8100 |
| Neuro | RF-ALL | E:O ratio | 0.6730 |
| Neuro | RF-LIM | E:O ratio | 0.7682 |
| Neuro | SL | E:O ratio | 0.6361 |
| Neuro | XGB-ALL | E:O ratio | 0.6033 |
| Neuro | XGB-LIM | E:O ratio | 0.6947 |
| ORL | CS-LIM | E:O ratio | 1.1563 |
| ORL | RF-ALL | E:O ratio | 1.1621 |
| ORL | RF-LIM | E:O ratio | 1.3088 |
| ORL | SL | E:O ratio | 1.0359 |
| ORL | XGB-ALL | E:O ratio | 0.9548 |
| ORL | XGB-LIM | E:O ratio | 1.1614 |
| Oncology | CS-LIM | E:O ratio | 1.4244 |
| Oncology | RF-ALL | E:O ratio | 1.1175 |
| Oncology | RF-LIM | E:O ratio | 1.2480 |
| Oncology | SL | E:O ratio | 1.1437 |
| Oncology | XGB-ALL | E:O ratio | 1.1363 |
| Oncology | XGB-LIM | E:O ratio | 1.1917 |
| Other | CS-LIM | E:O ratio | 2.0488 |
| Other | RF-ALL | E:O ratio | 2.1770 |
| Other | RF-LIM | E:O ratio | 2.1091 |
| Other | SL | E:O ratio | 1.8320 |
| Other | XGB-ALL | E:O ratio | 1.6568 |
| Other | XGB-LIM | E:O ratio | 1.9802 |
| Pediatrics | CS-LIM | E:O ratio | 2.1388 |
| Pediatrics | RF-ALL | E:O ratio | 2.6319 |
| Pediatrics | RF-LIM | E:O ratio | 3.0167 |
| Pediatrics | SL | E:O ratio | 2.3594 |
| Pediatrics | XGB-ALL | E:O ratio | 2.1931 |
| Pediatrics | XGB-LIM | E:O ratio | 2.3008 |
| Pneumology | CS-LIM | E:O ratio | 1.9705 |
| Pneumology | RF-ALL | E:O ratio | 1.7955 |
| Pneumology | RF-LIM | E:O ratio | 1.8892 |
| Pneumology | SL | E:O ratio | 1.4677 |
| Pneumology | XGB-ALL | E:O ratio | 1.2775 |
| Pneumology | XGB-LIM | E:O ratio | 1.6436 |
| Thoracic Surgery | CS-LIM | E:O ratio | 1.1423 |
| Thoracic Surgery | RF-ALL | E:O ratio | 1.2601 |
| Thoracic Surgery | RF-LIM | E:O ratio | 1.3440 |
| Thoracic Surgery | SL | E:O ratio | 1.1575 |
| Thoracic Surgery | XGB-ALL | E:O ratio | 1.0983 |
| Thoracic Surgery | XGB-LIM | E:O ratio | 1.1649 |
| Transplant | CS-LIM | E:O ratio | 2.0239 |
| Transplant | RF-ALL | E:O ratio | 1.6991 |
| Transplant | RF-LIM | E:O ratio | 2.0117 |
| Transplant | SL | E:O ratio | 1.5296 |
| Transplant | XGB-ALL | E:O ratio | 1.3998 |
| Transplant | XGB-LIM | E:O ratio | 1.7459 |
| Traumatology | CS-LIM | E:O ratio | 1.4638 |
| Traumatology | RF-ALL | E:O ratio | 1.8714 |
| Traumatology | RF-LIM | E:O ratio | 1.7828 |
| Traumatology | SL | E:O ratio | 1.3980 |
| Traumatology | XGB-ALL | E:O ratio | 1.1644 |
| Traumatology | XGB-LIM | E:O ratio | 1.4168 |
| UNKNOWN_VALUE | CS-LIM | E:O ratio | 1.9716 |
| UNKNOWN_VALUE | RF-ALL | E:O ratio | 2.1749 |
| UNKNOWN_VALUE | RF-LIM | E:O ratio | 2.6009 |
| UNKNOWN_VALUE | SL | E:O ratio | 1.9017 |
| UNKNOWN_VALUE | XGB-ALL | E:O ratio | 1.7223 |
| UNKNOWN_VALUE | XGB-LIM | E:O ratio | 1.9405 |
| Urology | CS-LIM | E:O ratio | 0.8033 |
| Urology | RF-ALL | E:O ratio | 0.7562 |
| Urology | RF-LIM | E:O ratio | 0.8404 |
| Urology | SL | E:O ratio | 0.8393 |
| Urology | XGB-ALL | E:O ratio | 0.8749 |
| Urology | XGB-LIM | E:O ratio | 0.8553 |
| NA | CS-LIM | E:O ratio | NA |
| NA | RF-ALL | E:O ratio | NA |
| NA | RF-LIM | E:O ratio | NA |
| NA | SL | E:O ratio | NA |
| NA | XGB-ALL | E:O ratio | NA |
| NA | XGB-LIM | E:O ratio | NA |
| Abdomen | CS-LIM | ECI | 0.0119 |
| Abdomen | RF-ALL | ECI | 0.0038 |
| Abdomen | RF-LIM | ECI | 0.0055 |
| Abdomen | SL | ECI | 0.0118 |
| Abdomen | XGB-ALL | ECI | 0.0216 |
| Abdomen | XGB-LIM | ECI | 0.0114 |
| Cardiac | CS-LIM | ECI | 0.0025 |
| Cardiac | RF-ALL | ECI | 0.0025 |
| Cardiac | RF-LIM | ECI | 0.0041 |
| Cardiac | SL | ECI | 0.0021 |
| Cardiac | XGB-ALL | ECI | 0.0022 |
| Cardiac | XGB-LIM | ECI | 0.0028 |
| Emergency | CS-LIM | ECI | 0.0087 |
| Emergency | RF-ALL | ECI | 0.0100 |
| Emergency | RF-LIM | ECI | 0.0117 |
| Emergency | SL | ECI | 0.0080 |
| Emergency | XGB-ALL | ECI | 0.0120 |
| Emergency | XGB-LIM | ECI | 0.0102 |
| Endocrinology | CS-LIM | ECI | 0.0055 |
| Endocrinology | RF-ALL | ECI | 0.0088 |
| Endocrinology | RF-LIM | ECI | 0.0045 |
| Endocrinology | SL | ECI | 0.0053 |
| Endocrinology | XGB-ALL | ECI | 0.0077 |
| Endocrinology | XGB-LIM | ECI | 0.0075 |
| Geriatrics | CS-LIM | ECI | 0.0083 |
| Geriatrics | RF-ALL | ECI | 0.0818 |
| Geriatrics | RF-LIM | ECI | 0.0372 |
| Geriatrics | SL | ECI | 0.0087 |
| Geriatrics | XGB-ALL | ECI | 0.0076 |
| Geriatrics | XGB-LIM | ECI | 0.0029 |
| Gynecology | CS-LIM | ECI | 0.0050 |
| Gynecology | RF-ALL | ECI | 0.0047 |
| Gynecology | RF-LIM | ECI | 0.0063 |
| Gynecology | SL | ECI | 0.0081 |
| Gynecology | XGB-ALL | ECI | 0.0058 |
| Gynecology | XGB-LIM | ECI | 0.0028 |
| Hematology | CS-LIM | ECI | 0.0282 |
| Hematology | RF-ALL | ECI | 0.0039 |
| Hematology | RF-LIM | ECI | 0.0040 |
| Hematology | SL | ECI | 0.0019 |
| Hematology | XGB-ALL | ECI | 0.0115 |
| Hematology | XGB-LIM | ECI | 0.0166 |
| ICU | CS-LIM | ECI | 0.0468 |
| ICU | RF-ALL | ECI | 0.0270 |
| ICU | RF-LIM | ECI | 0.0198 |
| ICU | SL | ECI | 0.0368 |
| ICU | XGB-ALL | ECI | 0.0625 |
| ICU | XGB-LIM | ECI | 0.0618 |
| Internal Medicine | CS-LIM | ECI | 0.0170 |
| Internal Medicine | RF-ALL | ECI | 0.0160 |
| Internal Medicine | RF-LIM | ECI | 0.0055 |
| Internal Medicine | SL | ECI | 0.0103 |
| Internal Medicine | XGB-ALL | ECI | 0.0189 |
| Internal Medicine | XGB-LIM | ECI | 0.0092 |
| Neonatology | CS-LIM | ECI | 0.0353 |
| Neonatology | RF-ALL | ECI | 0.0237 |
| Neonatology | RF-LIM | ECI | 0.0177 |
| Neonatology | SL | ECI | 0.0082 |
| Neonatology | XGB-ALL | ECI | 0.0054 |
| Neonatology | XGB-LIM | ECI | 0.0107 |
| Nephrology | CS-LIM | ECI | 0.0221 |
| Nephrology | RF-ALL | ECI | 0.0063 |
| Nephrology | RF-LIM | ECI | 0.0108 |
| Nephrology | SL | ECI | 0.0090 |
| Nephrology | XGB-ALL | ECI | 0.0128 |
| Nephrology | XGB-LIM | ECI | 0.0175 |
| Neuro | CS-LIM | ECI | 0.0033 |
| Neuro | RF-ALL | ECI | 0.0082 |
| Neuro | RF-LIM | ECI | 0.0035 |
| Neuro | SL | ECI | 0.0350 |
| Neuro | XGB-ALL | ECI | 0.0422 |
| Neuro | XGB-LIM | ECI | 0.0091 |
| ORL | CS-LIM | ECI | 0.0059 |
| ORL | RF-ALL | ECI | 0.0027 |
| ORL | RF-LIM | ECI | 0.0037 |
| ORL | SL | ECI | 0.0053 |
| ORL | XGB-ALL | ECI | 0.0067 |
| ORL | XGB-LIM | ECI | 0.0025 |
| Oncology | CS-LIM | ECI | 0.0055 |
| Oncology | RF-ALL | ECI | 0.0015 |
| Oncology | RF-LIM | ECI | 0.0014 |
| Oncology | SL | ECI | 0.0007 |
| Oncology | XGB-ALL | ECI | 0.0004 |
| Oncology | XGB-LIM | ECI | 0.0005 |
| Other | CS-LIM | ECI | 0.0168 |
| Other | RF-ALL | ECI | 0.0123 |
| Other | RF-LIM | ECI | 0.0140 |
| Other | SL | ECI | 0.0193 |
| Other | XGB-ALL | ECI | 0.0223 |
| Other | XGB-LIM | ECI | 0.0350 |
| Pediatrics | CS-LIM | ECI | 0.0054 |
| Pediatrics | RF-ALL | ECI | 0.0096 |
| Pediatrics | RF-LIM | ECI | 0.0203 |
| Pediatrics | SL | ECI | 0.0058 |
| Pediatrics | XGB-ALL | ECI | 0.0052 |
| Pediatrics | XGB-LIM | ECI | 0.0066 |
| Pneumology | CS-LIM | ECI | 0.0080 |
| Pneumology | RF-ALL | ECI | 0.0042 |
| Pneumology | RF-LIM | ECI | 0.0066 |
| Pneumology | SL | ECI | 0.0024 |
| Pneumology | XGB-ALL | ECI | 0.0035 |
| Pneumology | XGB-LIM | ECI | 0.0097 |
| Thoracic Surgery | CS-LIM | ECI | 0.0067 |
| Thoracic Surgery | RF-ALL | ECI | 0.0057 |
| Thoracic Surgery | RF-LIM | ECI | 0.0059 |
| Thoracic Surgery | SL | ECI | 0.0072 |
| Thoracic Surgery | XGB-ALL | ECI | 0.0062 |
| Thoracic Surgery | XGB-LIM | ECI | 0.0050 |
| Transplant | CS-LIM | ECI | 0.0116 |
| Transplant | RF-ALL | ECI | 0.0056 |
| Transplant | RF-LIM | ECI | 0.0096 |
| Transplant | SL | ECI | 0.0061 |
| Transplant | XGB-ALL | ECI | 0.0069 |
| Transplant | XGB-LIM | ECI | 0.0107 |
| Traumatology | CS-LIM | ECI | 0.0021 |
| Traumatology | RF-ALL | ECI | 0.0042 |
| Traumatology | RF-LIM | ECI | 0.0033 |
| Traumatology | SL | ECI | 0.0013 |
| Traumatology | XGB-ALL | ECI | 0.0006 |
| Traumatology | XGB-LIM | ECI | 0.0012 |
| UNKNOWN_VALUE | CS-LIM | ECI | 0.0113 |
| UNKNOWN_VALUE | RF-ALL | ECI | 0.0105 |
| UNKNOWN_VALUE | RF-LIM | ECI | 0.0175 |
| UNKNOWN_VALUE | SL | ECI | 0.0056 |
| UNKNOWN_VALUE | XGB-ALL | ECI | 0.0035 |
| UNKNOWN_VALUE | XGB-LIM | ECI | 0.0065 |
| Urology | CS-LIM | ECI | 0.0276 |
| Urology | RF-ALL | ECI | 0.0363 |
| Urology | RF-LIM | ECI | 0.0139 |
| Urology | SL | ECI | 0.0071 |
| Urology | XGB-ALL | ECI | 0.0043 |
| Urology | XGB-LIM | ECI | 0.0090 |
| NA | CS-LIM | ECI | NA |
| NA | RF-ALL | ECI | NA |
| NA | RF-LIM | ECI | NA |
| NA | SL | ECI | NA |
| NA | XGB-ALL | ECI | NA |
| NA | XGB-LIM | ECI | NA |

Table 9: Threshold independent metrics at different thresholds for each ward. AUPRC = Area Under the Precision Recall Curve; AUROC = Area Under the ROC curve; BSS = Brier Skill Score; E:O = Expected:Observed; ECI = Estimated Calibration Index. UNKNOWN_VALUE = The paediatric ward has changed name over time and the mapping done in training data did not cover the values in the test data.

#### Threshold dependent metric by ward

The RF-ALL model is the only model with utility in all wards at threshold 0.5%. At threshold 1%, the superlearner model has negative net benefit only in the paediatrics ward. To be noted though that paediatrics has changed name over time and the mapping done in training data did not cover the values in the test data.

| Model | n 0.005 | n 0.01 | n 0.02 | n 0.03 | n 0.04 | n 0.05 |
| --- | --- | --- | --- | --- | --- | --- |
| CS-LIM | 4 | 5 | 7 | 9 | 12 | 14 |
| RF-ALL | 0 | 4 | 6 | 8 | 10 | 11 |
| RF-LIM | 1 | 5 | 8 | 7 | 11 | 12 |
| SL | 2 | 1 | 3 | 4 | 7 | 12 |
| XGB-ALL | 3 | 2 | 4 | 5 | 8 | 12 |
| XGB-LIM | 2 | 2 | 5 | 6 | 8 | 12 |

Table 10: Number of wards per model on which Net Benefit is lower than the Net Benefit of treat all action or is negative.

| Model | Ward | Number of LMs | Number of CLABSI events in 7 days | 0.005 | 0.01 | 0.02 | 0.03 | 0.04 | 0.05 | 0.15 | 0.2 | 0.25 |
| --- | --- | --- | --- | --- | --- | --- | --- | --- | --- | --- | --- | --- |
| CS-LIM | ICU | 70273 | 1603 | 0.02 | 0.01* | 0.01* | 0* | 0* | -0 | -0 | -0* | -0 |
| CS-LIM | Abdomen | 59908 | 1171 | 0.01 | 0.01* | 0* | -0 | -0 | -0 | -0 | 0* | 0 |
| CS-LIM | Oncology | 32453 | 362 | 0.01* | 0* | 0* | 0* | 0* | 0* | -0 | 0* | 0 |
| CS-LIM | Hematology | 31359 | 1008 | 0.03 | 0.02* | 0.01* | 0* | 0* | -0 | -0 | -0* | -0 |
| CS-LIM | Pneumology | 29561 | 274 | 0* | -0 | -0 | -0 | -0 | -0 | 0 | 0* | 0 |
| CS-LIM | Traumatology | 22602 | 131 | 0* | 0* | 0* | 0* | 0* | 0* | 0 | 0* | 0 |
| CS-LIM | Cardiac | 21721 | 113 | 0* | 0* | 0* | 0* | -0 | -0 | 0 | 0* | 0 |
| CS-LIM | UNKNOWN_VALUE | 19808 | 195 | 0* | -0 | -0 | -0 | -0 | -0 | -0 | 0* | 0 |
| CS-LIM | Gynecology | 16706 | 113 | 0* | -0 | 0* | 0* | 0* | 0* | 0 | 0* | 0 |
| CS-LIM | Neonatology | 12672 | 274 | 0.02* | 0.01* | 0* | -0 | -0 | -0 | -0 | -0* | -0 |
| CS-LIM | Thoracic Surgery | 10985 | 134 | 0.01* | 0.01* | 0* | 0* | -0 | -0 | 0 | 0* | 0 |
| CS-LIM | Geriatrics | 9906 | 192 | 0.01 | 0.01* | 0.01* | 0.01* | 0* | 0* | -0 | 0* | 0 |
| CS-LIM | Transplant | 9277 | 70 | 0* | -0 | -0 | 0* | -0 | 0* | 0 | 0* | 0 |
| CS-LIM | Neuro | 8227 | 131 | 0.01* | 0.01* | 0* | 0* | 0* | 0* | 0 | 0* | 0 |
| CS-LIM | Internal Medicine | 6794 | 111 | 0.01* | 0.01* | 0* | 0* | 0* | 0* | -0 | 0* | 0 |
| CS-LIM | Pediatrics | 5530 | 31 | 0* | 0* | -0 | -0 | -0 | -0 | 0 | 0* | 0 |
| CS-LIM | Nephrology | 5035 | 56 | 0.01* | 0* | -0 | -0 | -0 | -0 | 0 | 0* | 0 |
| CS-LIM | Urology | 4854 | 86 | 0.01* | 0.01* | 0.01* | 0.01* | 0* | -0 | 0 | 0* | 0 |
| CS-LIM | Endocrinology | 4015 | 67 | 0.01* | 0* | 0* | -0 | -0 | -0 | 0 | 0* | 0 |
| CS-LIM | Emergency | 3730 | 39 | 0.01* | 0* | -0 | -0 | -0 | -0 | 0 | 0* | 0 |
| CS-LIM | ORL | 3569 | 48 | 0.01* | 0* | 0* | 0* | 0* | 0* | 0 | 0* | 0 |
| CS-LIM | Other | 1451 | 8 | 0* | -0 | -0 | -0 | -0 | -0 | 0 | 0* | 0 |
| RF-ALL | ICU | 70273 | 1603 | 0.02* | 0.01* | 0* | -0 | -0 | -0 | 0 | 0* | 0 |
| RF-ALL | Abdomen | 59908 | 1171 | 0.01* | 0.01* | 0* | -0 | -0 | 0* | 0 | 0* | 0 |
| RF-ALL | Oncology | 32453 | 362 | 0.01* | 0* | 0* | 0* | 0* | 0* | 0 | 0* | 0 |
| RF-ALL | Hematology | 31359 | 1008 | 0.03* | 0.02* | 0.01* | 0.01* | 0* | 0* | 0 | 0* | 0 |
| RF-ALL | Pneumology | 29561 | 274 | 0* | 0* | -0 | -0 | 0* | 0* | 0 | 0* | 0 |
| RF-ALL | Traumatology | 22602 | 131 | 0* | -0 | 0* | 0* | 0* | 0* | 0 | 0* | 0 |
| RF-ALL | Cardiac | 21721 | 113 | 0* | 0* | 0* | 0* | -0 | -0 | 0 | 0* | 0 |
| RF-ALL | UNKNOWN_VALUE | 19808 | 195 | 0* | -0 | -0 | 0* | -0 | -0 | 0 | 0* | 0 |
| RF-ALL | Gynecology | 16706 | 113 | 0* | 0* | 0* | 0* | 0* | -0 | 0 | 0* | 0 |
| RF-ALL | Neonatology | 12672 | 274 | 0.02* | 0.01* | 0.01* | 0* | 0* | 0* | 0 | 0* | 0 |
| RF-ALL | Thoracic Surgery | 10985 | 134 | 0.01* | 0* | 0* | 0* | 0* | 0* | 0 | 0* | 0 |
| RF-ALL | Geriatrics | 9906 | 192 | 0.02* | 0.01* | 0.01* | 0.01* | 0* | 0* | 0 | 0* | 0 |
| RF-ALL | Transplant | 9277 | 70 | 0* | 0* | 0* | 0* | -0 | -0 | 0 | 0* | 0 |
| RF-ALL | Neuro | 8227 | 131 | 0.01* | 0.01* | 0* | 0* | 0* | 0* | 0 | 0* | 0 |
| RF-ALL | Internal Medicine | 6794 | 111 | 0.01* | 0.01* | 0* | 0* | 0* | 0* | 0 | 0* | 0 |
| RF-ALL | Pediatrics | 5530 | 31 | 0* | -0 | -0 | -0 | -0 | 0 | 0 | 0* | 0 |
| RF-ALL | Nephrology | 5035 | 56 | 0.01* | 0* | -0 | -0 | -0 | -0 | 0 | 0* | 0 |
| RF-ALL | Urology | 4854 | 86 | 0.01* | 0.01* | 0.01* | 0* | 0* | 0* | 0 | 0* | 0 |
| RF-ALL | Endocrinology | 4015 | 67 | 0.01* | 0* | 0* | -0 | -0 | -0 | 0 | 0* | 0 |
| RF-ALL | Emergency | 3730 | 39 | 0.01* | 0* | -0 | -0 | -0 | -0 | 0 | 0* | 0 |
| RF-ALL | ORL | 3569 | 48 | 0.01* | 0* | 0* | 0* | 0* | -0 | 0 | 0* | 0 |
| RF-ALL | Other | 1451 | 8 | 0* | -0 | -0 | -0 | -0 | 0 | 0 | 0* | 0 |
| RF-LIM | ICU | 70273 | 1603 | 0.02* | 0.01* | 0* | 0* | -0 | -0 | 0 | 0* | 0 |
| RF-LIM | Abdomen | 59908 | 1171 | 0.01* | 0.01* | 0* | -0 | -0 | -0 | 0 | 0* | 0 |
| RF-LIM | Oncology | 32453 | 362 | 0.01* | 0* | 0* | 0* | 0* | 0* | 0 | 0* | 0 |
| RF-LIM | Hematology | 31359 | 1008 | 0.03* | 0.02* | 0.01* | 0.01* | 0* | 0* | -0 | 0* | 0 |
| RF-LIM | Pneumology | 29561 | 274 | 0* | 0* | -0 | -0 | -0 | -0 | 0 | 0* | 0 |
| RF-LIM | Traumatology | 22602 | 131 | 0* | -0 | 0* | 0* | 0* | 0* | 0 | 0* | 0 |
| RF-LIM | Cardiac | 21721 | 113 | 0* | -0 | 0* | 0* | -0 | -0 | 0 | 0* | 0 |
| RF-LIM | UNKNOWN_VALUE | 19808 | 195 | 0* | -0 | -0 | -0 | -0 | 0* | 0 | 0* | 0 |
| RF-LIM | Gynecology | 16706 | 113 | 0* | 0* | -0 | 0* | 0* | -0 | 0 | 0* | 0 |
| RF-LIM | Neonatology | 12672 | 274 | 0.02* | 0.01* | 0* | 0* | 0* | 0* | 0 | 0* | 0 |
| RF-LIM | Thoracic Surgery | 10985 | 134 | 0.01* | 0* | 0* | 0* | 0* | 0* | 0 | 0* | 0 |
| RF-LIM | Geriatrics | 9906 | 192 | 0.01 | 0.01* | 0.01* | 0.01* | 0* | 0* | 0 | 0* | 0 |
| RF-LIM | Transplant | 9277 | 70 | 0* | 0* | -0 | 0* | -0 | -0 | 0 | 0* | 0 |
| RF-LIM | Neuro | 8227 | 131 | 0.01* | 0.01* | 0* | 0* | 0* | 0* | -0 | 0* | 0 |
| RF-LIM | Internal Medicine | 6794 | 111 | 0.01* | 0.01* | 0* | 0* | 0* | -0 | 0 | 0* | 0 |
| RF-LIM | Pediatrics | 5530 | 31 | 0* | -0 | -0 | -0 | -0 | 0* | 0 | 0* | 0 |
| RF-LIM | Nephrology | 5035 | 56 | 0.01* | 0* | -0 | -0 | -0 | -0 | 0 | 0* | 0 |
| RF-LIM | Urology | 4854 | 86 | 0.01* | 0.01* | 0.01* | 0.01* | 0* | 0* | 0 | 0* | 0 |
| RF-LIM | Endocrinology | 4015 | 67 | 0.01* | 0.01* | 0* | 0* | 0* | -0 | 0 | 0* | 0 |
| RF-LIM | Emergency | 3730 | 39 | 0.01* | 0* | -0 | -0 | -0 | -0 | 0 | 0* | 0 |
| RF-LIM | ORL | 3569 | 48 | 0.01* | 0* | 0* | 0* | -0 | -0 | 0 | 0* | 0 |
| RF-LIM | Other | 1451 | 8 | 0* | -0 | -0 | -0 | -0 | 0 | 0 | 0* | 0 |
| SL | ICU | 70273 | 1603 | 0.02* | 0.01* | 0.01* | 0* | 0* | -0 | -0 | -0* | -0 |
| SL | Abdomen | 59908 | 1171 | 0.01* | 0.01* | 0* | 0* | -0 | -0 | -0 | -0* | 0 |
| SL | Oncology | 32453 | 362 | 0.01* | 0* | 0* | 0* | 0* | -0 | 0 | 0* | 0 |
| SL | Hematology | 31359 | 1008 | 0.03* | 0.02* | 0.01* | 0.01* | 0.01* | 0* | 0* | -0* | 0 |
| SL | Pneumology | 29561 | 274 | 0* | 0* | 0* | 0* | -0 | -0 | 0 | 0* | 0 |
| SL | Traumatology | 22602 | 131 | 0* | 0* | 0* | 0* | 0* | 0* | 0 | 0* | 0 |
| SL | Cardiac | 21721 | 113 | 0* | 0* | 0* | 0* | -0 | -0 | 0 | 0* | 0 |
| SL | UNKNOWN_VALUE | 19808 | 195 | 0* | 0* | 0* | 0* | 0* | 0* | 0 | 0* | 0 |
| SL | Gynecology | 16706 | 113 | 0* | 0* | 0* | 0* | 0* | 0* | 0* | 0* | 0 |
| SL | Neonatology | 12672 | 274 | 0.02* | 0.01* | 0.01* | 0.01* | 0* | 0* | -0 | 0* | 0 |
| SL | Thoracic Surgery | 10985 | 134 | 0.01* | 0.01* | 0* | 0* | 0* | 0* | 0 | 0* | 0 |
| SL | Geriatrics | 9906 | 192 | 0.01 | 0.01* | 0.01* | 0.01* | 0* | 0* | -0 | 0* | 0 |
| SL | Transplant | 9277 | 70 | 0* | 0* | 0* | 0* | 0* | -0 | -0 | 0* | 0 |
| SL | Neuro | 8227 | 131 | 0.01* | 0.01* | 0* | 0* | 0* | 0* | 0 | 0* | 0 |
| SL | Internal Medicine | 6794 | 111 | 0.01* | 0.01* | 0* | 0* | 0* | 0* | -0 | 0* | 0 |
| SL | Pediatrics | 5530 | 31 | 0* | -0 | 0* | -0 | -0 | -0 | 0 | 0* | 0 |
| SL | Nephrology | 5035 | 56 | 0.01* | 0* | -0 | -0 | -0 | -0 | -0 | 0* | 0 |
| SL | Urology | 4854 | 86 | 0.01* | 0.01* | 0.01* | 0.01* | 0* | 0* | 0 | 0* | 0 |
| SL | Endocrinology | 4015 | 67 | 0.01* | 0.01* | 0* | 0* | -0 | -0 | 0 | 0* | 0 |
| SL | Emergency | 3730 | 39 | 0.01 | 0* | -0 | -0 | 0* | -0 | 0 | 0* | 0 |
| SL | ORL | 3569 | 48 | 0.01* | 0.01* | 0* | 0* | 0* | -0 | 0 | 0* | 0 |
| SL | Other | 1451 | 8 | 0* | 0* | -0 | -0 | -0 | -0 | 0 | 0* | 0 |
| XGB-ALL | ICU | 70273 | 1603 | 0.02* | 0.01* | 0.01* | 0* | 0* | -0 | -0 | -0* | -0 |
| XGB-ALL | Abdomen | 59908 | 1171 | 0.01* | 0.01* | 0* | 0* | -0 | -0 | -0 | -0* | -0 |
| XGB-ALL | Oncology | 32453 | 362 | 0.01* | 0* | 0* | 0* | -0 | -0 | 0* | 0* | 0 |
| XGB-ALL | Hematology | 31359 | 1008 | 0.03 | 0.02* | 0.01* | 0.01* | 0* | 0* | 0* | 0* | -0 |
| XGB-ALL | Pneumology | 29561 | 274 | 0* | 0* | 0* | -0 | -0 | -0 | -0 | 0* | 0 |
| XGB-ALL | Traumatology | 22602 | 131 | 0* | 0* | 0* | 0* | 0* | 0* | 0* | 0* | 0 |
| XGB-ALL | Cardiac | 21721 | 113 | 0* | 0* | 0* | 0* | -0 | -0 | -0 | 0* | 0 |
| XGB-ALL | UNKNOWN_VALUE | 19808 | 195 | 0* | 0* | 0* | 0* | 0* | 0* | 0* | 0* | 0 |
| XGB-ALL | Gynecology | 16706 | 113 | 0* | 0* | 0* | 0* | 0* | 0* | 0* | 0* | 0 |
| XGB-ALL | Neonatology | 12672 | 274 | 0.02* | 0.01* | 0.01* | 0.01* | 0* | 0* | 0* | -0* | 0 |
| XGB-ALL | Thoracic Surgery | 10985 | 134 | 0.01* | 0.01* | 0* | 0* | 0* | 0* | -0 | 0* | 0 |
| XGB-ALL | Geriatrics | 9906 | 192 | 0.02* | 0.01* | 0.01* | 0.01* | 0* | 0* | -0 | -0* | 0 |
| XGB-ALL | Transplant | 9277 | 70 | 0* | 0* | 0* | 0* | 0* | -0 | -0 | -0* | 0 |
| XGB-ALL | Neuro | 8227 | 131 | 0.01* | 0.01* | 0* | 0* | 0* | 0* | 0 | 0* | 0 |
| XGB-ALL | Internal Medicine | 6794 | 111 | 0.01* | 0.01 | 0* | 0* | 0* | 0* | -0 | -0* | 0 |
| XGB-ALL | Pediatrics | 5530 | 31 | 0* | 0* | -0 | -0 | -0 | -0 | -0 | 0* | 0 |
| XGB-ALL | Nephrology | 5035 | 56 | 0.01 | 0* | -0 | -0 | -0 | -0 | -0 | -0* | 0 |
| XGB-ALL | Urology | 4854 | 86 | 0.01* | 0.01* | 0.01* | 0* | 0* | 0* | -0 | 0* | 0 |
| XGB-ALL | Endocrinology | 4015 | 67 | 0.01* | 0.01* | 0* | 0* | -0 | -0 | -0 | 0* | 0 |
| XGB-ALL | Emergency | 3730 | 39 | 0.01 | 0* | -0 | -0 | 0* | -0 | -0 | 0* | 0 |
| XGB-ALL | ORL | 3569 | 48 | 0.01* | 0.01* | 0* | 0* | 0* | -0 | 0 | 0* | 0 |
| XGB-ALL | Other | 1451 | 8 | 0* | -0 | -0 | -0 | -0 | -0 | 0 | 0* | 0 |
| XGB-LIM | ICU | 70273 | 1603 | 0.02* | 0.01* | 0.01* | 0* | -0 | -0 | -0 | -0* | -0 |
| XGB-LIM | Abdomen | 59908 | 1171 | 0.01* | 0.01* | 0* | 0* | -0 | -0 | -0 | -0* | 0 |
| XGB-LIM | Oncology | 32453 | 362 | 0.01* | 0* | 0* | 0* | 0* | 0* | -0 | -0* | 0 |
| XGB-LIM | Hematology | 31359 | 1008 | 0.03 | 0.02 | 0.01* | 0.01* | 0* | 0* | -0 | -0* | -0 |
| XGB-LIM | Pneumology | 29561 | 274 | 0* | 0* | -0 | -0 | -0 | -0 | -0 | 0* | 0 |
| XGB-LIM | Traumatology | 22602 | 131 | 0* | 0* | 0* | 0* | 0* | -0 | 0 | 0* | 0 |
| XGB-LIM | Cardiac | 21721 | 113 | 0* | 0* | 0* | 0* | -0 | -0 | 0 | 0* | 0 |
| XGB-LIM | UNKNOWN_VALUE | 19808 | 195 | 0* | 0* | 0* | -0 | 0* | -0 | 0* | 0* | 0 |
| XGB-LIM | Gynecology | 16706 | 113 | 0* | 0* | 0* | 0* | 0* | 0* | 0 | 0* | 0 |
| XGB-LIM | Neonatology | 12672 | 274 | 0.02* | 0.01* | 0.01* | 0* | 0* | 0* | 0* | -0* | 0 |
| XGB-LIM | Thoracic Surgery | 10985 | 134 | 0.01* | 0.01* | 0* | 0* | 0* | 0* | -0 | 0* | 0 |
| XGB-LIM | Geriatrics | 9906 | 192 | 0.02* | 0.01* | 0.01* | 0.01* | 0.01* | 0* | -0 | -0* | 0 |
| XGB-LIM | Transplant | 9277 | 70 | 0* | 0* | 0* | 0* | 0* | -0 | 0 | 0* | 0 |
| XGB-LIM | Neuro | 8227 | 131 | 0.01* | 0.01* | 0* | 0* | 0* | 0* | 0 | 0* | 0 |
| XGB-LIM | Internal Medicine | 6794 | 111 | 0.01* | 0.01 | 0* | 0* | 0* | 0* | 0 | 0* | 0 |
| XGB-LIM | Pediatrics | 5530 | 31 | 0* | 0* | -0 | -0 | -0 | -0 | -0 | 0* | 0 |
| XGB-LIM | Nephrology | 5035 | 56 | 0.01* | 0* | -0 | -0 | -0 | -0 | 0 | 0* | 0 |
| XGB-LIM | Urology | 4854 | 86 | 0.01* | 0.01* | 0.01* | 0.01* | 0* | 0* | 0 | 0* | 0 |
| XGB-LIM | Endocrinology | 4015 | 67 | 0.01* | 0.01* | 0* | 0* | 0* | -0 | 0 | 0* | 0 |
| XGB-LIM | Emergency | 3730 | 39 | 0.01 | 0* | -0 | -0 | -0 | -0 | 0 | 0* | 0 |
| XGB-LIM | ORL | 3569 | 48 | 0.01* | 0.01* | 0* | 0* | 0* | 0* | 0 | 0* | 0 |
| XGB-LIM | Other | 1451 | 8 | 0* | 0* | -0 | -0 | -0 | -0 | 0 | 0* | 0 |

Table 11: Net benefit at different thresholds for each ward. UNKNOWN_VALUE = The paediatric ward has changed name over time and the mapping done in training data did not cover the values in the test data.

| Model | Ward | Number of LMs | Number of CLABSI events in 7 days | metric | 0.005 | 0.01 | 0.02 | 0.03 | 0.04 | 0.05 | 0.15 | 0.2 | 0.25 |
| --- | --- | --- | --- | --- | --- | --- | --- | --- | --- | --- | --- | --- | --- |
| CS-LIM | ICU | 70273 | 1603 | Sensitivity | 1 | 0.97 | 0.77 | 0.61 | 0.48 | 0.38 | 0.03 | 0.01 | +0 |
| CS-LIM | ICU | 70273 | 1603 | Specificity | 0.01 | 0.07 | 0.39 | 0.61 | 0.74 | 0.82 | 0.99 | 1 | 1 |
| CS-LIM | ICU | 70273 | 1603 | Alert rate | 0.99 | 0.93 | 0.62 | 0.4 | 0.26 | 0.18 | 0.01 | +0 | +0 |
| CS-LIM | ICU | 70273 | 1603 | PPV | 0.02 | 0.02 | 0.03 | 0.03 | 0.04 | 0.05 | 0.06 | 0.06 | 0.04 |
| CS-LIM | ICU | 70273 | 1603 | NPV | 0.99 | 0.99 | 0.99 | 0.99 | 0.98 | 0.98 | 0.98 | 0.98 | 0.98 |
| RF-ALL | ICU | 70273 | 1603 | Sensitivity | 1 | 1 | 0.95 | 0.8 | 0.63 | 0.45 | +0 | +0 | +0 |
| RF-ALL | ICU | 70273 | 1603 | Specificity | 0.01 | 0.03 | 0.14 | 0.39 | 0.61 | 0.77 | 1 | 1 | 1 |
| RF-ALL | ICU | 70273 | 1603 | Alert rate | 0.99 | 0.97 | 0.86 | 0.61 | 0.39 | 0.23 | +0 | +0 | +0 |
| RF-ALL | ICU | 70273 | 1603 | PPV | 0.02 | 0.02 | 0.02 | 0.03 | 0.04 | 0.04 | NaN | NaN | NaN |
| RF-ALL | ICU | 70273 | 1603 | NPV | 1 | 1 | 0.99 | 0.99 | 0.99 | 0.98 | 0.98 | 0.98 | 0.98 |
| RF-LIM | ICU | 70273 | 1603 | Sensitivity | 1 | 1 | 0.93 | 0.76 | 0.56 | 0.37 | +0 | +0 | +0 |
| RF-LIM | ICU | 70273 | 1603 | Specificity | 0.01 | 0.03 | 0.18 | 0.46 | 0.67 | 0.82 | 1 | 1 | 1 |
| RF-LIM | ICU | 70273 | 1603 | Alert rate | 0.99 | 0.97 | 0.82 | 0.55 | 0.34 | 0.19 | +0 | +0 | +0 |
| RF-LIM | ICU | 70273 | 1603 | PPV | 0.02 | 0.02 | 0.03 | 0.03 | 0.04 | 0.04 | NaN | NaN | NaN |
| RF-LIM | ICU | 70273 | 1603 | NPV | 1 | 1 | 0.99 | 0.99 | 0.99 | 0.98 | 0.98 | 0.98 | 0.98 |
| SL | ICU | 70273 | 1603 | Sensitivity | 1 | 0.99 | 0.91 | 0.74 | 0.6 | 0.47 | 0.02 | 0.01 | +0 |
| SL | ICU | 70273 | 1603 | Specificity | 0.01 | 0.05 | 0.27 | 0.51 | 0.67 | 0.78 | 0.99 | 1 | 1 |
| SL | ICU | 70273 | 1603 | Alert rate | 0.99 | 0.95 | 0.74 | 0.5 | 0.33 | 0.23 | 0.01 | +0 | +0 |
| SL | ICU | 70273 | 1603 | PPV | 0.02 | 0.02 | 0.03 | 0.03 | 0.04 | 0.05 | 0.08 | 0.16 | 0.06 |
| SL | ICU | 70273 | 1603 | NPV | 1 | 1 | 0.99 | 0.99 | 0.99 | 0.98 | 0.98 | 0.98 | 0.98 |
| XGB-ALL | ICU | 70273 | 1603 | Sensitivity | 1 | 0.97 | 0.86 | 0.69 | 0.58 | 0.46 | 0.06 | 0.02 | 0.01 |
| XGB-ALL | ICU | 70273 | 1603 | Specificity | 0.02 | 0.11 | 0.36 | 0.56 | 0.69 | 0.77 | 0.99 | 1 | 1 |
| XGB-ALL | ICU | 70273 | 1603 | Alert rate | 0.98 | 0.89 | 0.64 | 0.45 | 0.32 | 0.23 | 0.02 | 0.01 | +0 |
| XGB-ALL | ICU | 70273 | 1603 | PPV | 0.02 | 0.02 | 0.03 | 0.03 | 0.04 | 0.04 | 0.09 | 0.07 | 0.14 |
| XGB-ALL | ICU | 70273 | 1603 | NPV | 1 | 0.99 | 0.99 | 0.99 | 0.99 | 0.98 | 0.98 | 0.98 | 0.98 |
| XGB-LIM | ICU | 70273 | 1603 | Sensitivity | 1 | 0.99 | 0.88 | 0.71 | 0.57 | 0.44 | 0.04 | 0.01 | +0 |
| XGB-LIM | ICU | 70273 | 1603 | Specificity | 0.02 | 0.07 | 0.3 | 0.52 | 0.67 | 0.77 | 0.99 | 1 | 1 |
| XGB-LIM | ICU | 70273 | 1603 | Alert rate | 0.98 | 0.93 | 0.7 | 0.49 | 0.34 | 0.24 | 0.01 | +0 | +0 |
| XGB-LIM | ICU | 70273 | 1603 | PPV | 0.02 | 0.02 | 0.03 | 0.03 | 0.04 | 0.04 | 0.08 | 0.07 | 0.07 |
| XGB-LIM | ICU | 70273 | 1603 | NPV | 1 | 1 | 0.99 | 0.99 | 0.99 | 0.98 | 0.98 | 0.98 | 0.98 |
| CS-LIM | Abdomen | 59908 | 1171 | Sensitivity | 0.98 | 0.94 | 0.64 | 0.46 | 0.29 | 0.15 | +0 | +0 | +0 |
| CS-LIM | Abdomen | 59908 | 1171 | Specificity | 0.02 | 0.15 | 0.53 | 0.69 | 0.83 | 0.92 | 1 | 1 | 1 |
| CS-LIM | Abdomen | 59908 | 1171 | Alert rate | 0.98 | 0.86 | 0.47 | 0.31 | 0.18 | 0.08 | +0 | +0 | +0 |
| CS-LIM | Abdomen | 59908 | 1171 | PPV | 0.02 | 0.02 | 0.03 | 0.03 | 0.03 | 0.03 | +0 | NaN | NaN |
| CS-LIM | Abdomen | 59908 | 1171 | NPV | 0.98 | 0.99 | 0.99 | 0.99 | 0.98 | 0.98 | 0.98 | 0.98 | 0.98 |
| RF-ALL | Abdomen | 59908 | 1171 | Sensitivity | 1 | 0.96 | 0.67 | 0.46 | 0.27 | 0.07 | +0 | +0 | +0 |
| RF-ALL | Abdomen | 59908 | 1171 | Specificity | 0.04 | 0.11 | 0.52 | 0.71 | 0.87 | 0.97 | 1 | 1 | 1 |
| RF-ALL | Abdomen | 59908 | 1171 | Alert rate | 0.96 | 0.89 | 0.48 | 0.3 | 0.14 | 0.03 | +0 | +0 | +0 |
| RF-ALL | Abdomen | 59908 | 1171 | PPV | 0.02 | 0.02 | 0.03 | 0.03 | 0.04 | 0.05 | NaN | NaN | NaN |
| RF-ALL | Abdomen | 59908 | 1171 | NPV | 1 | 0.99 | 0.99 | 0.99 | 0.98 | 0.98 | 0.98 | 0.98 | 0.98 |
| RF-LIM | Abdomen | 59908 | 1171 | Sensitivity | 0.99 | 0.95 | 0.69 | 0.49 | 0.32 | 0.09 | +0 | +0 | +0 |
| RF-LIM | Abdomen | 59908 | 1171 | Specificity | 0.05 | 0.15 | 0.51 | 0.68 | 0.84 | 0.97 | 1 | 1 | 1 |
| RF-LIM | Abdomen | 59908 | 1171 | Alert rate | 0.95 | 0.86 | 0.49 | 0.33 | 0.16 | 0.03 | +0 | +0 | +0 |
| RF-LIM | Abdomen | 59908 | 1171 | PPV | 0.02 | 0.02 | 0.03 | 0.03 | 0.04 | 0.05 | NaN | NaN | NaN |
| RF-LIM | Abdomen | 59908 | 1171 | NPV | 1 | 0.99 | 0.99 | 0.99 | 0.98 | 0.98 | 0.98 | 0.98 | 0.98 |
| SL | Abdomen | 59908 | 1171 | Sensitivity | 1 | 0.96 | 0.74 | 0.51 | 0.31 | 0.2 | +0 | +0 | +0 |
| SL | Abdomen | 59908 | 1171 | Specificity | 0.05 | 0.17 | 0.51 | 0.7 | 0.84 | 0.91 | 1 | 1 | 1 |
| SL | Abdomen | 59908 | 1171 | Alert rate | 0.96 | 0.83 | 0.5 | 0.3 | 0.17 | 0.09 | +0 | +0 | +0 |
| SL | Abdomen | 59908 | 1171 | PPV | 0.02 | 0.02 | 0.03 | 0.03 | 0.04 | 0.04 | +0 | +0 | NaN |
| SL | Abdomen | 59908 | 1171 | NPV | 1 | 1 | 0.99 | 0.99 | 0.98 | 0.98 | 0.98 | 0.98 | 0.98 |
| XGB-ALL | Abdomen | 59908 | 1171 | Sensitivity | 0.99 | 0.94 | 0.73 | 0.51 | 0.34 | 0.23 | 0.01 | +0 | +0 |
| XGB-ALL | Abdomen | 59908 | 1171 | Specificity | 0.06 | 0.23 | 0.52 | 0.7 | 0.82 | 0.89 | 1 | 1 | 1 |
| XGB-ALL | Abdomen | 59908 | 1171 | Alert rate | 0.94 | 0.78 | 0.48 | 0.3 | 0.18 | 0.11 | +0 | +0 | +0 |
| XGB-ALL | Abdomen | 59908 | 1171 | PPV | 0.02 | 0.02 | 0.03 | 0.03 | 0.04 | 0.04 | 0.06 | +0 | +0 |
| XGB-ALL | Abdomen | 59908 | 1171 | NPV | 1 | 1 | 0.99 | 0.99 | 0.98 | 0.98 | 0.98 | 0.98 | 0.98 |
| XGB-LIM | Abdomen | 59908 | 1171 | Sensitivity | 0.99 | 0.93 | 0.72 | 0.51 | 0.35 | 0.23 | +0 | +0 | +0 |
| XGB-LIM | Abdomen | 59908 | 1171 | Specificity | 0.05 | 0.2 | 0.52 | 0.7 | 0.82 | 0.89 | 1 | 1 | 1 |
| XGB-LIM | Abdomen | 59908 | 1171 | Alert rate | 0.95 | 0.8 | 0.49 | 0.3 | 0.19 | 0.11 | +0 | +0 | +0 |
| XGB-LIM | Abdomen | 59908 | 1171 | PPV | 0.02 | 0.02 | 0.03 | 0.03 | 0.04 | 0.04 | 0.06 | +0 | NaN |
| XGB-LIM | Abdomen | 59908 | 1171 | NPV | 1 | 0.99 | 0.99 | 0.99 | 0.98 | 0.98 | 0.98 | 0.98 | 0.98 |
| CS-LIM | Oncology | 32453 | 362 | Sensitivity | 0.99 | 0.89 | 0.5 | 0.29 | 0.18 | 0.09 | +0 | +0 | +0 |
| CS-LIM | Oncology | 32453 | 362 | Specificity | 0.05 | 0.31 | 0.79 | 0.93 | 0.97 | 0.99 | 1 | 1 | 1 |
| CS-LIM | Oncology | 32453 | 362 | Alert rate | 0.95 | 0.69 | 0.21 | 0.07 | 0.03 | 0.02 | +0 | +0 | +0 |
| CS-LIM | Oncology | 32453 | 362 | PPV | 0.01 | 0.01 | 0.03 | 0.05 | 0.06 | 0.06 | +0 | NaN | NaN |
| CS-LIM | Oncology | 32453 | 362 | NPV | 1 | 1 | 0.99 | 0.99 | 0.99 | 0.99 | 0.99 | 0.99 | 0.99 |
| RF-ALL | Oncology | 32453 | 362 | Sensitivity | 0.97 | 0.85 | 0.36 | 0.18 | 0.08 | 0.01 | +0 | +0 | +0 |
| RF-ALL | Oncology | 32453 | 362 | Specificity | 0.18 | 0.41 | 0.88 | 0.97 | 0.99 | 1 | 1 | 1 | 1 |
| RF-ALL | Oncology | 32453 | 362 | Alert rate | 0.83 | 0.59 | 0.12 | 0.04 | 0.01 | +0 | +0 | +0 | +0 |
| RF-ALL | Oncology | 32453 | 362 | PPV | 0.01 | 0.02 | 0.03 | 0.06 | 0.08 | 0.11 | NaN | NaN | NaN |
| RF-ALL | Oncology | 32453 | 362 | NPV | 1 | 1 | 0.99 | 0.99 | 0.99 | 0.99 | 0.99 | 0.99 | 0.99 |
| RF-LIM | Oncology | 32453 | 362 | Sensitivity | 0.97 | 0.88 | 0.47 | 0.27 | 0.13 | 0.02 | +0 | +0 | +0 |
| RF-LIM | Oncology | 32453 | 362 | Specificity | 0.17 | 0.38 | 0.81 | 0.94 | 0.98 | 1 | 1 | 1 | 1 |
| RF-LIM | Oncology | 32453 | 362 | Alert rate | 0.83 | 0.63 | 0.19 | 0.06 | 0.02 | +0 | +0 | +0 | +0 |
| RF-LIM | Oncology | 32453 | 362 | PPV | 0.01 | 0.02 | 0.03 | 0.05 | 0.06 | 0.06 | NaN | NaN | NaN |
| RF-LIM | Oncology | 32453 | 362 | NPV | 1 | 1 | 0.99 | 0.99 | 0.99 | 0.99 | 0.99 | 0.99 | 0.99 |
| SL | Oncology | 32453 | 362 | Sensitivity | 0.96 | 0.86 | 0.44 | 0.24 | 0.1 | 0.05 | +0 | +0 | +0 |
| SL | Oncology | 32453 | 362 | Specificity | 0.22 | 0.48 | 0.86 | 0.95 | 0.98 | 0.99 | 1 | 1 | 1 |
| SL | Oncology | 32453 | 362 | Alert rate | 0.78 | 0.52 | 0.15 | 0.06 | 0.03 | 0.01 | +0 | +0 | +0 |
| SL | Oncology | 32453 | 362 | PPV | 0.01 | 0.02 | 0.03 | 0.05 | 0.04 | 0.04 | NaN | NaN | NaN |
| SL | Oncology | 32453 | 362 | NPV | 1 | 1 | 0.99 | 0.99 | 0.99 | 0.99 | 0.99 | 0.99 | 0.99 |
| XGB-ALL | Oncology | 32453 | 362 | Sensitivity | 0.96 | 0.8 | 0.43 | 0.27 | 0.12 | 0.06 | 0.01 | +0 | +0 |
| XGB-ALL | Oncology | 32453 | 362 | Specificity | 0.28 | 0.54 | 0.84 | 0.93 | 0.97 | 0.98 | 1 | 1 | 1 |
| XGB-ALL | Oncology | 32453 | 362 | Alert rate | 0.72 | 0.46 | 0.16 | 0.07 | 0.03 | 0.02 | +0 | +0 | +0 |
| XGB-ALL | Oncology | 32453 | 362 | PPV | 0.01 | 0.02 | 0.03 | 0.04 | 0.04 | 0.03 | 0.15 | NaN | NaN |
| XGB-ALL | Oncology | 32453 | 362 | NPV | 1 | 1 | 0.99 | 0.99 | 0.99 | 0.99 | 0.99 | 0.99 | 0.99 |
| XGB-LIM | Oncology | 32453 | 362 | Sensitivity | 0.96 | 0.84 | 0.48 | 0.25 | 0.16 | 0.09 | +0 | +0 | +0 |
| XGB-LIM | Oncology | 32453 | 362 | Specificity | 0.23 | 0.49 | 0.83 | 0.94 | 0.97 | 0.98 | 1 | 1 | 1 |
| XGB-LIM | Oncology | 32453 | 362 | Alert rate | 0.77 | 0.51 | 0.18 | 0.07 | 0.03 | 0.02 | +0 | +0 | +0 |
| XGB-LIM | Oncology | 32453 | 362 | PPV | 0.01 | 0.02 | 0.03 | 0.04 | 0.05 | 0.05 | +0 | +0 | NaN |
| XGB-LIM | Oncology | 32453 | 362 | NPV | 1 | 1 | 0.99 | 0.99 | 0.99 | 0.99 | 0.99 | 0.99 | 0.99 |
| CS-LIM | Hematology | 31359 | 1008 | Sensitivity | 1 | 0.98 | 0.81 | 0.56 | 0.41 | 0.33 | +0 | +0 | +0 |
| CS-LIM | Hematology | 31359 | 1008 | Specificity | 0.01 | 0.08 | 0.34 | 0.57 | 0.71 | 0.79 | 1 | 1 | 1 |
| CS-LIM | Hematology | 31359 | 1008 | Alert rate | 0.99 | 0.92 | 0.66 | 0.43 | 0.29 | 0.22 | +0 | +0 | +0 |
| CS-LIM | Hematology | 31359 | 1008 | PPV | 0.03 | 0.03 | 0.04 | 0.04 | 0.04 | 0.05 | 0.04 | 0.05 | +0 |
| CS-LIM | Hematology | 31359 | 1008 | NPV | 0.99 | 0.99 | 0.98 | 0.98 | 0.97 | 0.97 | 0.97 | 0.97 | 0.97 |
| RF-ALL | Hematology | 31359 | 1008 | Sensitivity | 1 | 1 | 0.93 | 0.77 | 0.56 | 0.21 | +0 | +0 | +0 |
| RF-ALL | Hematology | 31359 | 1008 | Specificity | 0.01 | 0.04 | 0.23 | 0.45 | 0.68 | 0.89 | 1 | 1 | 1 |
| RF-ALL | Hematology | 31359 | 1008 | Alert rate | 0.99 | 0.96 | 0.78 | 0.56 | 0.33 | 0.11 | +0 | +0 | +0 |
| RF-ALL | Hematology | 31359 | 1008 | PPV | 0.03 | 0.03 | 0.04 | 0.04 | 0.05 | 0.06 | NaN | NaN | NaN |
| RF-ALL | Hematology | 31359 | 1008 | NPV | 1 | 1 | 0.99 | 0.98 | 0.98 | 0.97 | 0.97 | 0.97 | 0.97 |
| RF-LIM | Hematology | 31359 | 1008 | Sensitivity | 1 | 1 | 0.9 | 0.75 | 0.52 | 0.29 | +0 | +0 | +0 |
| RF-LIM | Hematology | 31359 | 1008 | Specificity | 0.02 | 0.06 | 0.27 | 0.5 | 0.7 | 0.87 | 1 | 1 | 1 |
| RF-LIM | Hematology | 31359 | 1008 | Alert rate | 0.98 | 0.94 | 0.74 | 0.51 | 0.31 | 0.13 | +0 | +0 | +0 |
| RF-LIM | Hematology | 31359 | 1008 | PPV | 0.03 | 0.03 | 0.04 | 0.05 | 0.05 | 0.07 | +0 | NaN | NaN |
| RF-LIM | Hematology | 31359 | 1008 | NPV | 1 | 1 | 0.99 | 0.98 | 0.98 | 0.97 | 0.97 | 0.97 | 0.97 |
| SL | Hematology | 31359 | 1008 | Sensitivity | 1 | 0.98 | 0.86 | 0.7 | 0.55 | 0.38 | 0.01 | +0 | +0 |
| SL | Hematology | 31359 | 1008 | Specificity | 0.02 | 0.09 | 0.33 | 0.54 | 0.7 | 0.81 | 1 | 1 | 1 |
| SL | Hematology | 31359 | 1008 | Alert rate | 0.98 | 0.91 | 0.68 | 0.47 | 0.31 | 0.2 | +0 | +0 | +0 |
| SL | Hematology | 31359 | 1008 | PPV | 0.03 | 0.03 | 0.04 | 0.05 | 0.06 | 0.06 | 0.19 | 0.17 | NaN |
| SL | Hematology | 31359 | 1008 | NPV | 1 | 0.99 | 0.99 | 0.98 | 0.98 | 0.98 | 0.97 | 0.97 | 0.97 |
| XGB-ALL | Hematology | 31359 | 1008 | Sensitivity | 0.99 | 0.95 | 0.82 | 0.66 | 0.52 | 0.4 | 0.03 | 0.01 | +0 |
| XGB-ALL | Hematology | 31359 | 1008 | Specificity | 0.05 | 0.16 | 0.4 | 0.58 | 0.7 | 0.79 | 0.99 | 1 | 1 |
| XGB-ALL | Hematology | 31359 | 1008 | Alert rate | 0.95 | 0.84 | 0.61 | 0.43 | 0.31 | 0.22 | 0.01 | +0 | +0 |
| XGB-ALL | Hematology | 31359 | 1008 | PPV | 0.03 | 0.04 | 0.04 | 0.05 | 0.05 | 0.06 | 0.16 | 0.2 | 0.15 |
| XGB-ALL | Hematology | 31359 | 1008 | NPV | 0.99 | 0.99 | 0.99 | 0.98 | 0.98 | 0.98 | 0.97 | 0.97 | 0.97 |
| XGB-LIM | Hematology | 31359 | 1008 | Sensitivity | 0.98 | 0.95 | 0.82 | 0.65 | 0.5 | 0.39 | 0.02 | 0.01 | +0 |
| XGB-LIM | Hematology | 31359 | 1008 | Specificity | 0.03 | 0.14 | 0.38 | 0.56 | 0.69 | 0.79 | 0.99 | 1 | 1 |
| XGB-LIM | Hematology | 31359 | 1008 | Alert rate | 0.97 | 0.86 | 0.63 | 0.44 | 0.32 | 0.22 | 0.01 | +0 | +0 |
| XGB-LIM | Hematology | 31359 | 1008 | PPV | 0.03 | 0.03 | 0.04 | 0.05 | 0.05 | 0.06 | 0.12 | 0.15 | +0 |
| XGB-LIM | Hematology | 31359 | 1008 | NPV | 0.98 | 0.99 | 0.98 | 0.98 | 0.98 | 0.98 | 0.97 | 0.97 | 0.97 |
| CS-LIM | Pneumology | 29561 | 274 | Sensitivity | 0.99 | 0.84 | 0.32 | 0.19 | 0.12 | 0.08 | +0 | +0 | +0 |
| CS-LIM | Pneumology | 29561 | 274 | Specificity | 0.05 | 0.3 | 0.83 | 0.93 | 0.97 | 0.99 | 1 | 1 | 1 |
| CS-LIM | Pneumology | 29561 | 274 | Alert rate | 0.95 | 0.7 | 0.17 | 0.07 | 0.03 | 0.01 | +0 | +0 | +0 |
| CS-LIM | Pneumology | 29561 | 274 | PPV | 0.01 | 0.01 | 0.01 | 0.02 | 0.03 | 0.04 | NaN | NaN | NaN |
| CS-LIM | Pneumology | 29561 | 274 | NPV | 1 | 1 | 0.99 | 0.99 | 0.99 | 0.99 | 0.99 | 0.99 | 0.99 |
| RF-ALL | Pneumology | 29561 | 274 | Sensitivity | 1 | 0.9 | 0.38 | 0.16 | 0.07 | 0.01 | +0 | +0 | +0 |
| RF-ALL | Pneumology | 29561 | 274 | Specificity | 0.12 | 0.35 | 0.83 | 0.95 | 0.99 | 1 | 1 | 1 | 1 |
| RF-ALL | Pneumology | 29561 | 274 | Alert rate | 0.89 | 0.65 | 0.17 | 0.05 | 0.01 | +0 | +0 | +0 | +0 |
| RF-ALL | Pneumology | 29561 | 274 | PPV | 0.01 | 0.01 | 0.02 | 0.03 | 0.05 | 0.14 | NaN | NaN | NaN |
| RF-ALL | Pneumology | 29561 | 274 | NPV | 1 | 1 | 0.99 | 0.99 | 0.99 | 0.99 | 0.99 | 0.99 | 0.99 |
| RF-LIM | Pneumology | 29561 | 274 | Sensitivity | 1 | 0.89 | 0.41 | 0.21 | 0.1 | 0.01 | +0 | +0 | +0 |
| RF-LIM | Pneumology | 29561 | 274 | Specificity | 0.09 | 0.37 | 0.79 | 0.93 | 0.98 | 1 | 1 | 1 | 1 |
| RF-LIM | Pneumology | 29561 | 274 | Alert rate | 0.91 | 0.63 | 0.21 | 0.07 | 0.02 | +0 | +0 | +0 | +0 |
| RF-LIM | Pneumology | 29561 | 274 | PPV | 0.01 | 0.01 | 0.01 | 0.02 | 0.03 | 0.04 | NaN | NaN | NaN |
| RF-LIM | Pneumology | 29561 | 274 | NPV | 1 | 1 | 0.99 | 0.99 | 0.99 | 0.99 | 0.99 | 0.99 | 0.99 |
| SL | Pneumology | 29561 | 274 | Sensitivity | 0.96 | 0.68 | 0.32 | 0.19 | 0.09 | 0.05 | +0 | +0 | +0 |
| SL | Pneumology | 29561 | 274 | Specificity | 0.24 | 0.59 | 0.9 | 0.95 | 0.98 | 0.99 | 1 | 1 | 1 |
| SL | Pneumology | 29561 | 274 | Alert rate | 0.76 | 0.41 | 0.11 | 0.05 | 0.03 | 0.01 | +0 | +0 | +0 |
| SL | Pneumology | 29561 | 274 | PPV | 0.01 | 0.01 | 0.02 | 0.03 | 0.03 | 0.03 | NaN | NaN | NaN |
| SL | Pneumology | 29561 | 274 | NPV | 1 | 1 | 0.99 | 0.99 | 0.99 | 0.99 | 0.99 | 0.99 | 0.99 |
| XGB-ALL | Pneumology | 29561 | 274 | Sensitivity | 0.85 | 0.53 | 0.29 | 0.18 | 0.11 | 0.06 | +0 | +0 | +0 |
| XGB-ALL | Pneumology | 29561 | 274 | Specificity | 0.41 | 0.7 | 0.9 | 0.95 | 0.97 | 0.98 | 1 | 1 | 1 |
| XGB-ALL | Pneumology | 29561 | 274 | Alert rate | 0.59 | 0.3 | 0.1 | 0.05 | 0.03 | 0.02 | +0 | +0 | +0 |
| XGB-ALL | Pneumology | 29561 | 274 | PPV | 0.01 | 0.01 | 0.02 | 0.03 | 0.03 | 0.03 | +0 | NaN | NaN |
| XGB-ALL | Pneumology | 29561 | 274 | NPV | 1 | 0.99 | 0.99 | 0.99 | 0.99 | 0.99 | 0.99 | 0.99 | 0.99 |
| XGB-LIM | Pneumology | 29561 | 274 | Sensitivity | 0.97 | 0.72 | 0.32 | 0.2 | 0.14 | 0.09 | +0 | +0 | +0 |
| XGB-LIM | Pneumology | 29561 | 274 | Specificity | 0.27 | 0.57 | 0.86 | 0.93 | 0.96 | 0.97 | 1 | 1 | 1 |
| XGB-LIM | Pneumology | 29561 | 274 | Alert rate | 0.73 | 0.43 | 0.15 | 0.07 | 0.04 | 0.03 | +0 | +0 | +0 |
| XGB-LIM | Pneumology | 29561 | 274 | PPV | 0.01 | 0.01 | 0.02 | 0.02 | 0.02 | 0.02 | +0 | NaN | NaN |
| XGB-LIM | Pneumology | 29561 | 274 | NPV | 1 | 1 | 0.99 | 0.99 | 0.99 | 0.99 | 0.99 | 0.99 | 0.99 |
| CS-LIM | Traumatology | 22602 | 131 | Sensitivity | 0.95 | 0.38 | 0.18 | 0.09 | 0.06 | 0.06 | +0 | +0 | +0 |
| CS-LIM | Traumatology | 22602 | 131 | Specificity | 0.11 | 0.78 | 0.98 | 1 | 1 | 1 | 1 | 1 | 1 |
| CS-LIM | Traumatology | 22602 | 131 | Alert rate | 0.89 | 0.22 | 0.02 | 0.01 | +0 | +0 | +0 | +0 | +0 |
| CS-LIM | Traumatology | 22602 | 131 | PPV | 0.01 | 0.01 | 0.06 | 0.1 | 0.12 | 0.16 | NaN | NaN | NaN |
| CS-LIM | Traumatology | 22602 | 131 | NPV | 1 | 1 | 1 | 0.99 | 0.99 | 0.99 | 0.99 | 0.99 | 0.99 |
| RF-ALL | Traumatology | 22602 | 131 | Sensitivity | 0.98 | 0.76 | 0.32 | 0.14 | 0.03 | 0.02 | +0 | +0 | +0 |
| RF-ALL | Traumatology | 22602 | 131 | Specificity | 0.08 | 0.49 | 0.95 | 0.99 | 1 | 1 | 1 | 1 | 1 |
| RF-ALL | Traumatology | 22602 | 131 | Alert rate | 0.92 | 0.52 | 0.06 | 0.01 | +0 | +0 | +0 | +0 | +0 |
| RF-ALL | Traumatology | 22602 | 131 | PPV | 0.01 | 0.01 | 0.03 | 0.08 | 0.08 | 0.11 | NaN | NaN | NaN |
| RF-ALL | Traumatology | 22602 | 131 | NPV | 1 | 1 | 1 | 0.99 | 0.99 | 0.99 | 0.99 | 0.99 | 0.99 |
| RF-LIM | Traumatology | 22602 | 131 | Sensitivity | 0.99 | 0.73 | 0.25 | 0.12 | 0.08 | 0.02 | +0 | +0 | +0 |
| RF-LIM | Traumatology | 22602 | 131 | Specificity | 0.09 | 0.52 | 0.96 | 0.99 | 1 | 1 | 1 | 1 | 1 |
| RF-LIM | Traumatology | 22602 | 131 | Alert rate | 0.91 | 0.48 | 0.04 | 0.01 | +0 | +0 | +0 | +0 | +0 |
| RF-LIM | Traumatology | 22602 | 131 | PPV | 0.01 | 0.01 | 0.04 | 0.06 | 0.11 | 0.08 | NaN | NaN | NaN |
| RF-LIM | Traumatology | 22602 | 131 | NPV | 1 | 1 | 1 | 0.99 | 0.99 | 0.99 | 0.99 | 0.99 | 0.99 |
| SL | Traumatology | 22602 | 131 | Sensitivity | 0.94 | 0.6 | 0.18 | 0.11 | 0.06 | 0.03 | +0 | +0 | +0 |
| SL | Traumatology | 22602 | 131 | Specificity | 0.18 | 0.78 | 0.98 | 0.99 | 1 | 1 | 1 | 1 | 1 |
| SL | Traumatology | 22602 | 131 | Alert rate | 0.82 | 0.22 | 0.02 | 0.01 | +0 | +0 | +0 | +0 | +0 |
| SL | Traumatology | 22602 | 131 | PPV | 0.01 | 0.02 | 0.05 | 0.1 | 0.1 | 0.1 | NaN | NaN | NaN |
| SL | Traumatology | 22602 | 131 | NPV | 1 | 1 | 1 | 0.99 | 0.99 | 0.99 | 0.99 | 0.99 | 0.99 |
| XGB-ALL | Traumatology | 22602 | 131 | Sensitivity | 0.85 | 0.4 | 0.19 | 0.09 | 0.06 | 0.04 | 0.01 | +0 | +0 |
| XGB-ALL | Traumatology | 22602 | 131 | Specificity | 0.44 | 0.85 | 0.98 | 0.99 | 1 | 1 | 1 | 1 | 1 |
| XGB-ALL | Traumatology | 22602 | 131 | Alert rate | 0.56 | 0.15 | 0.02 | 0.01 | +0 | +0 | +0 | +0 | +0 |
| XGB-ALL | Traumatology | 22602 | 131 | PPV | 0.01 | 0.02 | 0.05 | 0.08 | 0.09 | 0.11 | 0.33 | NaN | NaN |
| XGB-ALL | Traumatology | 22602 | 131 | NPV | 1 | 1 | 1 | 0.99 | 0.99 | 0.99 | 0.99 | 0.99 | 0.99 |
| XGB-LIM | Traumatology | 22602 | 131 | Sensitivity | 0.92 | 0.5 | 0.17 | 0.12 | 0.04 | 0.02 | +0 | +0 | +0 |
| XGB-LIM | Traumatology | 22602 | 131 | Specificity | 0.23 | 0.76 | 0.97 | 0.99 | 1 | 1 | 1 | 1 | 1 |
| XGB-LIM | Traumatology | 22602 | 131 | Alert rate | 0.77 | 0.24 | 0.03 | 0.01 | +0 | +0 | +0 | +0 | +0 |
| XGB-LIM | Traumatology | 22602 | 131 | PPV | 0.01 | 0.01 | 0.04 | 0.08 | 0.06 | 0.05 | NaN | NaN | NaN |
| XGB-LIM | Traumatology | 22602 | 131 | NPV | 1 | 1 | 1 | 0.99 | 0.99 | 0.99 | 0.99 | 0.99 | 0.99 |
| CS-LIM | Cardiac | 21721 | 113 | Sensitivity | 0.89 | 0.65 | 0.32 | 0.16 | 0.02 | 0.01 | +0 | +0 | +0 |
| CS-LIM | Cardiac | 21721 | 113 | Specificity | 0.33 | 0.73 | 0.97 | 0.99 | 1 | 1 | 1 | 1 | 1 |
| CS-LIM | Cardiac | 21721 | 113 | Alert rate | 0.67 | 0.27 | 0.03 | 0.01 | +0 | +0 | +0 | +0 | +0 |
| CS-LIM | Cardiac | 21721 | 113 | PPV | 0.01 | 0.01 | 0.05 | 0.08 | 0.02 | 0.03 | NaN | NaN | NaN |
| CS-LIM | Cardiac | 21721 | 113 | NPV | 1 | 1 | 1 | 1 | 0.99 | 0.99 | 0.99 | 0.99 | 0.99 |
| RF-ALL | Cardiac | 21721 | 113 | Sensitivity | 0.92 | 0.8 | 0.39 | 0.13 | 0.03 | +0 | +0 | +0 | +0 |
| RF-ALL | Cardiac | 21721 | 113 | Specificity | 0.32 | 0.63 | 0.94 | 0.99 | 1 | 1 | 1 | 1 | 1 |
| RF-ALL | Cardiac | 21721 | 113 | Alert rate | 0.68 | 0.38 | 0.06 | 0.01 | +0 | +0 | +0 | +0 | +0 |
| RF-ALL | Cardiac | 21721 | 113 | PPV | 0.01 | 0.01 | 0.03 | 0.05 | 0.03 | +0 | NaN | NaN | NaN |
| RF-ALL | Cardiac | 21721 | 113 | NPV | 1 | 1 | 1 | 1 | 0.99 | 0.99 | 0.99 | 0.99 | 0.99 |
| RF-LIM | Cardiac | 21721 | 113 | Sensitivity | 0.93 | 0.79 | 0.41 | 0.16 | 0.06 | +0 | +0 | +0 | +0 |
| RF-LIM | Cardiac | 21721 | 113 | Specificity | 0.31 | 0.57 | 0.9 | 0.98 | 0.99 | 1 | 1 | 1 | 1 |
| RF-LIM | Cardiac | 21721 | 113 | Alert rate | 0.69 | 0.43 | 0.11 | 0.02 | 0.01 | +0 | +0 | +0 | +0 |
| RF-LIM | Cardiac | 21721 | 113 | PPV | 0.01 | 0.01 | 0.02 | 0.04 | 0.04 | +0 | NaN | NaN | NaN |
| RF-LIM | Cardiac | 21721 | 113 | NPV | 1 | 1 | 1 | 1 | 0.99 | 0.99 | 0.99 | 0.99 | 0.99 |
| SL | Cardiac | 21721 | 113 | Sensitivity | 0.87 | 0.58 | 0.31 | 0.14 | 0.05 | 0.02 | +0 | +0 | +0 |
| SL | Cardiac | 21721 | 113 | Specificity | 0.47 | 0.82 | 0.97 | 0.99 | 0.99 | 1 | 1 | 1 | 1 |
| SL | Cardiac | 21721 | 113 | Alert rate | 0.53 | 0.18 | 0.03 | 0.01 | 0.01 | +0 | +0 | +0 | +0 |
| SL | Cardiac | 21721 | 113 | PPV | 0.01 | 0.02 | 0.05 | 0.05 | 0.03 | 0.02 | NaN | NaN | NaN |
| SL | Cardiac | 21721 | 113 | NPV | 1 | 1 | 1 | 1 | 0.99 | 0.99 | 0.99 | 0.99 | 0.99 |
| XGB-ALL | Cardiac | 21721 | 113 | Sensitivity | 0.77 | 0.5 | 0.28 | 0.15 | 0.06 | 0.04 | +0 | +0 | +0 |
| XGB-ALL | Cardiac | 21721 | 113 | Specificity | 0.66 | 0.89 | 0.97 | 0.99 | 0.99 | 0.99 | 1 | 1 | 1 |
| XGB-ALL | Cardiac | 21721 | 113 | Alert rate | 0.34 | 0.11 | 0.03 | 0.01 | 0.01 | 0.01 | +0 | +0 | +0 |
| XGB-ALL | Cardiac | 21721 | 113 | PPV | 0.01 | 0.02 | 0.05 | 0.06 | 0.04 | 0.03 | +0 | NaN | NaN |
| XGB-ALL | Cardiac | 21721 | 113 | NPV | 1 | 1 | 1 | 1 | 0.99 | 0.99 | 0.99 | 0.99 | 0.99 |
| XGB-LIM | Cardiac | 21721 | 113 | Sensitivity | 0.9 | 0.52 | 0.25 | 0.11 | 0.04 | +0 | +0 | +0 | +0 |
| XGB-LIM | Cardiac | 21721 | 113 | Specificity | 0.51 | 0.82 | 0.96 | 0.98 | 0.99 | 0.99 | 1 | 1 | 1 |
| XGB-LIM | Cardiac | 21721 | 113 | Alert rate | 0.49 | 0.18 | 0.04 | 0.02 | 0.01 | 0.01 | +0 | +0 | +0 |
| XGB-LIM | Cardiac | 21721 | 113 | PPV | 0.01 | 0.02 | 0.03 | 0.03 | 0.02 | +0 | NaN | NaN | NaN |
| XGB-LIM | Cardiac | 21721 | 113 | NPV | 1 | 1 | 1 | 1 | 0.99 | 0.99 | 0.99 | 0.99 | 0.99 |
| CS-LIM | UNKNOWN_VALUE | 19808 | 195 | Sensitivity | 0.99 | 0.8 | 0.55 | 0.27 | 0.16 | 0.08 | +0 | +0 | +0 |
| CS-LIM | UNKNOWN_VALUE | 19808 | 195 | Specificity | 0.09 | 0.37 | 0.78 | 0.91 | 0.96 | 0.98 | 1 | 1 | 1 |
| CS-LIM | UNKNOWN_VALUE | 19808 | 195 | Alert rate | 0.91 | 0.63 | 0.22 | 0.09 | 0.04 | 0.02 | +0 | +0 | +0 |
| CS-LIM | UNKNOWN_VALUE | 19808 | 195 | PPV | 0.01 | 0.01 | 0.02 | 0.02 | 0.03 | 0.03 | +0 | NaN | NaN |
| CS-LIM | UNKNOWN_VALUE | 19808 | 195 | NPV | 1 | 1 | 1 | 0.99 | 0.99 | 0.99 | 0.99 | 0.99 | 0.99 |
| RF-ALL | UNKNOWN_VALUE | 19808 | 195 | Sensitivity | 1 | 0.98 | 0.65 | 0.34 | 0.01 | +0 | +0 | +0 | +0 |
| RF-ALL | UNKNOWN_VALUE | 19808 | 195 | Specificity | 0.04 | 0.17 | 0.73 | 0.93 | 0.99 | 1 | 1 | 1 | 1 |
| RF-ALL | UNKNOWN_VALUE | 19808 | 195 | Alert rate | 0.97 | 0.83 | 0.27 | 0.08 | 0.01 | +0 | +0 | +0 | +0 |
| RF-ALL | UNKNOWN_VALUE | 19808 | 195 | PPV | 0.01 | 0.01 | 0.02 | 0.03 | 0.01 | +0 | NaN | NaN | NaN |
| RF-ALL | UNKNOWN_VALUE | 19808 | 195 | NPV | 1 | 1 | 1 | 0.99 | 0.99 | 0.99 | 0.99 | 0.99 | 0.99 |
| RF-LIM | UNKNOWN_VALUE | 19808 | 195 | Sensitivity | 1 | 0.99 | 0.79 | 0.5 | 0.17 | 0.06 | +0 | +0 | +0 |
| RF-LIM | UNKNOWN_VALUE | 19808 | 195 | Specificity | 0.04 | 0.14 | 0.54 | 0.85 | 0.97 | 0.99 | 1 | 1 | 1 |
| RF-LIM | UNKNOWN_VALUE | 19808 | 195 | Alert rate | 0.96 | 0.86 | 0.47 | 0.15 | 0.03 | 0.01 | +0 | +0 | +0 |
| RF-LIM | UNKNOWN_VALUE | 19808 | 195 | PPV | 0.01 | 0.01 | 0.01 | 0.03 | 0.04 | 0.05 | NaN | NaN | NaN |
| RF-LIM | UNKNOWN_VALUE | 19808 | 195 | NPV | 1 | 1 | 1 | 1 | 0.99 | 0.99 | 0.99 | 0.99 | 0.99 |
| SL | UNKNOWN_VALUE | 19808 | 195 | Sensitivity | 1 | 0.96 | 0.65 | 0.27 | 0.14 | 0.08 | +0 | +0 | +0 |
| SL | UNKNOWN_VALUE | 19808 | 195 | Specificity | 0.07 | 0.33 | 0.82 | 0.94 | 0.98 | 0.99 | 1 | 1 | 1 |
| SL | UNKNOWN_VALUE | 19808 | 195 | Alert rate | 0.93 | 0.67 | 0.19 | 0.06 | 0.02 | 0.01 | +0 | +0 | +0 |
| SL | UNKNOWN_VALUE | 19808 | 195 | PPV | 0.01 | 0.01 | 0.03 | 0.03 | 0.04 | 0.06 | NaN | NaN | NaN |
| SL | UNKNOWN_VALUE | 19808 | 195 | NPV | 1 | 1 | 1 | 0.99 | 0.99 | 0.99 | 0.99 | 0.99 | 0.99 |
| XGB-ALL | UNKNOWN_VALUE | 19808 | 195 | Sensitivity | 0.98 | 0.94 | 0.59 | 0.26 | 0.17 | 0.11 | 0.01 | +0 | +0 |
| XGB-ALL | UNKNOWN_VALUE | 19808 | 195 | Specificity | 0.16 | 0.47 | 0.84 | 0.94 | 0.97 | 0.98 | 1 | 1 | 1 |
| XGB-ALL | UNKNOWN_VALUE | 19808 | 195 | Alert rate | 0.85 | 0.53 | 0.16 | 0.06 | 0.03 | 0.02 | +0 | +0 | +0 |
| XGB-ALL | UNKNOWN_VALUE | 19808 | 195 | PPV | 0.01 | 0.01 | 0.03 | 0.03 | 0.05 | 0.05 | 0.33 | NaN | NaN |
| XGB-ALL | UNKNOWN_VALUE | 19808 | 195 | NPV | 1 | 1 | 1 | 0.99 | 0.99 | 0.99 | 0.99 | 0.99 | 0.99 |
| XGB-LIM | UNKNOWN_VALUE | 19808 | 195 | Sensitivity | 1 | 0.94 | 0.64 | 0.3 | 0.21 | 0.13 | 0.01 | +0 | +0 |
| XGB-LIM | UNKNOWN_VALUE | 19808 | 195 | Specificity | 0.11 | 0.4 | 0.79 | 0.92 | 0.96 | 0.98 | 1 | 1 | 1 |
| XGB-LIM | UNKNOWN_VALUE | 19808 | 195 | Alert rate | 0.89 | 0.61 | 0.21 | 0.08 | 0.04 | 0.02 | +0 | +0 | +0 |
| XGB-LIM | UNKNOWN_VALUE | 19808 | 195 | PPV | 0.01 | 0.01 | 0.02 | 0.03 | 0.04 | 0.04 | 0.33 | NaN | NaN |
| XGB-LIM | UNKNOWN_VALUE | 19808 | 195 | NPV | 1 | 1 | 1 | 0.99 | 0.99 | 0.99 | 0.99 | 0.99 | 0.99 |
| CS-LIM | Gynecology | 16706 | 113 | Sensitivity | 1 | 0.86 | 0.46 | 0.26 | 0.18 | 0.05 | +0 | +0 | +0 |
| CS-LIM | Gynecology | 16706 | 113 | Specificity | 0.14 | 0.44 | 0.87 | 0.96 | 0.99 | 1 | 1 | 1 | 1 |
| CS-LIM | Gynecology | 16706 | 113 | Alert rate | 0.86 | 0.56 | 0.13 | 0.04 | 0.02 | +0 | +0 | +0 | +0 |
| CS-LIM | Gynecology | 16706 | 113 | PPV | 0.01 | 0.01 | 0.02 | 0.04 | 0.07 | 0.08 | NaN | NaN | NaN |
| CS-LIM | Gynecology | 16706 | 113 | NPV | 1 | 1 | 1 | 1 | 0.99 | 0.99 | 0.99 | 0.99 | 0.99 |
| RF-ALL | Gynecology | 16706 | 113 | Sensitivity | 1 | 0.98 | 0.42 | 0.24 | 0.08 | +0 | +0 | +0 | +0 |
| RF-ALL | Gynecology | 16706 | 113 | Specificity | 0.24 | 0.52 | 0.9 | 0.97 | 1 | 1 | 1 | 1 | 1 |
| RF-ALL | Gynecology | 16706 | 113 | Alert rate | 0.77 | 0.48 | 0.1 | 0.03 | +0 | +0 | +0 | +0 | +0 |
| RF-ALL | Gynecology | 16706 | 113 | PPV | 0.01 | 0.01 | 0.03 | 0.05 | 0.11 | +0 | NaN | NaN | NaN |
| RF-ALL | Gynecology | 16706 | 113 | NPV | 1 | 1 | 1 | 1 | 0.99 | 0.99 | 0.99 | 0.99 | 0.99 |
| RF-LIM | Gynecology | 16706 | 113 | Sensitivity | 1 | 0.98 | 0.52 | 0.34 | 0.18 | 0.01 | +0 | +0 | +0 |
| RF-LIM | Gynecology | 16706 | 113 | Specificity | 0.16 | 0.43 | 0.82 | 0.95 | 0.98 | 1 | 1 | 1 | 1 |
| RF-LIM | Gynecology | 16706 | 113 | Alert rate | 0.84 | 0.57 | 0.18 | 0.05 | 0.02 | +0 | +0 | +0 | +0 |
| RF-LIM | Gynecology | 16706 | 113 | PPV | 0.01 | 0.01 | 0.02 | 0.04 | 0.07 | 0.05 | NaN | NaN | NaN |
| RF-LIM | Gynecology | 16706 | 113 | NPV | 1 | 1 | 1 | 1 | 0.99 | 0.99 | 0.99 | 0.99 | 0.99 |
| SL | Gynecology | 16706 | 113 | Sensitivity | 1 | 0.89 | 0.44 | 0.27 | 0.09 | 0.04 | 0.01 | +0 | +0 |
| SL | Gynecology | 16706 | 113 | Specificity | 0.35 | 0.65 | 0.92 | 0.98 | 0.99 | 1 | 1 | 1 | 1 |
| SL | Gynecology | 16706 | 113 | Alert rate | 0.65 | 0.35 | 0.08 | 0.02 | 0.01 | +0 | +0 | +0 | +0 |
| SL | Gynecology | 16706 | 113 | PPV | 0.01 | 0.02 | 0.03 | 0.08 | 0.08 | 0.11 | 1 | NaN | NaN |
| SL | Gynecology | 16706 | 113 | NPV | 1 | 1 | 1 | 1 | 0.99 | 0.99 | 0.99 | 0.99 | 0.99 |
| XGB-ALL | Gynecology | 16706 | 113 | Sensitivity | 0.96 | 0.81 | 0.43 | 0.26 | 0.11 | 0.05 | 0.01 | 0.01 | +0 |
| XGB-ALL | Gynecology | 16706 | 113 | Specificity | 0.46 | 0.72 | 0.93 | 0.98 | 0.99 | 1 | 1 | 1 | 1 |
| XGB-ALL | Gynecology | 16706 | 113 | Alert rate | 0.54 | 0.28 | 0.07 | 0.02 | 0.01 | +0 | +0 | +0 | +0 |
| XGB-ALL | Gynecology | 16706 | 113 | PPV | 0.01 | 0.02 | 0.04 | 0.07 | 0.08 | 0.07 | 1 | 1 | NaN |
| XGB-ALL | Gynecology | 16706 | 113 | NPV | 1 | 1 | 1 | 1 | 0.99 | 0.99 | 0.99 | 0.99 | 0.99 |
| XGB-LIM | Gynecology | 16706 | 113 | Sensitivity | 0.99 | 0.88 | 0.47 | 0.28 | 0.16 | 0.08 | +0 | +0 | +0 |
| XGB-LIM | Gynecology | 16706 | 113 | Specificity | 0.33 | 0.6 | 0.89 | 0.96 | 0.98 | 0.99 | 1 | 1 | 1 |
| XGB-LIM | Gynecology | 16706 | 113 | Alert rate | 0.67 | 0.4 | 0.12 | 0.04 | 0.02 | 0.01 | +0 | +0 | +0 |
| XGB-LIM | Gynecology | 16706 | 113 | PPV | 0.01 | 0.01 | 0.02 | 0.05 | 0.06 | 0.06 | NaN | NaN | NaN |
| XGB-LIM | Gynecology | 16706 | 113 | NPV | 1 | 1 | 1 | 1 | 0.99 | 0.99 | 0.99 | 0.99 | 0.99 |
| CS-LIM | Neonatology | 12672 | 274 | Sensitivity | 1 | 1 | 0.98 | 0.93 | 0.63 | 0.26 | +0 | +0 | +0 |
| CS-LIM | Neonatology | 12672 | 274 | Specificity | +0 | 0.01 | 0.06 | 0.2 | 0.57 | 0.84 | 1 | 1 | 1 |
| CS-LIM | Neonatology | 12672 | 274 | Alert rate | 1 | 0.99 | 0.94 | 0.8 | 0.44 | 0.16 | +0 | +0 | +0 |
| CS-LIM | Neonatology | 12672 | 274 | PPV | 0.02 | 0.02 | 0.02 | 0.03 | 0.03 | 0.03 | +0 | +0 | +0 |
| CS-LIM | Neonatology | 12672 | 274 | NPV | 1 | 1 | 0.99 | 0.99 | 0.99 | 0.98 | 0.98 | 0.98 | 0.98 |
| RF-ALL | Neonatology | 12672 | 274 | Sensitivity | 1 | 0.99 | 0.97 | 0.82 | 0.61 | 0.41 | +0 | +0 | +0 |
| RF-ALL | Neonatology | 12672 | 274 | Specificity | 0.04 | 0.07 | 0.2 | 0.49 | 0.74 | 0.88 | 1 | 1 | 1 |
| RF-ALL | Neonatology | 12672 | 274 | Alert rate | 0.96 | 0.93 | 0.8 | 0.51 | 0.27 | 0.12 | +0 | +0 | +0 |
| RF-ALL | Neonatology | 12672 | 274 | PPV | 0.02 | 0.02 | 0.03 | 0.03 | 0.05 | 0.07 | NaN | NaN | NaN |
| RF-ALL | Neonatology | 12672 | 274 | NPV | 1 | 1 | 1 | 0.99 | 0.99 | 0.99 | 0.98 | 0.98 | 0.98 |
| RF-LIM | Neonatology | 12672 | 274 | Sensitivity | 1 | 0.99 | 0.96 | 0.81 | 0.59 | 0.36 | +0 | +0 | +0 |
| RF-LIM | Neonatology | 12672 | 274 | Specificity | 0.04 | 0.07 | 0.18 | 0.46 | 0.73 | 0.9 | 1 | 1 | 1 |
| RF-LIM | Neonatology | 12672 | 274 | Alert rate | 0.96 | 0.93 | 0.82 | 0.54 | 0.27 | 0.11 | +0 | +0 | +0 |
| RF-LIM | Neonatology | 12672 | 274 | PPV | 0.02 | 0.02 | 0.03 | 0.03 | 0.05 | 0.07 | NaN | NaN | NaN |
| RF-LIM | Neonatology | 12672 | 274 | NPV | 1 | 1 | 1 | 0.99 | 0.99 | 0.98 | 0.98 | 0.98 | 0.98 |
| SL | Neonatology | 12672 | 274 | Sensitivity | 1 | 0.99 | 0.9 | 0.73 | 0.56 | 0.42 | 0.01 | +0 | +0 |
| SL | Neonatology | 12672 | 274 | Specificity | 0.05 | 0.1 | 0.4 | 0.64 | 0.79 | 0.87 | 1 | 1 | 1 |
| SL | Neonatology | 12672 | 274 | Alert rate | 0.95 | 0.9 | 0.61 | 0.37 | 0.22 | 0.13 | +0 | +0 | +0 |
| SL | Neonatology | 12672 | 274 | PPV | 0.02 | 0.02 | 0.03 | 0.04 | 0.06 | 0.07 | 0.11 | NaN | NaN |
| SL | Neonatology | 12672 | 274 | NPV | 1 | 1 | 0.99 | 0.99 | 0.99 | 0.99 | 0.98 | 0.98 | 0.98 |
| XGB-ALL | Neonatology | 12672 | 274 | Sensitivity | 0.99 | 0.97 | 0.85 | 0.67 | 0.55 | 0.43 | 0.05 | +0 | +0 |
| XGB-ALL | Neonatology | 12672 | 274 | Specificity | 0.07 | 0.23 | 0.51 | 0.69 | 0.8 | 0.87 | 1 | 1 | 1 |
| XGB-ALL | Neonatology | 12672 | 274 | Alert rate | 0.93 | 0.78 | 0.5 | 0.32 | 0.21 | 0.14 | 0.01 | +0 | +0 |
| XGB-ALL | Neonatology | 12672 | 274 | PPV | 0.02 | 0.03 | 0.04 | 0.05 | 0.06 | 0.07 | 0.2 | 0.09 | NaN |
| XGB-ALL | Neonatology | 12672 | 274 | NPV | 1 | 1 | 0.99 | 0.99 | 0.99 | 0.99 | 0.98 | 0.98 | 0.98 |
| XGB-LIM | Neonatology | 12672 | 274 | Sensitivity | 1 | 0.96 | 0.82 | 0.67 | 0.52 | 0.42 | 0.04 | 0.01 | +0 |
| XGB-LIM | Neonatology | 12672 | 274 | Specificity | 0.07 | 0.2 | 0.48 | 0.66 | 0.78 | 0.85 | 1 | 1 | 1 |
| XGB-LIM | Neonatology | 12672 | 274 | Alert rate | 0.93 | 0.8 | 0.53 | 0.34 | 0.23 | 0.15 | 0.01 | +0 | +0 |
| XGB-LIM | Neonatology | 12672 | 274 | PPV | 0.02 | 0.03 | 0.03 | 0.04 | 0.05 | 0.06 | 0.16 | 0.17 | NaN |
| XGB-LIM | Neonatology | 12672 | 274 | NPV | 1 | 1 | 0.99 | 0.99 | 0.99 | 0.99 | 0.98 | 0.98 | 0.98 |
| CS-LIM | Thoracic Surgery | 10985 | 134 | Sensitivity | 0.99 | 0.9 | 0.62 | 0.27 | 0.08 | 0.03 | +0 | +0 | +0 |
| CS-LIM | Thoracic Surgery | 10985 | 134 | Specificity | 0.11 | 0.45 | 0.83 | 0.92 | 0.97 | 0.99 | 1 | 1 | 1 |
| CS-LIM | Thoracic Surgery | 10985 | 134 | Alert rate | 0.89 | 0.55 | 0.17 | 0.09 | 0.03 | 0.01 | +0 | +0 | +0 |
| CS-LIM | Thoracic Surgery | 10985 | 134 | PPV | 0.01 | 0.02 | 0.04 | 0.04 | 0.04 | 0.04 | NaN | NaN | NaN |
| CS-LIM | Thoracic Surgery | 10985 | 134 | NPV | 1 | 1 | 0.99 | 0.99 | 0.99 | 0.99 | 0.99 | 0.99 | 0.99 |
| RF-ALL | Thoracic Surgery | 10985 | 134 | Sensitivity | 1 | 0.97 | 0.69 | 0.42 | 0.15 | 0.02 | +0 | +0 | +0 |
| RF-ALL | Thoracic Surgery | 10985 | 134 | Specificity | 0.1 | 0.3 | 0.8 | 0.9 | 0.98 | 1 | 1 | 1 | 1 |
| RF-ALL | Thoracic Surgery | 10985 | 134 | Alert rate | 0.9 | 0.7 | 0.21 | 0.1 | 0.02 | +0 | +0 | +0 | +0 |
| RF-ALL | Thoracic Surgery | 10985 | 134 | PPV | 0.01 | 0.02 | 0.04 | 0.05 | 0.08 | 0.06 | NaN | NaN | NaN |
| RF-ALL | Thoracic Surgery | 10985 | 134 | NPV | 1 | 1 | 1 | 0.99 | 0.99 | 0.99 | 0.99 | 0.99 | 0.99 |
| RF-LIM | Thoracic Surgery | 10985 | 134 | Sensitivity | 1 | 0.95 | 0.73 | 0.48 | 0.19 | 0.05 | +0 | +0 | +0 |
| RF-LIM | Thoracic Surgery | 10985 | 134 | Specificity | 0.1 | 0.27 | 0.76 | 0.88 | 0.97 | 1 | 1 | 1 | 1 |
| RF-LIM | Thoracic Surgery | 10985 | 134 | Alert rate | 0.9 | 0.74 | 0.25 | 0.13 | 0.03 | +0 | +0 | +0 | +0 |
| RF-LIM | Thoracic Surgery | 10985 | 134 | PPV | 0.01 | 0.02 | 0.03 | 0.04 | 0.06 | 0.12 | NaN | NaN | NaN |
| RF-LIM | Thoracic Surgery | 10985 | 134 | NPV | 1 | 1 | 1 | 0.99 | 0.99 | 0.99 | 0.99 | 0.99 | 0.99 |
| SL | Thoracic Surgery | 10985 | 134 | Sensitivity | 0.99 | 0.9 | 0.66 | 0.44 | 0.22 | 0.13 | +0 | +0 | +0 |
| SL | Thoracic Surgery | 10985 | 134 | Specificity | 0.12 | 0.48 | 0.83 | 0.92 | 0.96 | 0.98 | 1 | 1 | 1 |
| SL | Thoracic Surgery | 10985 | 134 | Alert rate | 0.88 | 0.53 | 0.17 | 0.09 | 0.04 | 0.02 | +0 | +0 | +0 |
| SL | Thoracic Surgery | 10985 | 134 | PPV | 0.01 | 0.02 | 0.05 | 0.06 | 0.06 | 0.08 | NaN | NaN | NaN |
| SL | Thoracic Surgery | 10985 | 134 | NPV | 1 | 1 | 1 | 0.99 | 0.99 | 0.99 | 0.99 | 0.99 | 0.99 |
| XGB-ALL | Thoracic Surgery | 10985 | 134 | Sensitivity | 0.98 | 0.9 | 0.6 | 0.4 | 0.24 | 0.19 | +0 | +0 | +0 |
| XGB-ALL | Thoracic Surgery | 10985 | 134 | Specificity | 0.21 | 0.56 | 0.85 | 0.92 | 0.96 | 0.97 | 1 | 1 | 1 |
| XGB-ALL | Thoracic Surgery | 10985 | 134 | Alert rate | 0.79 | 0.44 | 0.16 | 0.09 | 0.05 | 0.03 | +0 | +0 | +0 |
| XGB-ALL | Thoracic Surgery | 10985 | 134 | PPV | 0.01 | 0.02 | 0.04 | 0.05 | 0.06 | 0.08 | +0 | NaN | NaN |
| XGB-ALL | Thoracic Surgery | 10985 | 134 | NPV | 1 | 1 | 0.99 | 0.99 | 0.99 | 0.99 | 0.99 | 0.99 | 0.99 |
| XGB-LIM | Thoracic Surgery | 10985 | 134 | Sensitivity | 0.99 | 0.85 | 0.65 | 0.41 | 0.24 | 0.12 | +0 | +0 | +0 |
| XGB-LIM | Thoracic Surgery | 10985 | 134 | Specificity | 0.16 | 0.5 | 0.83 | 0.9 | 0.95 | 0.98 | 1 | 1 | 1 |
| XGB-LIM | Thoracic Surgery | 10985 | 134 | Alert rate | 0.85 | 0.51 | 0.18 | 0.1 | 0.05 | 0.02 | +0 | +0 | +0 |
| XGB-LIM | Thoracic Surgery | 10985 | 134 | PPV | 0.01 | 0.02 | 0.04 | 0.05 | 0.05 | 0.06 | +0 | NaN | NaN |
| XGB-LIM | Thoracic Surgery | 10985 | 134 | NPV | 1 | 1 | 0.99 | 0.99 | 0.99 | 0.99 | 0.99 | 0.99 | 0.99 |
| CS-LIM | Geriatrics | 9906 | 192 | Sensitivity | 0.99 | 0.97 | 0.6 | 0.46 | 0.36 | 0.21 | +0 | +0 | +0 |
| CS-LIM | Geriatrics | 9906 | 192 | Specificity | +0 | 0.08 | 0.69 | 0.89 | 0.94 | 0.97 | 1 | 1 | 1 |
| CS-LIM | Geriatrics | 9906 | 192 | Alert rate | 1 | 0.92 | 0.31 | 0.12 | 0.07 | 0.04 | +0 | +0 | +0 |
| CS-LIM | Geriatrics | 9906 | 192 | PPV | 0.02 | 0.02 | 0.04 | 0.08 | 0.1 | 0.11 | +0 | NaN | NaN |
| CS-LIM | Geriatrics | 9906 | 192 | NPV | 0.95 | 0.99 | 0.99 | 0.99 | 0.99 | 0.98 | 0.98 | 0.98 | 0.98 |
| RF-ALL | Geriatrics | 9906 | 192 | Sensitivity | 1 | 0.99 | 0.82 | 0.5 | 0.16 | 0.05 | +0 | +0 | +0 |
| RF-ALL | Geriatrics | 9906 | 192 | Specificity | 0.01 | 0.04 | 0.44 | 0.9 | 0.98 | 1 | 1 | 1 | 1 |
| RF-ALL | Geriatrics | 9906 | 192 | Alert rate | 0.99 | 0.96 | 0.56 | 0.11 | 0.02 | +0 | +0 | +0 | +0 |
| RF-ALL | Geriatrics | 9906 | 192 | PPV | 0.02 | 0.02 | 0.03 | 0.09 | 0.17 | 0.38 | NaN | NaN | NaN |
| RF-ALL | Geriatrics | 9906 | 192 | NPV | 1 | 1 | 0.99 | 0.99 | 0.98 | 0.98 | 0.98 | 0.98 | 0.98 |
| RF-LIM | Geriatrics | 9906 | 192 | Sensitivity | 0.99 | 0.99 | 0.76 | 0.47 | 0.2 | 0.03 | +0 | +0 | +0 |
| RF-LIM | Geriatrics | 9906 | 192 | Specificity | 0.02 | 0.05 | 0.58 | 0.9 | 0.98 | 1 | 1 | 1 | 1 |
| RF-LIM | Geriatrics | 9906 | 192 | Alert rate | 0.98 | 0.95 | 0.43 | 0.11 | 0.03 | +0 | +0 | +0 | +0 |
| RF-LIM | Geriatrics | 9906 | 192 | PPV | 0.02 | 0.02 | 0.03 | 0.09 | 0.15 | 0.24 | NaN | NaN | NaN |
| RF-LIM | Geriatrics | 9906 | 192 | NPV | 0.99 | 1 | 0.99 | 0.99 | 0.98 | 0.98 | 0.98 | 0.98 | 0.98 |
| SL | Geriatrics | 9906 | 192 | Sensitivity | 0.99 | 0.98 | 0.65 | 0.46 | 0.34 | 0.25 | +0 | +0 | +0 |
| SL | Geriatrics | 9906 | 192 | Specificity | 0.02 | 0.12 | 0.76 | 0.9 | 0.94 | 0.97 | 1 | 1 | 1 |
| SL | Geriatrics | 9906 | 192 | Alert rate | 0.98 | 0.88 | 0.25 | 0.11 | 0.06 | 0.04 | +0 | +0 | +0 |
| SL | Geriatrics | 9906 | 192 | PPV | 0.02 | 0.02 | 0.05 | 0.08 | 0.11 | 0.14 | +0 | NaN | NaN |
| SL | Geriatrics | 9906 | 192 | NPV | 0.99 | 1 | 0.99 | 0.99 | 0.99 | 0.98 | 0.98 | 0.98 | 0.98 |
| XGB-ALL | Geriatrics | 9906 | 192 | Sensitivity | 0.99 | 0.87 | 0.56 | 0.46 | 0.33 | 0.29 | 0.01 | +0 | +0 |
| XGB-ALL | Geriatrics | 9906 | 192 | Specificity | 0.05 | 0.35 | 0.79 | 0.89 | 0.93 | 0.96 | 1 | 1 | 1 |
| XGB-ALL | Geriatrics | 9906 | 192 | Alert rate | 0.95 | 0.66 | 0.22 | 0.11 | 0.07 | 0.05 | +0 | +0 | +0 |
| XGB-ALL | Geriatrics | 9906 | 192 | PPV | 0.02 | 0.03 | 0.05 | 0.08 | 0.09 | 0.12 | 0.06 | +0 | NaN |
| XGB-ALL | Geriatrics | 9906 | 192 | NPV | 1 | 0.99 | 0.99 | 0.99 | 0.99 | 0.99 | 0.98 | 0.98 | 0.98 |
| XGB-LIM | Geriatrics | 9906 | 192 | Sensitivity | 0.99 | 0.91 | 0.62 | 0.45 | 0.37 | 0.23 | +0 | +0 | +0 |
| XGB-LIM | Geriatrics | 9906 | 192 | Specificity | 0.04 | 0.28 | 0.76 | 0.89 | 0.95 | 0.97 | 1 | 1 | 1 |
| XGB-LIM | Geriatrics | 9906 | 192 | Alert rate | 0.96 | 0.72 | 0.25 | 0.12 | 0.06 | 0.03 | +0 | +0 | +0 |
| XGB-LIM | Geriatrics | 9906 | 192 | PPV | 0.02 | 0.03 | 0.05 | 0.08 | 0.12 | 0.13 | +0 | +0 | NaN |
| XGB-LIM | Geriatrics | 9906 | 192 | NPV | 1 | 0.99 | 0.99 | 0.99 | 0.99 | 0.98 | 0.98 | 0.98 | 0.98 |
| CS-LIM | Transplant | 9277 | 70 | Sensitivity | 1 | 0.82 | 0.51 | 0.32 | 0.18 | 0.11 | +0 | +0 | +0 |
| CS-LIM | Transplant | 9277 | 70 | Specificity | 0.03 | 0.24 | 0.8 | 0.92 | 0.96 | 0.99 | 1 | 1 | 1 |
| CS-LIM | Transplant | 9277 | 70 | Alert rate | 0.97 | 0.76 | 0.2 | 0.08 | 0.04 | 0.01 | +0 | +0 | +0 |
| CS-LIM | Transplant | 9277 | 70 | PPV | 0.01 | 0.01 | 0.02 | 0.03 | 0.04 | 0.06 | NaN | NaN | NaN |
| CS-LIM | Transplant | 9277 | 70 | NPV | 1 | 0.99 | 1 | 0.99 | 0.99 | 0.99 | 0.99 | 0.99 | 0.99 |
| RF-ALL | Transplant | 9277 | 70 | Sensitivity | 1 | 0.89 | 0.54 | 0.32 | 0.06 | +0 | +0 | +0 | +0 |
| RF-ALL | Transplant | 9277 | 70 | Specificity | 0.11 | 0.42 | 0.84 | 0.94 | 0.98 | 1 | 1 | 1 | 1 |
| RF-ALL | Transplant | 9277 | 70 | Alert rate | 0.89 | 0.59 | 0.16 | 0.06 | 0.02 | +0 | +0 | +0 | +0 |
| RF-ALL | Transplant | 9277 | 70 | PPV | 0.01 | 0.01 | 0.03 | 0.04 | 0.02 | +0 | NaN | NaN | NaN |
| RF-ALL | Transplant | 9277 | 70 | NPV | 1 | 1 | 1 | 0.99 | 0.99 | 0.99 | 0.99 | 0.99 | 0.99 |
| RF-LIM | Transplant | 9277 | 70 | Sensitivity | 1 | 0.98 | 0.57 | 0.46 | 0.18 | 0.02 | +0 | +0 | +0 |
| RF-LIM | Transplant | 9277 | 70 | Specificity | 0.09 | 0.33 | 0.75 | 0.9 | 0.96 | 0.99 | 1 | 1 | 1 |
| RF-LIM | Transplant | 9277 | 70 | Alert rate | 0.91 | 0.68 | 0.25 | 0.11 | 0.05 | 0.01 | +0 | +0 | +0 |
| RF-LIM | Transplant | 9277 | 70 | PPV | 0.01 | 0.01 | 0.02 | 0.03 | 0.03 | 0.01 | NaN | NaN | NaN |
| RF-LIM | Transplant | 9277 | 70 | NPV | 1 | 1 | 1 | 1 | 0.99 | 0.99 | 0.99 | 0.99 | 0.99 |
| SL | Transplant | 9277 | 70 | Sensitivity | 1 | 0.86 | 0.54 | 0.35 | 0.25 | 0.05 | +0 | +0 | +0 |
| SL | Transplant | 9277 | 70 | Specificity | 0.16 | 0.54 | 0.88 | 0.95 | 0.97 | 0.99 | 1 | 1 | 1 |
| SL | Transplant | 9277 | 70 | Alert rate | 0.84 | 0.46 | 0.13 | 0.05 | 0.03 | 0.02 | +0 | +0 | +0 |
| SL | Transplant | 9277 | 70 | PPV | 0.01 | 0.01 | 0.03 | 0.05 | 0.07 | 0.02 | +0 | NaN | NaN |
| SL | Transplant | 9277 | 70 | NPV | 1 | 1 | 1 | 0.99 | 0.99 | 0.99 | 0.99 | 0.99 | 0.99 |
| XGB-ALL | Transplant | 9277 | 70 | Sensitivity | 0.95 | 0.82 | 0.55 | 0.38 | 0.26 | 0.09 | +0 | +0 | +0 |
| XGB-ALL | Transplant | 9277 | 70 | Specificity | 0.26 | 0.65 | 0.89 | 0.95 | 0.97 | 0.98 | 1 | 1 | 1 |
| XGB-ALL | Transplant | 9277 | 70 | Alert rate | 0.74 | 0.36 | 0.12 | 0.05 | 0.03 | 0.02 | +0 | +0 | +0 |
| XGB-ALL | Transplant | 9277 | 70 | PPV | 0.01 | 0.02 | 0.04 | 0.06 | 0.07 | 0.04 | +0 | +0 | NaN |
| XGB-ALL | Transplant | 9277 | 70 | NPV | 1 | 1 | 1 | 0.99 | 0.99 | 0.99 | 0.99 | 0.99 | 0.99 |
| XGB-LIM | Transplant | 9277 | 70 | Sensitivity | 1 | 0.88 | 0.62 | 0.49 | 0.28 | 0.06 | +0 | +0 | +0 |
| XGB-LIM | Transplant | 9277 | 70 | Specificity | 0.16 | 0.49 | 0.82 | 0.92 | 0.96 | 0.98 | 1 | 1 | 1 |
| XGB-LIM | Transplant | 9277 | 70 | Alert rate | 0.84 | 0.51 | 0.18 | 0.09 | 0.04 | 0.02 | +0 | +0 | +0 |
| XGB-LIM | Transplant | 9277 | 70 | PPV | 0.01 | 0.01 | 0.03 | 0.05 | 0.05 | 0.02 | NaN | NaN | NaN |
| XGB-LIM | Transplant | 9277 | 70 | NPV | 1 | 1 | 1 | 1 | 0.99 | 0.99 | 0.99 | 0.99 | 0.99 |
| CS-LIM | Neuro | 8227 | 131 | Sensitivity | 0.98 | 0.83 | 0.37 | 0.22 | 0.11 | 0.07 | +0 | +0 | +0 |
| CS-LIM | Neuro | 8227 | 131 | Specificity | 0.18 | 0.51 | 0.86 | 0.94 | 0.98 | 0.99 | 1 | 1 | 1 |
| CS-LIM | Neuro | 8227 | 131 | Alert rate | 0.82 | 0.5 | 0.15 | 0.06 | 0.03 | 0.01 | +0 | +0 | +0 |
| CS-LIM | Neuro | 8227 | 131 | PPV | 0.02 | 0.03 | 0.04 | 0.06 | 0.07 | 0.09 | NaN | NaN | NaN |
| CS-LIM | Neuro | 8227 | 131 | NPV | 1 | 0.99 | 0.99 | 0.99 | 0.99 | 0.99 | 0.98 | 0.98 | 0.98 |
| RF-ALL | Neuro | 8227 | 131 | Sensitivity | 0.98 | 0.86 | 0.25 | 0.11 | 0.03 | 0.01 | +0 | +0 | +0 |
| RF-ALL | Neuro | 8227 | 131 | Specificity | 0.25 | 0.53 | 0.91 | 0.98 | 0.99 | 1 | 1 | 1 | 1 |
| RF-ALL | Neuro | 8227 | 131 | Alert rate | 0.75 | 0.48 | 0.09 | 0.02 | 0.01 | +0 | +0 | +0 | +0 |
| RF-ALL | Neuro | 8227 | 131 | PPV | 0.02 | 0.03 | 0.04 | 0.07 | 0.07 | 0.09 | NaN | NaN | NaN |
| RF-ALL | Neuro | 8227 | 131 | NPV | 1 | 1 | 0.99 | 0.99 | 0.98 | 0.98 | 0.98 | 0.98 | 0.98 |
| RF-LIM | Neuro | 8227 | 131 | Sensitivity | 0.98 | 0.89 | 0.39 | 0.17 | 0.08 | 0.02 | +0 | +0 | +0 |
| RF-LIM | Neuro | 8227 | 131 | Specificity | 0.23 | 0.48 | 0.85 | 0.95 | 0.98 | 0.99 | 1 | 1 | 1 |
| RF-LIM | Neuro | 8227 | 131 | Alert rate | 0.78 | 0.52 | 0.15 | 0.05 | 0.02 | 0.01 | +0 | +0 | +0 |
| RF-LIM | Neuro | 8227 | 131 | PPV | 0.02 | 0.03 | 0.04 | 0.05 | 0.06 | 0.07 | +0 | NaN | NaN |
| RF-LIM | Neuro | 8227 | 131 | NPV | 1 | 1 | 0.99 | 0.99 | 0.99 | 0.98 | 0.98 | 0.98 | 0.98 |
| SL | Neuro | 8227 | 131 | Sensitivity | 0.94 | 0.77 | 0.38 | 0.12 | 0.07 | 0.04 | +0 | +0 | +0 |
| SL | Neuro | 8227 | 131 | Specificity | 0.29 | 0.63 | 0.91 | 0.97 | 0.99 | 0.99 | 1 | 1 | 1 |
| SL | Neuro | 8227 | 131 | Alert rate | 0.71 | 0.38 | 0.1 | 0.03 | 0.01 | 0.01 | +0 | +0 | +0 |
| SL | Neuro | 8227 | 131 | PPV | 0.02 | 0.03 | 0.06 | 0.07 | 0.1 | 0.1 | NaN | NaN | NaN |
| SL | Neuro | 8227 | 131 | NPV | 1 | 0.99 | 0.99 | 0.99 | 0.99 | 0.98 | 0.98 | 0.98 | 0.98 |
| XGB-ALL | Neuro | 8227 | 131 | Sensitivity | 0.92 | 0.7 | 0.38 | 0.12 | 0.09 | 0.07 | +0 | +0 | +0 |
| XGB-ALL | Neuro | 8227 | 131 | Specificity | 0.38 | 0.68 | 0.91 | 0.97 | 0.99 | 0.99 | 1 | 1 | 1 |
| XGB-ALL | Neuro | 8227 | 131 | Alert rate | 0.63 | 0.33 | 0.1 | 0.03 | 0.01 | 0.01 | +0 | +0 | +0 |
| XGB-ALL | Neuro | 8227 | 131 | PPV | 0.02 | 0.03 | 0.06 | 0.05 | 0.1 | 0.12 | NaN | NaN | NaN |
| XGB-ALL | Neuro | 8227 | 131 | NPV | 1 | 0.99 | 0.99 | 0.99 | 0.99 | 0.99 | 0.98 | 0.98 | 0.98 |
| XGB-LIM | Neuro | 8227 | 131 | Sensitivity | 0.93 | 0.72 | 0.38 | 0.26 | 0.15 | 0.08 | +0 | +0 | +0 |
| XGB-LIM | Neuro | 8227 | 131 | Specificity | 0.31 | 0.63 | 0.87 | 0.95 | 0.98 | 0.99 | 1 | 1 | 1 |
| XGB-LIM | Neuro | 8227 | 131 | Alert rate | 0.69 | 0.38 | 0.13 | 0.06 | 0.03 | 0.01 | +0 | +0 | +0 |
| XGB-LIM | Neuro | 8227 | 131 | PPV | 0.02 | 0.03 | 0.05 | 0.07 | 0.09 | 0.1 | NaN | NaN | NaN |
| XGB-LIM | Neuro | 8227 | 131 | NPV | 1 | 0.99 | 0.99 | 0.99 | 0.99 | 0.99 | 0.98 | 0.98 | 0.98 |
| CS-LIM | Internal Medicine | 6794 | 111 | Sensitivity | 1 | 0.99 | 0.7 | 0.38 | 0.32 | 0.27 | +0 | +0 | +0 |
| CS-LIM | Internal Medicine | 6794 | 111 | Specificity | 0.01 | 0.07 | 0.51 | 0.79 | 0.88 | 0.93 | 1 | 1 | 1 |
| CS-LIM | Internal Medicine | 6794 | 111 | Alert rate | 1 | 0.93 | 0.49 | 0.21 | 0.12 | 0.08 | +0 | +0 | +0 |
| CS-LIM | Internal Medicine | 6794 | 111 | PPV | 0.02 | 0.02 | 0.02 | 0.03 | 0.05 | 0.06 | +0 | NaN | NaN |
| CS-LIM | Internal Medicine | 6794 | 111 | NPV | 1 | 1 | 0.99 | 0.99 | 0.99 | 0.99 | 0.98 | 0.98 | 0.98 |
| RF-ALL | Internal Medicine | 6794 | 111 | Sensitivity | 1 | 1 | 0.73 | 0.42 | 0.25 | 0.13 | +0 | +0 | +0 |
| RF-ALL | Internal Medicine | 6794 | 111 | Specificity | 0.01 | 0.04 | 0.39 | 0.8 | 0.93 | 0.99 | 1 | 1 | 1 |
| RF-ALL | Internal Medicine | 6794 | 111 | Alert rate | 0.99 | 0.96 | 0.61 | 0.21 | 0.07 | 0.02 | +0 | +0 | +0 |
| RF-ALL | Internal Medicine | 6794 | 111 | PPV | 0.02 | 0.02 | 0.02 | 0.04 | 0.06 | 0.14 | NaN | NaN | NaN |
| RF-ALL | Internal Medicine | 6794 | 111 | NPV | 1 | 1 | 0.99 | 0.99 | 0.99 | 0.98 | 0.98 | 0.98 | 0.98 |
| RF-LIM | Internal Medicine | 6794 | 111 | Sensitivity | 1 | 0.99 | 0.65 | 0.43 | 0.21 | 0.03 | +0 | +0 | +0 |
| RF-LIM | Internal Medicine | 6794 | 111 | Specificity | 0.02 | 0.06 | 0.49 | 0.8 | 0.92 | 0.98 | 1 | 1 | 1 |
| RF-LIM | Internal Medicine | 6794 | 111 | Alert rate | 0.98 | 0.94 | 0.52 | 0.2 | 0.08 | 0.02 | +0 | +0 | +0 |
| RF-LIM | Internal Medicine | 6794 | 111 | PPV | 0.02 | 0.02 | 0.02 | 0.04 | 0.04 | 0.03 | NaN | NaN | NaN |
| RF-LIM | Internal Medicine | 6794 | 111 | NPV | 1 | 1 | 0.99 | 0.99 | 0.98 | 0.98 | 0.98 | 0.98 | 0.98 |
| SL | Internal Medicine | 6794 | 111 | Sensitivity | 1 | 0.96 | 0.64 | 0.48 | 0.36 | 0.27 | +0 | +0 | +0 |
| SL | Internal Medicine | 6794 | 111 | Specificity | 0.02 | 0.09 | 0.56 | 0.79 | 0.89 | 0.93 | 1 | 1 | 1 |
| SL | Internal Medicine | 6794 | 111 | Alert rate | 0.98 | 0.91 | 0.45 | 0.21 | 0.12 | 0.07 | +0 | +0 | +0 |
| SL | Internal Medicine | 6794 | 111 | PPV | 0.02 | 0.02 | 0.02 | 0.04 | 0.05 | 0.07 | +0 | NaN | NaN |
| SL | Internal Medicine | 6794 | 111 | NPV | 1 | 0.99 | 0.99 | 0.99 | 0.99 | 0.99 | 0.98 | 0.98 | 0.98 |
| XGB-ALL | Internal Medicine | 6794 | 111 | Sensitivity | 0.99 | 0.85 | 0.6 | 0.48 | 0.39 | 0.32 | 0.02 | +0 | +0 |
| XGB-ALL | Internal Medicine | 6794 | 111 | Specificity | 0.03 | 0.21 | 0.6 | 0.79 | 0.87 | 0.92 | 1 | 1 | 1 |
| XGB-ALL | Internal Medicine | 6794 | 111 | Alert rate | 0.97 | 0.79 | 0.41 | 0.22 | 0.13 | 0.09 | +0 | +0 | +0 |
| XGB-ALL | Internal Medicine | 6794 | 111 | PPV | 0.02 | 0.02 | 0.03 | 0.04 | 0.05 | 0.07 | 0.07 | +0 | NaN |
| XGB-ALL | Internal Medicine | 6794 | 111 | NPV | 1 | 0.99 | 0.99 | 0.99 | 0.99 | 0.99 | 0.98 | 0.98 | 0.98 |
| XGB-LIM | Internal Medicine | 6794 | 111 | Sensitivity | 1 | 0.87 | 0.6 | 0.41 | 0.25 | 0.2 | +0 | +0 | +0 |
| XGB-LIM | Internal Medicine | 6794 | 111 | Specificity | 0.03 | 0.2 | 0.64 | 0.81 | 0.9 | 0.94 | 1 | 1 | 1 |
| XGB-LIM | Internal Medicine | 6794 | 111 | Alert rate | 0.97 | 0.8 | 0.36 | 0.19 | 0.11 | 0.06 | +0 | +0 | +0 |
| XGB-LIM | Internal Medicine | 6794 | 111 | PPV | 0.02 | 0.02 | 0.03 | 0.04 | 0.04 | 0.06 | NaN | NaN | NaN |
| XGB-LIM | Internal Medicine | 6794 | 111 | NPV | 1 | 0.99 | 0.99 | 0.99 | 0.99 | 0.99 | 0.98 | 0.98 | 0.98 |
| CS-LIM | Pediatrics | 5530 | 31 | Sensitivity | 0.92 | 0.76 | 0.28 | 0.16 | 0.04 | 0.04 | +0 | +0 | +0 |
| CS-LIM | Pediatrics | 5530 | 31 | Specificity | 0.28 | 0.66 | 0.89 | 0.96 | 0.98 | 0.99 | 1 | 1 | 1 |
| CS-LIM | Pediatrics | 5530 | 31 | Alert rate | 0.72 | 0.34 | 0.11 | 0.05 | 0.02 | 0.01 | +0 | +0 | +0 |
| CS-LIM | Pediatrics | 5530 | 31 | PPV | 0.01 | 0.01 | 0.01 | 0.02 | 0.01 | 0.04 | NaN | NaN | NaN |
| CS-LIM | Pediatrics | 5530 | 31 | NPV | 1 | 1 | 1 | 1 | 1 | 1 | 1 | 1 | 1 |
| RF-ALL | Pediatrics | 5530 | 31 | Sensitivity | 0.96 | 0.88 | 0.68 | 0.16 | +0 | +0 | +0 | +0 | +0 |
| RF-ALL | Pediatrics | 5530 | 31 | Specificity | 0.15 | 0.47 | 0.81 | 0.94 | 1 | 1 | 1 | 1 | 1 |
| RF-ALL | Pediatrics | 5530 | 31 | Alert rate | 0.85 | 0.53 | 0.19 | 0.06 | +0 | +0 | +0 | +0 | +0 |
| RF-ALL | Pediatrics | 5530 | 31 | PPV | 0.01 | 0.01 | 0.02 | 0.01 | +0 | NaN | NaN | NaN | NaN |
| RF-ALL | Pediatrics | 5530 | 31 | NPV | 1 | 1 | 1 | 1 | 1 | 1 | 1 | 1 | 1 |
| RF-LIM | Pediatrics | 5530 | 31 | Sensitivity | 0.96 | 0.84 | 0.76 | 0.32 | 0.16 | 0.12 | +0 | +0 | +0 |
| RF-LIM | Pediatrics | 5530 | 31 | Specificity | 0.15 | 0.43 | 0.74 | 0.9 | 0.98 | 1 | 1 | 1 | 1 |
| RF-LIM | Pediatrics | 5530 | 31 | Alert rate | 0.85 | 0.58 | 0.26 | 0.1 | 0.02 | 0.01 | +0 | +0 | +0 |
| RF-LIM | Pediatrics | 5530 | 31 | PPV | 0.01 | 0.01 | 0.01 | 0.02 | 0.03 | 0.11 | NaN | NaN | NaN |
| RF-LIM | Pediatrics | 5530 | 31 | NPV | 1 | 1 | 1 | 1 | 1 | 1 | 1 | 1 | 1 |
| SL | Pediatrics | 5530 | 31 | Sensitivity | 0.92 | 0.84 | 0.6 | 0.12 | +0 | +0 | +0 | +0 | +0 |
| SL | Pediatrics | 5530 | 31 | Specificity | 0.25 | 0.59 | 0.86 | 0.95 | 0.98 | 0.99 | 1 | 1 | 1 |
| SL | Pediatrics | 5530 | 31 | Alert rate | 0.75 | 0.42 | 0.14 | 0.05 | 0.02 | 0.01 | +0 | +0 | +0 |
| SL | Pediatrics | 5530 | 31 | PPV | 0.01 | 0.01 | 0.02 | 0.01 | +0 | +0 | NaN | NaN | NaN |
| SL | Pediatrics | 5530 | 31 | NPV | 1 | 1 | 1 | 1 | 1 | 1 | 1 | 1 | 1 |
| XGB-ALL | Pediatrics | 5530 | 31 | Sensitivity | 0.88 | 0.84 | 0.44 | 0.12 | 0.04 | +0 | +0 | +0 | +0 |
| XGB-ALL | Pediatrics | 5530 | 31 | Specificity | 0.35 | 0.64 | 0.87 | 0.95 | 0.97 | 0.99 | 1 | 1 | 1 |
| XGB-ALL | Pediatrics | 5530 | 31 | Alert rate | 0.65 | 0.36 | 0.13 | 0.05 | 0.03 | 0.01 | +0 | +0 | +0 |
| XGB-ALL | Pediatrics | 5530 | 31 | PPV | 0.01 | 0.01 | 0.02 | 0.01 | 0.01 | +0 | +0 | NaN | NaN |
| XGB-ALL | Pediatrics | 5530 | 31 | NPV | 1 | 1 | 1 | 1 | 1 | 1 | 1 | 1 | 1 |
| XGB-LIM | Pediatrics | 5530 | 31 | Sensitivity | 0.88 | 0.84 | 0.44 | +0 | +0 | +0 | +0 | +0 | +0 |
| XGB-LIM | Pediatrics | 5530 | 31 | Specificity | 0.32 | 0.63 | 0.86 | 0.94 | 0.97 | 0.99 | 1 | 1 | 1 |
| XGB-LIM | Pediatrics | 5530 | 31 | Alert rate | 0.68 | 0.37 | 0.14 | 0.06 | 0.03 | 0.01 | +0 | +0 | +0 |
| XGB-LIM | Pediatrics | 5530 | 31 | PPV | 0.01 | 0.01 | 0.01 | +0 | +0 | +0 | +0 | NaN | NaN |
| XGB-LIM | Pediatrics | 5530 | 31 | NPV | 1 | 1 | 1 | 0.99 | 1 | 1 | 1 | 1 | 1 |
| CS-LIM | Nephrology | 5035 | 56 | Sensitivity | 1 | 0.92 | 0.43 | 0.27 | 0.18 | 0.1 | +0 | +0 | +0 |
| CS-LIM | Nephrology | 5035 | 56 | Specificity | 0.03 | 0.29 | 0.72 | 0.86 | 0.92 | 0.95 | 1 | 1 | 1 |
| CS-LIM | Nephrology | 5035 | 56 | Alert rate | 0.97 | 0.72 | 0.28 | 0.14 | 0.08 | 0.05 | +0 | +0 | +0 |
| CS-LIM | Nephrology | 5035 | 56 | PPV | 0.01 | 0.01 | 0.02 | 0.02 | 0.02 | 0.02 | NaN | NaN | NaN |
| CS-LIM | Nephrology | 5035 | 56 | NPV | 1 | 1 | 0.99 | 0.99 | 0.99 | 0.99 | 0.99 | 0.99 | 0.99 |
| RF-ALL | Nephrology | 5035 | 56 | Sensitivity | 0.98 | 0.84 | 0.33 | 0.16 | 0.02 | +0 | +0 | +0 | +0 |
| RF-ALL | Nephrology | 5035 | 56 | Specificity | 0.13 | 0.35 | 0.78 | 0.92 | 0.98 | 1 | 1 | 1 | 1 |
| RF-ALL | Nephrology | 5035 | 56 | Alert rate | 0.87 | 0.65 | 0.22 | 0.08 | 0.02 | +0 | +0 | +0 | +0 |
| RF-ALL | Nephrology | 5035 | 56 | PPV | 0.01 | 0.01 | 0.02 | 0.02 | 0.01 | +0 | NaN | NaN | NaN |
| RF-ALL | Nephrology | 5035 | 56 | NPV | 1 | 0.99 | 0.99 | 0.99 | 0.99 | 0.99 | 0.99 | 0.99 | 0.99 |
| RF-LIM | Nephrology | 5035 | 56 | Sensitivity | 0.98 | 0.92 | 0.37 | 0.24 | 0.08 | 0.02 | +0 | +0 | +0 |
| RF-LIM | Nephrology | 5035 | 56 | Specificity | 0.11 | 0.31 | 0.72 | 0.86 | 0.94 | 0.99 | 1 | 1 | 1 |
| RF-LIM | Nephrology | 5035 | 56 | Alert rate | 0.89 | 0.69 | 0.28 | 0.14 | 0.06 | 0.01 | +0 | +0 | +0 |
| RF-LIM | Nephrology | 5035 | 56 | PPV | 0.01 | 0.01 | 0.01 | 0.02 | 0.02 | 0.03 | NaN | NaN | NaN |
| RF-LIM | Nephrology | 5035 | 56 | NPV | 1 | 1 | 0.99 | 0.99 | 0.99 | 0.99 | 0.99 | 0.99 | 0.99 |
| SL | Nephrology | 5035 | 56 | Sensitivity | 0.98 | 0.65 | 0.31 | 0.2 | 0.06 | +0 | +0 | +0 | +0 |
| SL | Nephrology | 5035 | 56 | Specificity | 0.18 | 0.49 | 0.81 | 0.9 | 0.96 | 0.98 | 1 | 1 | 1 |
| SL | Nephrology | 5035 | 56 | Alert rate | 0.82 | 0.51 | 0.2 | 0.1 | 0.04 | 0.02 | +0 | +0 | +0 |
| SL | Nephrology | 5035 | 56 | PPV | 0.01 | 0.01 | 0.02 | 0.02 | 0.02 | +0 | +0 | NaN | NaN |
| SL | Nephrology | 5035 | 56 | NPV | 1 | 0.99 | 0.99 | 0.99 | 0.99 | 0.99 | 0.99 | 0.99 | 0.99 |
| XGB-ALL | Nephrology | 5035 | 56 | Sensitivity | 0.82 | 0.59 | 0.31 | 0.18 | 0.08 | +0 | +0 | +0 | +0 |
| XGB-ALL | Nephrology | 5035 | 56 | Specificity | 0.27 | 0.57 | 0.82 | 0.9 | 0.95 | 0.97 | 1 | 1 | 1 |
| XGB-ALL | Nephrology | 5035 | 56 | Alert rate | 0.73 | 0.43 | 0.18 | 0.1 | 0.05 | 0.03 | +0 | +0 | +0 |
| XGB-ALL | Nephrology | 5035 | 56 | PPV | 0.01 | 0.02 | 0.02 | 0.02 | 0.02 | +0 | +0 | +0 | NaN |
| XGB-ALL | Nephrology | 5035 | 56 | NPV | 0.99 | 0.99 | 0.99 | 0.99 | 0.99 | 0.99 | 0.99 | 0.99 | 0.99 |
| XGB-LIM | Nephrology | 5035 | 56 | Sensitivity | 1 | 0.63 | 0.35 | 0.27 | 0.18 | 0.16 | +0 | +0 | +0 |
| XGB-LIM | Nephrology | 5035 | 56 | Specificity | 0.19 | 0.47 | 0.75 | 0.85 | 0.92 | 0.95 | 1 | 1 | 1 |
| XGB-LIM | Nephrology | 5035 | 56 | Alert rate | 0.81 | 0.53 | 0.25 | 0.15 | 0.09 | 0.05 | +0 | +0 | +0 |
| XGB-LIM | Nephrology | 5035 | 56 | PPV | 0.01 | 0.01 | 0.02 | 0.02 | 0.02 | 0.04 | NaN | NaN | NaN |
| XGB-LIM | Nephrology | 5035 | 56 | NPV | 1 | 0.99 | 0.99 | 0.99 | 0.99 | 0.99 | 0.99 | 0.99 | 0.99 |
| CS-LIM | Urology | 4854 | 86 | Sensitivity | 1 | 0.84 | 0.59 | 0.42 | 0.16 | 0.02 | +0 | +0 | +0 |
| CS-LIM | Urology | 4854 | 86 | Specificity | 0.07 | 0.39 | 0.84 | 0.92 | 0.96 | 0.99 | 1 | 1 | 1 |
| CS-LIM | Urology | 4854 | 86 | Alert rate | 0.93 | 0.61 | 0.17 | 0.09 | 0.04 | 0.02 | +0 | +0 | +0 |
| CS-LIM | Urology | 4854 | 86 | PPV | 0.02 | 0.02 | 0.06 | 0.09 | 0.07 | 0.03 | NaN | NaN | NaN |
| CS-LIM | Urology | 4854 | 86 | NPV | 1 | 0.99 | 0.99 | 0.99 | 0.98 | 0.98 | 0.98 | 0.98 | 0.98 |
| RF-ALL | Urology | 4854 | 86 | Sensitivity | 1 | 0.81 | 0.6 | 0.36 | 0.17 | 0.01 | +0 | +0 | +0 |
| RF-ALL | Urology | 4854 | 86 | Specificity | 0.09 | 0.37 | 0.86 | 0.93 | 0.98 | 1 | 1 | 1 | 1 |
| RF-ALL | Urology | 4854 | 86 | Alert rate | 0.91 | 0.63 | 0.15 | 0.08 | 0.02 | +0 | +0 | +0 | +0 |
| RF-ALL | Urology | 4854 | 86 | PPV | 0.02 | 0.02 | 0.07 | 0.08 | 0.12 | 0.07 | NaN | NaN | NaN |
| RF-ALL | Urology | 4854 | 86 | NPV | 1 | 0.99 | 0.99 | 0.99 | 0.98 | 0.98 | 0.98 | 0.98 | 0.98 |
| RF-LIM | Urology | 4854 | 86 | Sensitivity | 0.99 | 0.88 | 0.6 | 0.51 | 0.22 | 0.07 | +0 | +0 | +0 |
| RF-LIM | Urology | 4854 | 86 | Specificity | 0.08 | 0.33 | 0.82 | 0.9 | 0.96 | 0.99 | 1 | 1 | 1 |
| RF-LIM | Urology | 4854 | 86 | Alert rate | 0.92 | 0.67 | 0.19 | 0.11 | 0.04 | 0.01 | +0 | +0 | +0 |
| RF-LIM | Urology | 4854 | 86 | PPV | 0.02 | 0.02 | 0.06 | 0.08 | 0.09 | 0.13 | NaN | NaN | NaN |
| RF-LIM | Urology | 4854 | 86 | NPV | 1 | 0.99 | 0.99 | 0.99 | 0.99 | 0.98 | 0.98 | 0.98 | 0.98 |
| SL | Urology | 4854 | 86 | Sensitivity | 1 | 0.86 | 0.57 | 0.43 | 0.27 | 0.13 | +0 | +0 | +0 |
| SL | Urology | 4854 | 86 | Specificity | 0.1 | 0.37 | 0.82 | 0.92 | 0.96 | 0.98 | 1 | 1 | 1 |
| SL | Urology | 4854 | 86 | Alert rate | 0.9 | 0.63 | 0.19 | 0.09 | 0.05 | 0.02 | +0 | +0 | +0 |
| SL | Urology | 4854 | 86 | PPV | 0.02 | 0.02 | 0.05 | 0.09 | 0.1 | 0.09 | NaN | NaN | NaN |
| SL | Urology | 4854 | 86 | NPV | 1 | 0.99 | 0.99 | 0.99 | 0.99 | 0.98 | 0.98 | 0.98 | 0.98 |
| XGB-ALL | Urology | 4854 | 86 | Sensitivity | 0.99 | 0.85 | 0.59 | 0.38 | 0.29 | 0.16 | +0 | +0 | +0 |
| XGB-ALL | Urology | 4854 | 86 | Specificity | 0.14 | 0.4 | 0.79 | 0.91 | 0.95 | 0.97 | 1 | 1 | 1 |
| XGB-ALL | Urology | 4854 | 86 | Alert rate | 0.86 | 0.6 | 0.22 | 0.1 | 0.06 | 0.03 | +0 | +0 | +0 |
| XGB-ALL | Urology | 4854 | 86 | PPV | 0.02 | 0.03 | 0.05 | 0.07 | 0.09 | 0.09 | +0 | NaN | NaN |
| XGB-ALL | Urology | 4854 | 86 | NPV | 1 | 0.99 | 0.99 | 0.99 | 0.99 | 0.98 | 0.98 | 0.98 | 0.98 |
| XGB-LIM | Urology | 4854 | 86 | Sensitivity | 0.99 | 0.84 | 0.65 | 0.45 | 0.3 | 0.22 | +0 | +0 | +0 |
| XGB-LIM | Urology | 4854 | 86 | Specificity | 0.12 | 0.43 | 0.81 | 0.9 | 0.94 | 0.97 | 1 | 1 | 1 |
| XGB-LIM | Urology | 4854 | 86 | Alert rate | 0.88 | 0.58 | 0.2 | 0.1 | 0.06 | 0.04 | +0 | +0 | +0 |
| XGB-LIM | Urology | 4854 | 86 | PPV | 0.02 | 0.03 | 0.06 | 0.08 | 0.09 | 0.11 | NaN | NaN | NaN |
| XGB-LIM | Urology | 4854 | 86 | NPV | 1 | 0.99 | 0.99 | 0.99 | 0.99 | 0.99 | 0.98 | 0.98 | 0.98 |
| CS-LIM | Endocrinology | 4015 | 67 | Sensitivity | 1 | 0.91 | 0.65 | 0.39 | 0.17 | 0.02 | +0 | +0 | +0 |
| CS-LIM | Endocrinology | 4015 | 67 | Specificity | 0.03 | 0.2 | 0.67 | 0.82 | 0.93 | 0.98 | 1 | 1 | 1 |
| CS-LIM | Endocrinology | 4015 | 67 | Alert rate | 0.97 | 0.8 | 0.33 | 0.18 | 0.07 | 0.02 | +0 | +0 | +0 |
| CS-LIM | Endocrinology | 4015 | 67 | PPV | 0.01 | 0.02 | 0.03 | 0.03 | 0.03 | 0.01 | NaN | NaN | NaN |
| CS-LIM | Endocrinology | 4015 | 67 | NPV | 1 | 0.99 | 0.99 | 0.99 | 0.99 | 0.99 | 0.99 | 0.99 | 0.99 |
| RF-ALL | Endocrinology | 4015 | 67 | Sensitivity | 1 | 1 | 0.67 | 0.24 | 0.02 | +0 | +0 | +0 | +0 |
| RF-ALL | Endocrinology | 4015 | 67 | Specificity | 0.03 | 0.12 | 0.66 | 0.88 | 0.97 | 1 | 1 | 1 | 1 |
| RF-ALL | Endocrinology | 4015 | 67 | Alert rate | 0.97 | 0.88 | 0.35 | 0.12 | 0.03 | +0 | +0 | +0 | +0 |
| RF-ALL | Endocrinology | 4015 | 67 | PPV | 0.01 | 0.02 | 0.03 | 0.03 | 0.01 | +0 | NaN | NaN | NaN |
| RF-ALL | Endocrinology | 4015 | 67 | NPV | 1 | 1 | 0.99 | 0.99 | 0.99 | 0.99 | 0.99 | 0.99 | 0.99 |
| RF-LIM | Endocrinology | 4015 | 67 | Sensitivity | 1 | 0.98 | 0.72 | 0.41 | 0.11 | +0 | +0 | +0 | +0 |
| RF-LIM | Endocrinology | 4015 | 67 | Specificity | 0.04 | 0.19 | 0.65 | 0.85 | 0.97 | 1 | 1 | 1 | 1 |
| RF-LIM | Endocrinology | 4015 | 67 | Alert rate | 0.96 | 0.81 | 0.36 | 0.15 | 0.03 | +0 | +0 | +0 | +0 |
| RF-LIM | Endocrinology | 4015 | 67 | PPV | 0.01 | 0.02 | 0.03 | 0.04 | 0.05 | +0 | NaN | NaN | NaN |
| RF-LIM | Endocrinology | 4015 | 67 | NPV | 1 | 1 | 0.99 | 0.99 | 0.99 | 0.99 | 0.99 | 0.99 | 0.99 |
| SL | Endocrinology | 4015 | 67 | Sensitivity | 1 | 0.91 | 0.67 | 0.3 | 0.07 | 0.02 | +0 | +0 | +0 |
| SL | Endocrinology | 4015 | 67 | Specificity | 0.05 | 0.42 | 0.73 | 0.88 | 0.95 | 0.98 | 1 | 1 | 1 |
| SL | Endocrinology | 4015 | 67 | Alert rate | 0.95 | 0.59 | 0.27 | 0.13 | 0.05 | 0.02 | +0 | +0 | +0 |
| SL | Endocrinology | 4015 | 67 | PPV | 0.01 | 0.02 | 0.03 | 0.03 | 0.02 | 0.01 | NaN | NaN | NaN |
| SL | Endocrinology | 4015 | 67 | NPV | 1 | 1 | 0.99 | 0.99 | 0.99 | 0.99 | 0.99 | 0.99 | 0.99 |
| XGB-ALL | Endocrinology | 4015 | 67 | Sensitivity | 0.98 | 0.8 | 0.67 | 0.35 | 0.09 | 0.04 | +0 | +0 | +0 |
| XGB-ALL | Endocrinology | 4015 | 67 | Specificity | 0.22 | 0.54 | 0.76 | 0.88 | 0.94 | 0.96 | 1 | 1 | 1 |
| XGB-ALL | Endocrinology | 4015 | 67 | Alert rate | 0.78 | 0.47 | 0.24 | 0.13 | 0.06 | 0.04 | +0 | +0 | +0 |
| XGB-ALL | Endocrinology | 4015 | 67 | PPV | 0.02 | 0.02 | 0.04 | 0.04 | 0.02 | 0.02 | +0 | NaN | NaN |
| XGB-ALL | Endocrinology | 4015 | 67 | NPV | 1 | 1 | 0.99 | 0.99 | 0.99 | 0.99 | 0.99 | 0.99 | 0.99 |
| XGB-LIM | Endocrinology | 4015 | 67 | Sensitivity | 0.96 | 0.87 | 0.67 | 0.39 | 0.24 | 0.11 | +0 | +0 | +0 |
| XGB-LIM | Endocrinology | 4015 | 67 | Specificity | 0.15 | 0.46 | 0.76 | 0.87 | 0.94 | 0.97 | 1 | 1 | 1 |
| XGB-LIM | Endocrinology | 4015 | 67 | Alert rate | 0.86 | 0.54 | 0.25 | 0.13 | 0.06 | 0.03 | +0 | +0 | +0 |
| XGB-LIM | Endocrinology | 4015 | 67 | PPV | 0.02 | 0.02 | 0.04 | 0.04 | 0.05 | 0.04 | NaN | NaN | NaN |
| XGB-LIM | Endocrinology | 4015 | 67 | NPV | 1 | 1 | 0.99 | 0.99 | 0.99 | 0.99 | 0.99 | 0.99 | 0.99 |
| CS-LIM | Emergency | 3730 | 39 | Sensitivity | 1 | 0.59 | 0.18 | 0.1 | 0.08 | 0.05 | +0 | +0 | +0 |
| CS-LIM | Emergency | 3730 | 39 | Specificity | 0.04 | 0.43 | 0.83 | 0.94 | 0.97 | 0.99 | 1 | 1 | 1 |
| CS-LIM | Emergency | 3730 | 39 | Alert rate | 0.96 | 0.57 | 0.17 | 0.06 | 0.03 | 0.01 | +0 | +0 | +0 |
| CS-LIM | Emergency | 3730 | 39 | PPV | 0.01 | 0.01 | 0.01 | 0.02 | 0.03 | 0.05 | NaN | NaN | NaN |
| CS-LIM | Emergency | 3730 | 39 | NPV | 1 | 0.99 | 0.99 | 0.99 | 0.99 | 0.99 | 0.99 | 0.99 | 0.99 |
| RF-ALL | Emergency | 3730 | 39 | Sensitivity | 1 | 0.95 | 0.33 | 0.05 | 0.03 | +0 | +0 | +0 | +0 |
| RF-ALL | Emergency | 3730 | 39 | Specificity | +0 | 0.11 | 0.69 | 0.93 | 0.99 | 1 | 1 | 1 | 1 |
| RF-ALL | Emergency | 3730 | 39 | Alert rate | 1 | 0.89 | 0.31 | 0.07 | 0.01 | +0 | +0 | +0 | +0 |
| RF-ALL | Emergency | 3730 | 39 | PPV | 0.01 | 0.01 | 0.01 | 0.01 | 0.02 | +0 | NaN | NaN | NaN |
| RF-ALL | Emergency | 3730 | 39 | NPV | 1 | 0.99 | 0.99 | 0.99 | 0.99 | 0.99 | 0.99 | 0.99 | 0.99 |
| RF-LIM | Emergency | 3730 | 39 | Sensitivity | 1 | 0.9 | 0.36 | 0.13 | 0.05 | +0 | +0 | +0 | +0 |
| RF-LIM | Emergency | 3730 | 39 | Specificity | +0 | 0.15 | 0.69 | 0.89 | 0.98 | 1 | 1 | 1 | 1 |
| RF-LIM | Emergency | 3730 | 39 | Alert rate | 1 | 0.85 | 0.31 | 0.11 | 0.02 | +0 | +0 | +0 | +0 |
| RF-LIM | Emergency | 3730 | 39 | PPV | 0.01 | 0.01 | 0.01 | 0.01 | 0.03 | +0 | NaN | NaN | NaN |
| RF-LIM | Emergency | 3730 | 39 | NPV | 1 | 0.99 | 0.99 | 0.99 | 0.99 | 0.99 | 0.99 | 0.99 | 0.99 |
| SL | Emergency | 3730 | 39 | Sensitivity | 0.97 | 0.87 | 0.38 | 0.18 | 0.1 | +0 | +0 | +0 | +0 |
| SL | Emergency | 3730 | 39 | Specificity | 0.01 | 0.23 | 0.75 | 0.94 | 0.98 | 0.99 | 1 | 1 | 1 |
| SL | Emergency | 3730 | 39 | Alert rate | 0.99 | 0.77 | 0.25 | 0.07 | 0.03 | 0.01 | +0 | +0 | +0 |
| SL | Emergency | 3730 | 39 | PPV | 0.01 | 0.01 | 0.02 | 0.03 | 0.04 | +0 | NaN | NaN | NaN |
| SL | Emergency | 3730 | 39 | NPV | 0.97 | 0.99 | 0.99 | 0.99 | 0.99 | 0.99 | 0.99 | 0.99 | 0.99 |
| XGB-ALL | Emergency | 3730 | 39 | Sensitivity | 0.97 | 0.77 | 0.33 | 0.21 | 0.15 | +0 | +0 | +0 | +0 |
| XGB-ALL | Emergency | 3730 | 39 | Specificity | 0.05 | 0.32 | 0.77 | 0.92 | 0.97 | 0.98 | 1 | 1 | 1 |
| XGB-ALL | Emergency | 3730 | 39 | Alert rate | 0.95 | 0.68 | 0.23 | 0.08 | 0.03 | 0.02 | +0 | +0 | +0 |
| XGB-ALL | Emergency | 3730 | 39 | PPV | 0.01 | 0.01 | 0.01 | 0.03 | 0.05 | +0 | +0 | NaN | NaN |
| XGB-ALL | Emergency | 3730 | 39 | NPV | 0.99 | 0.99 | 0.99 | 0.99 | 0.99 | 0.99 | 0.99 | 0.99 | 0.99 |
| XGB-LIM | Emergency | 3730 | 39 | Sensitivity | 0.97 | 0.87 | 0.38 | 0.23 | 0.08 | 0.03 | +0 | +0 | +0 |
| XGB-LIM | Emergency | 3730 | 39 | Specificity | 0.04 | 0.29 | 0.72 | 0.9 | 0.96 | 0.99 | 1 | 1 | 1 |
| XGB-LIM | Emergency | 3730 | 39 | Alert rate | 0.96 | 0.72 | 0.28 | 0.1 | 0.04 | 0.01 | +0 | +0 | +0 |
| XGB-LIM | Emergency | 3730 | 39 | PPV | 0.01 | 0.01 | 0.01 | 0.02 | 0.02 | 0.02 | NaN | NaN | NaN |
| XGB-LIM | Emergency | 3730 | 39 | NPV | 0.99 | 1 | 0.99 | 0.99 | 0.99 | 0.99 | 0.99 | 0.99 | 0.99 |
| CS-LIM | ORL | 3569 | 48 | Sensitivity | 0.98 | 0.85 | 0.63 | 0.44 | 0.07 | 0.05 | +0 | +0 | +0 |
| CS-LIM | ORL | 3569 | 48 | Specificity | 0.12 | 0.47 | 0.81 | 0.92 | 0.98 | 0.99 | 1 | 1 | 1 |
| CS-LIM | ORL | 3569 | 48 | Alert rate | 0.88 | 0.54 | 0.2 | 0.08 | 0.02 | 0.01 | +0 | +0 | +0 |
| CS-LIM | ORL | 3569 | 48 | PPV | 0.01 | 0.02 | 0.04 | 0.06 | 0.04 | 0.08 | NaN | NaN | NaN |
| CS-LIM | ORL | 3569 | 48 | NPV | 1 | 1 | 0.99 | 0.99 | 0.99 | 0.99 | 0.99 | 0.99 | 0.99 |
| RF-ALL | ORL | 3569 | 48 | Sensitivity | 1 | 0.85 | 0.66 | 0.27 | 0.15 | +0 | +0 | +0 | +0 |
| RF-ALL | ORL | 3569 | 48 | Specificity | 0.14 | 0.46 | 0.79 | 0.92 | 0.98 | 1 | 1 | 1 | 1 |
| RF-ALL | ORL | 3569 | 48 | Alert rate | 0.86 | 0.55 | 0.22 | 0.08 | 0.02 | +0 | +0 | +0 | +0 |
| RF-ALL | ORL | 3569 | 48 | PPV | 0.01 | 0.02 | 0.04 | 0.04 | 0.08 | +0 | NaN | NaN | NaN |
| RF-ALL | ORL | 3569 | 48 | NPV | 1 | 1 | 0.99 | 0.99 | 0.99 | 0.99 | 0.99 | 0.99 | 0.99 |
| RF-LIM | ORL | 3569 | 48 | Sensitivity | 1 | 0.93 | 0.73 | 0.37 | 0.12 | +0 | +0 | +0 | +0 |
| RF-LIM | ORL | 3569 | 48 | Specificity | 0.14 | 0.38 | 0.73 | 0.86 | 0.96 | 1 | 1 | 1 | 1 |
| RF-LIM | ORL | 3569 | 48 | Alert rate | 0.86 | 0.62 | 0.27 | 0.14 | 0.04 | +0 | +0 | +0 | +0 |
| RF-LIM | ORL | 3569 | 48 | PPV | 0.01 | 0.02 | 0.03 | 0.03 | 0.04 | +0 | NaN | NaN | NaN |
| RF-LIM | ORL | 3569 | 48 | NPV | 1 | 1 | 1 | 0.99 | 0.99 | 0.99 | 0.99 | 0.99 | 0.99 |
| SL | ORL | 3569 | 48 | Sensitivity | 0.98 | 0.85 | 0.61 | 0.46 | 0.15 | +0 | +0 | +0 | +0 |
| SL | ORL | 3569 | 48 | Specificity | 0.2 | 0.63 | 0.83 | 0.91 | 0.97 | 0.99 | 1 | 1 | 1 |
| SL | ORL | 3569 | 48 | Alert rate | 0.8 | 0.37 | 0.18 | 0.09 | 0.03 | 0.01 | +0 | +0 | +0 |
| SL | ORL | 3569 | 48 | PPV | 0.01 | 0.03 | 0.04 | 0.06 | 0.06 | +0 | NaN | NaN | NaN |
| SL | ORL | 3569 | 48 | NPV | 1 | 1 | 0.99 | 0.99 | 0.99 | 0.99 | 0.99 | 0.99 | 0.99 |
| XGB-ALL | ORL | 3569 | 48 | Sensitivity | 0.93 | 0.78 | 0.61 | 0.44 | 0.24 | 0.05 | +0 | +0 | +0 |
| XGB-ALL | ORL | 3569 | 48 | Specificity | 0.37 | 0.68 | 0.85 | 0.92 | 0.96 | 0.98 | 1 | 1 | 1 |
| XGB-ALL | ORL | 3569 | 48 | Alert rate | 0.64 | 0.32 | 0.16 | 0.09 | 0.04 | 0.02 | +0 | +0 | +0 |
| XGB-ALL | ORL | 3569 | 48 | PPV | 0.02 | 0.03 | 0.05 | 0.06 | 0.07 | 0.03 | NaN | NaN | NaN |
| XGB-ALL | ORL | 3569 | 48 | NPV | 1 | 1 | 0.99 | 0.99 | 0.99 | 0.99 | 0.99 | 0.99 | 0.99 |
| XGB-LIM | ORL | 3569 | 48 | Sensitivity | 1 | 0.83 | 0.63 | 0.39 | 0.27 | 0.17 | +0 | +0 | +0 |
| XGB-LIM | ORL | 3569 | 48 | Specificity | 0.22 | 0.59 | 0.8 | 0.88 | 0.94 | 0.97 | 1 | 1 | 1 |
| XGB-LIM | ORL | 3569 | 48 | Alert rate | 0.78 | 0.42 | 0.2 | 0.12 | 0.07 | 0.03 | +0 | +0 | +0 |
| XGB-LIM | ORL | 3569 | 48 | PPV | 0.02 | 0.02 | 0.04 | 0.04 | 0.05 | 0.06 | NaN | NaN | NaN |
| XGB-LIM | ORL | 3569 | 48 | NPV | 1 | 1 | 0.99 | 0.99 | 0.99 | 0.99 | 0.99 | 0.99 | 0.99 |
| CS-LIM | Other | 1451 | 8 | Sensitivity | 1 | 0.88 | 0.12 | +0 | +0 | +0 | +0 | +0 | +0 |
| CS-LIM | Other | 1451 | 8 | Specificity | 0.03 | 0.3 | 0.76 | 0.94 | 0.99 | 1 | 1 | 1 | 1 |
| CS-LIM | Other | 1451 | 8 | Alert rate | 0.97 | 0.7 | 0.24 | 0.06 | 0.01 | +0 | +0 | +0 | +0 |
| CS-LIM | Other | 1451 | 8 | PPV | 0.01 | 0.01 | +0 | +0 | +0 | +0 | NaN | NaN | NaN |
| CS-LIM | Other | 1451 | 8 | NPV | 1 | 1 | 0.99 | 0.99 | 0.99 | 0.99 | 0.99 | 0.99 | 0.99 |
| RF-ALL | Other | 1451 | 8 | Sensitivity | 1 | 1 | 0.38 | 0.12 | +0 | +0 | +0 | +0 | +0 |
| RF-ALL | Other | 1451 | 8 | Specificity | 0.04 | 0.2 | 0.75 | 0.93 | 1 | 1 | 1 | 1 | 1 |
| RF-ALL | Other | 1451 | 8 | Alert rate | 0.96 | 0.8 | 0.25 | 0.07 | +0 | +0 | +0 | +0 | +0 |
| RF-ALL | Other | 1451 | 8 | PPV | 0.01 | 0.01 | 0.01 | 0.01 | +0 | NaN | NaN | NaN | NaN |
| RF-ALL | Other | 1451 | 8 | NPV | 1 | 1 | 0.99 | 0.99 | 0.99 | 0.99 | 0.99 | 0.99 | 0.99 |
| RF-LIM | Other | 1451 | 8 | Sensitivity | 1 | 1 | 0.38 | 0.12 | +0 | +0 | +0 | +0 | +0 |
| RF-LIM | Other | 1451 | 8 | Specificity | 0.06 | 0.25 | 0.78 | 0.92 | 0.98 | 1 | 1 | 1 | 1 |
| RF-LIM | Other | 1451 | 8 | Alert rate | 0.94 | 0.76 | 0.22 | 0.08 | 0.02 | +0 | +0 | +0 | +0 |
| RF-LIM | Other | 1451 | 8 | PPV | 0.01 | 0.01 | 0.01 | 0.01 | +0 | NaN | NaN | NaN | NaN |
| RF-LIM | Other | 1451 | 8 | NPV | 1 | 1 | 0.99 | 0.99 | 0.99 | 0.99 | 0.99 | 0.99 | 0.99 |
| SL | Other | 1451 | 8 | Sensitivity | 1 | 0.88 | 0.12 | +0 | +0 | +0 | +0 | +0 | +0 |
| SL | Other | 1451 | 8 | Specificity | 0.1 | 0.44 | 0.81 | 0.95 | 0.99 | 1 | 1 | 1 | 1 |
| SL | Other | 1451 | 8 | Alert rate | 0.9 | 0.56 | 0.19 | 0.05 | 0.01 | +0 | +0 | +0 | +0 |
| SL | Other | 1451 | 8 | PPV | 0.01 | 0.01 | +0 | +0 | +0 | +0 | NaN | NaN | NaN |
| SL | Other | 1451 | 8 | NPV | 1 | 1 | 0.99 | 0.99 | 0.99 | 0.99 | 0.99 | 0.99 | 0.99 |
| XGB-ALL | Other | 1451 | 8 | Sensitivity | 0.88 | 0.12 | +0 | +0 | +0 | +0 | +0 | +0 | +0 |
| XGB-ALL | Other | 1451 | 8 | Specificity | 0.23 | 0.55 | 0.81 | 0.95 | 0.98 | 0.99 | 1 | 1 | 1 |
| XGB-ALL | Other | 1451 | 8 | Alert rate | 0.77 | 0.45 | 0.19 | 0.05 | 0.02 | 0.01 | +0 | +0 | +0 |
| XGB-ALL | Other | 1451 | 8 | PPV | 0.01 | +0 | +0 | +0 | +0 | +0 | NaN | NaN | NaN |
| XGB-ALL | Other | 1451 | 8 | NPV | 1 | 0.99 | 0.99 | 0.99 | 0.99 | 0.99 | 0.99 | 0.99 | 0.99 |
| XGB-LIM | Other | 1451 | 8 | Sensitivity | 1 | 0.88 | +0 | +0 | +0 | +0 | +0 | +0 | +0 |
| XGB-LIM | Other | 1451 | 8 | Specificity | 0.16 | 0.48 | 0.78 | 0.88 | 0.93 | 0.97 | 1 | 1 | 1 |
| XGB-LIM | Other | 1451 | 8 | Alert rate | 0.84 | 0.52 | 0.22 | 0.12 | 0.07 | 0.03 | +0 | +0 | +0 |
| XGB-LIM | Other | 1451 | 8 | PPV | 0.01 | 0.01 | +0 | +0 | +0 | +0 | NaN | NaN | NaN |
| XGB-LIM | Other | 1451 | 8 | NPV | 1 | 1 | 0.99 | 0.99 | 0.99 | 0.99 | 0.99 | 0.99 | 0.99 |

Table 12: Threshold dependent metrics at different thresholds for each ward. PPV = Positive Predictive Value; NPV = Negative Predictive Value. UNKNOWN_VALUE = The paediatric ward has changed name over time and the mapping done in training data did not cover the values in the test data.

### Supplementary material 8 - Model investigation

#### Notable shifts of percentage of missingness of variables per year

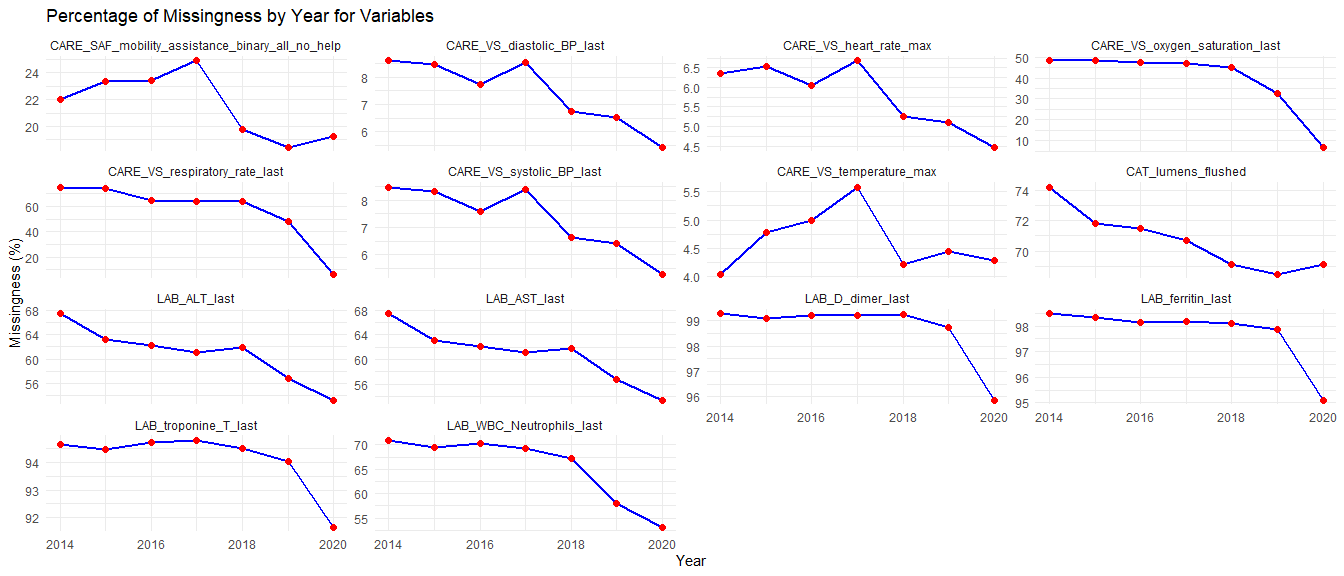

Figure 22: Notable shifts of percentage of missingness of variables per year

#### Notable shifts of percentage of sparseness of variables per year

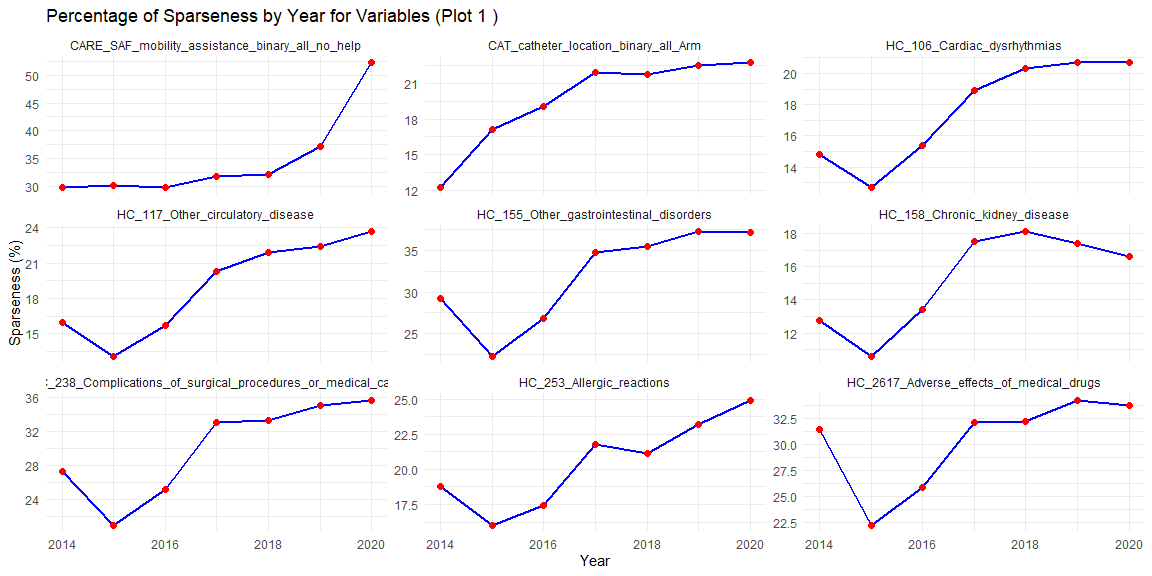

Figure 23: Notable shifts of percentage of sparseness of variables per year

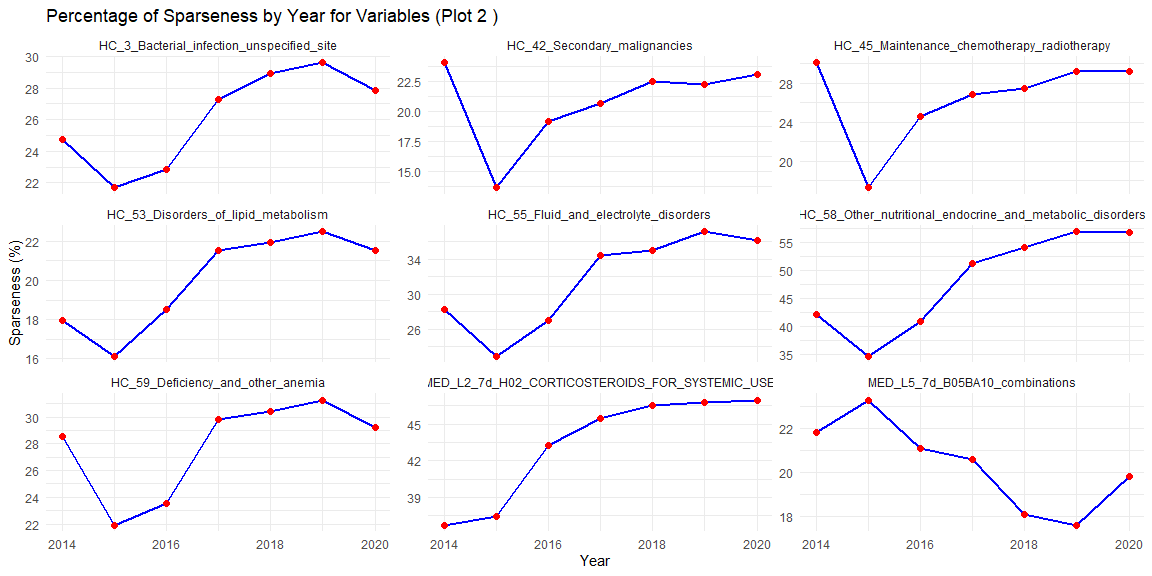

Figure 24: Notable shifts of percentage of sparseness of variables per year

#### Density distribution of log(D-dimer) before imputation

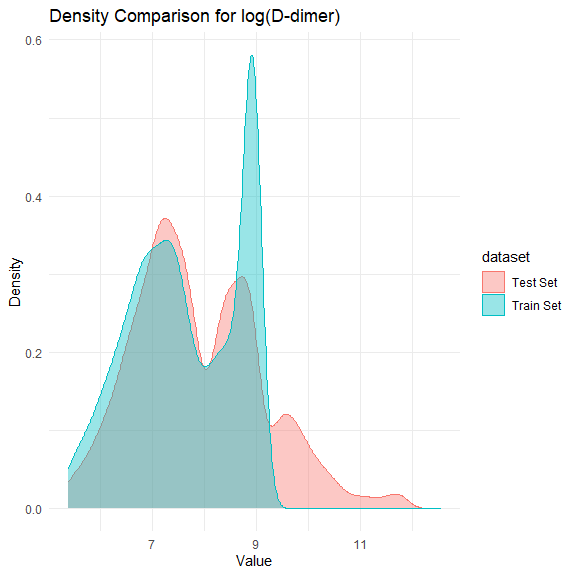

Figure 25: Density distribution of log(D-dimer) before imputation

#### Calibration curves of landmark cause-specific supermodels with and without D-dimer

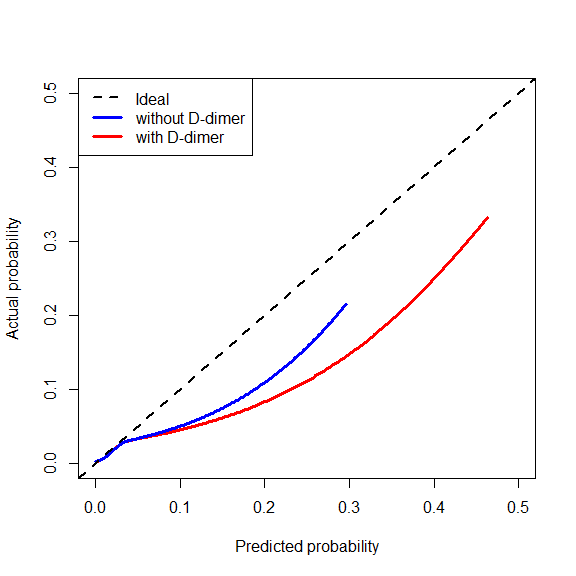

Figure 26: Calibration curves using restricted cubic splines for landmark cause-specific supermodel on temporal evaluation using all landmarks. The dashed grey line represents the identity function (perfect calibration).

#### Percentage of CLABSI per medical specialty from 2014 to 2020

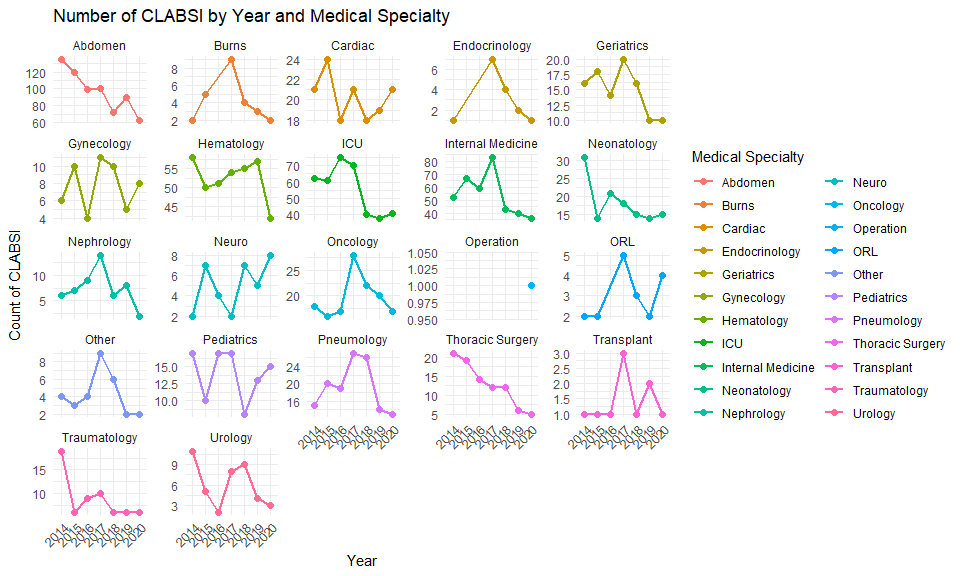

Figure 27: The Number and percentage of CLABSI per medical specialty from 2014 to 2020. The number is calculated as the count of catheter episodes which eventually experienced CLABSI at specific medical specialty at the corresponding year. The percentage is calculated using the number of catheter episodes which eventually experienced CLABSI at specific medical specialty divided by the total number of catheter episodes at the corresponding year.

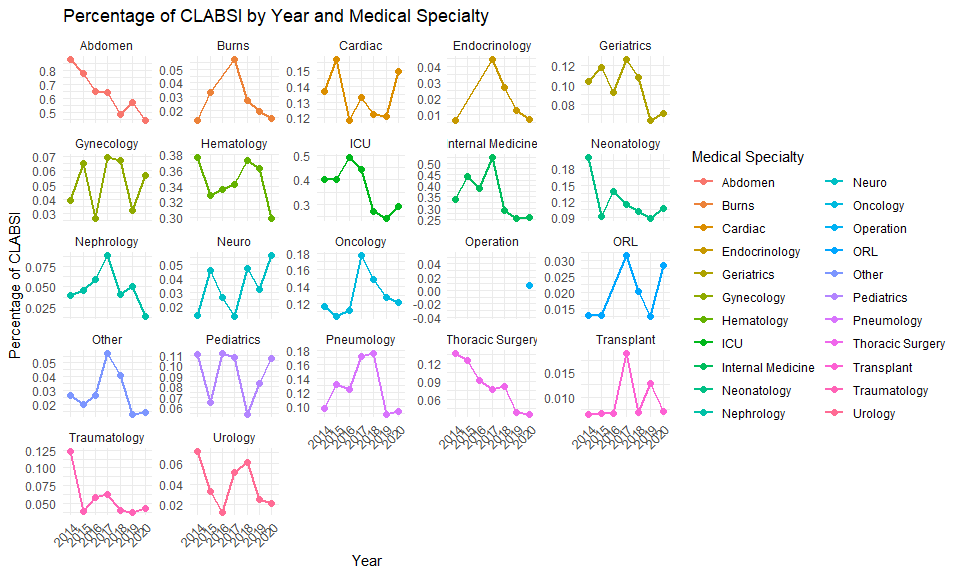

Figure 28: The Number and percentage of CLABSI per medical specialty from 2014 to 2020. The number is calculated as the count of catheter episodes which eventually experienced CLABSI at specific medical specialty at the corresponding year. The percentage is calculated using the number of catheter episodes which eventually experienced CLABSI at specific medical specialty divided by the total number of catheter episodes at the corresponding year.

#### Number of shifts of positive cultures (blood vs. non-blood samples), catheter episodes and CLABSIs per year

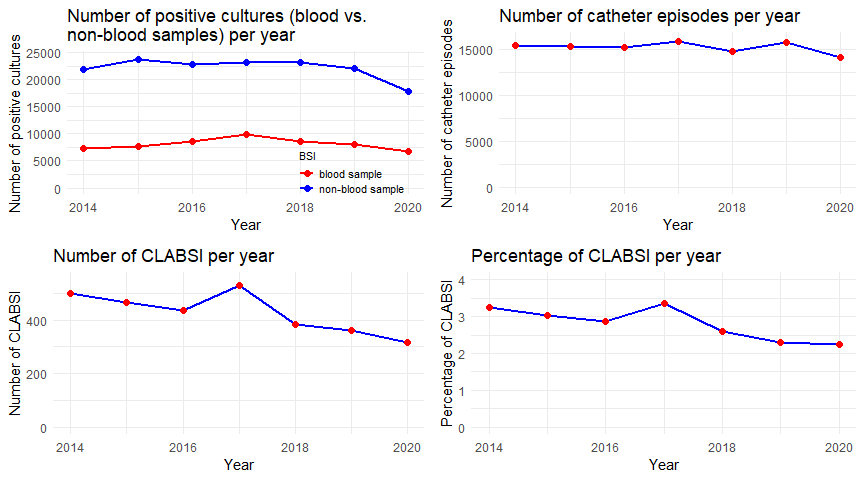

Figure 29: Number of shifts of positive cultures (blood vs. non-blood samples), catheter episode and CLABSIs per year

#### Number and percentage of positive cultures (blood vs. non-blood samples) by medical specialty per year

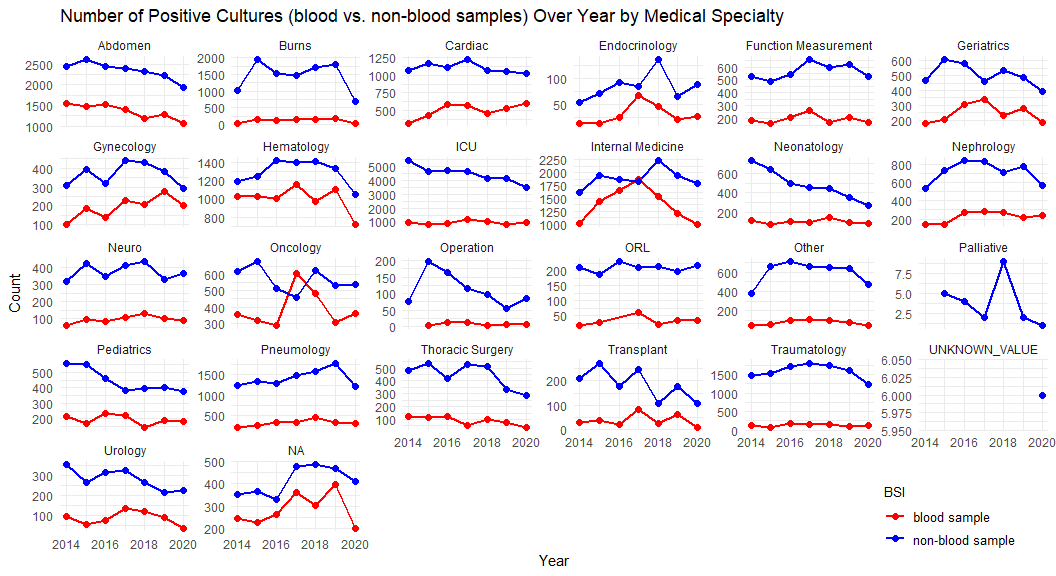

Figure 30: Number and percentage of positive cultures (blood vs. non-blood samples) by medical specialty per year. The number is calculated as the count of positive cultures (blood vs. non-blood samples) by medical specialty per year. The percentage is calculated using the number of positive cultures (blood vs. non-blood samples) by medical specialty divided by the total number of positive cultures at the corresponding year.

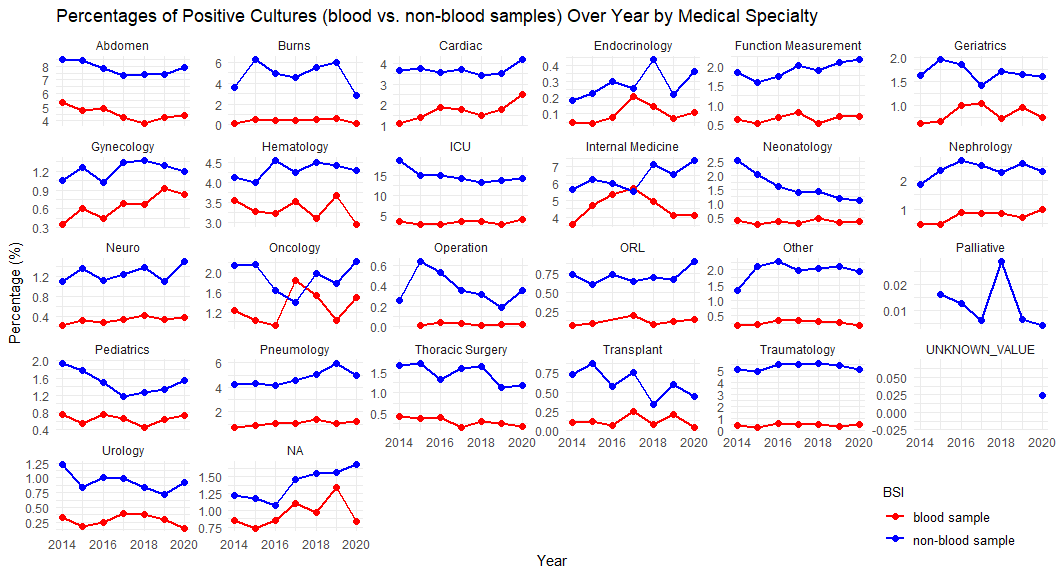

Figure 31: Number and percentage of positive cultures (blood vs. non-blood samples) by medical specialty per year. The number is calculated as the count of positive cultures (blood vs. non-blood samples) by medical specialty per year. The percentage is calculated using the number of positive cultures (blood vs. non-blood samples) by medical specialty divided by the total number of positive cultures at the corresponding year.

#### Apparent AUC performance per year across first 15 landmarks

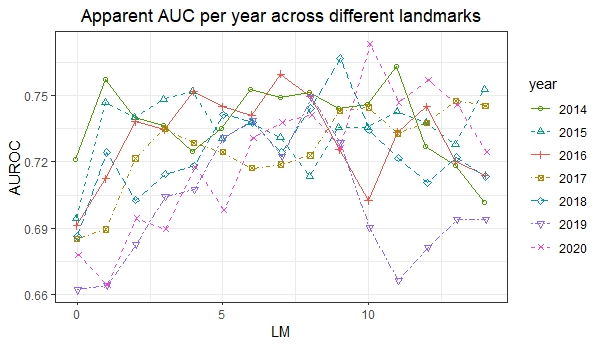

Figure 32: Apparent AUC per year from LM0 to LM14

### Supplementary material 9 - RF & XGB models details

#### Hyperparamaters

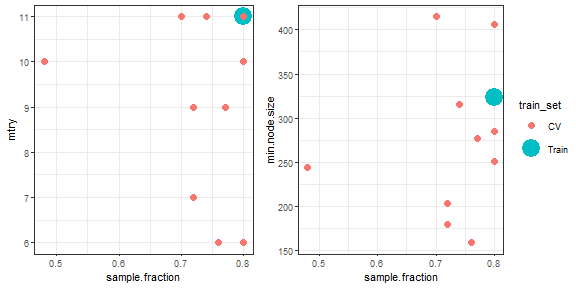

Tuned hyperparameters for RF model on the limited set of variables (for outer CV and final model)

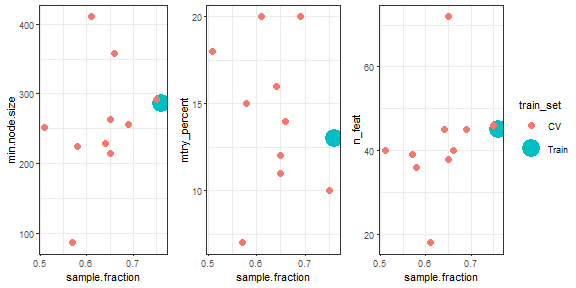

Tuned hyperparameters for RF model on the complete set of variables (for outer CV and final model)

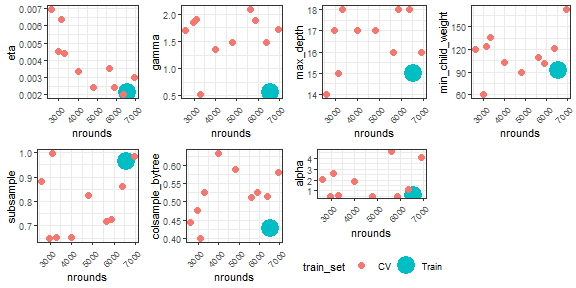

Tuned hyperparameters for XGB model on the limited set of variables (for outer CV and final model)

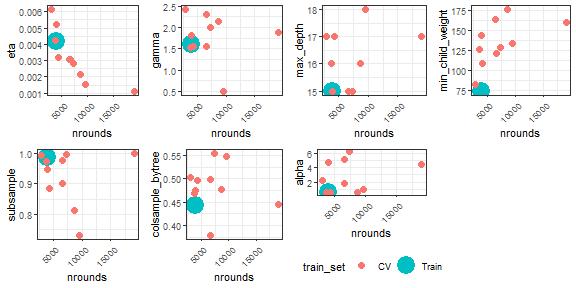

Tuned hyperparameters for XGB model on the complete set of variables (for outer CV and final model)

#### Variable importance for RF models

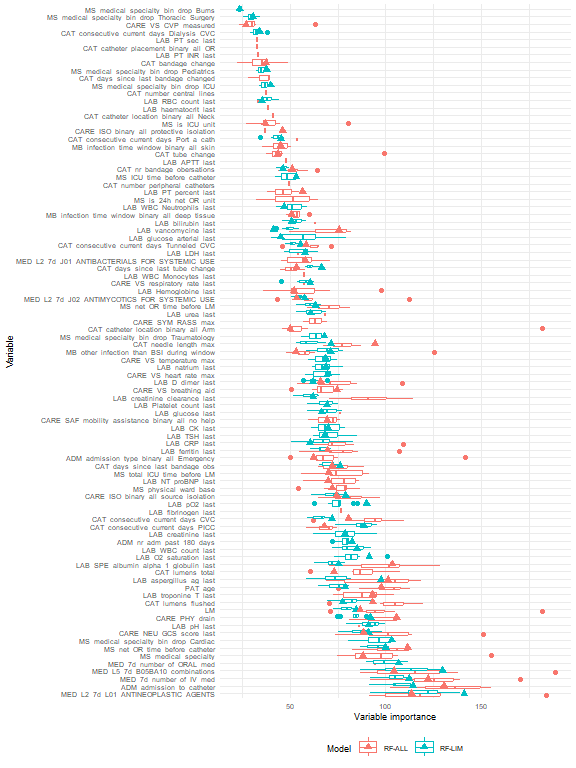

Permutation variable importance (scaled decrease in Gini index) for cross-validation (boxplots) and training set (triangle shape) for RF models

#### Variables used in final RF model on the complete set

| Variable | Variable importance |
| --- | --- |
| ADM_admission_to_catheter | 130.55385 |
| MED_7d_number_of_IV_med | 122.00353 |
| MED_L2_7d_L01_ANTINEOPLASTIC_AGENTS | 113.54341 |
| MS_net_OR_time_before_catheter | 111.29845 |
| CARE_PHY_drain | 105.71741 |
| MED_L5_7d_B05BA10_combinations | 104.29593 |
| LAB_SPE_albumin_alpha_1_globulin_last | 103.34429 |
| LAB_aspergillus_ag_last | 101.25035 |
| PAT_age | 97.46073 |
| CAT_needle_length_max | 94.39091 |
| LAB_troponine_T_last | 93.15771 |
| CAT_lumens_flushed | 93.05831 |
| CARE_NEU_GCS_score_last | 88.40497 |
| MS_medical_specialty | 88.23181 |
| LM | 86.74232 |
| CAT_consecutive_current_days_CVC | 80.49138 |
| LAB_vancomycine_last | 75.60084 |
| CARE_VS_breathing_aid | 74.73145 |
| CARE_ISO_binary_all_source_isolation | 74.22613 |
| CAT_lumens_total | 73.06975 |
| CAT_days_since_last_bandage_obs | 72.11796 |
| MS_physical_ward_base | 71.93927 |
| MS_total_ICU_time_before_LM | 70.17704 |
| LAB_NT_proBNP_last | 69.88504 |
| LAB_ferritin_last | 69.73229 |
| CARE_SAF_mobility_assistance_binary_all_no_help | 69.51147 |
| CAT_consecutive_current_days_PICC | 67.69258 |
| LAB_D_dimer_last | 66.03808 |
| ADM_admission_type_binary_all_Emergency | 62.24947 |
| CAT_consecutive_current_days_Tunneled_CVC | 58.49022 |
| MED_L2_7d_J01_ANTIBACTERIALS_FOR_SYSTEMIC_USE | 58.00482 |
| LAB_PT_percent_last | 56.69363 |
| MED_L2_7d_J02_ANTIMYCOTICS_FOR_SYSTEMIC_USE | 53.56339 |
| CAT_days_since_last_tube_change | 53.55546 |
| MB_other_infection_than_BSI_during_window | 53.11201 |
| LAB_Hemoglobine_last | 52.02352 |
| CAT_nr_bandage_obersations | 51.20850 |
| MB_infection_time_window_binary_all_deep_tissue | 50.91195 |
| CAT_catheter_location_binary_all_Arm | 50.18927 |
| CARE_ISO_binary_all_protective_isolation | 46.20432 |
| MB_infection_time_window_binary_all_skin | 45.03431 |
| CAT_tube_change | 43.82744 |
| CAT_bandage_change | 37.54254 |
| MS_is_ICU_unit | 36.93228 |
| CARE_VS_CVP_measured | 27.06831 |

Table 13: Variables selected in the final RF model on the complete set of variables, after variable selection

#### Variable importance for XGB models

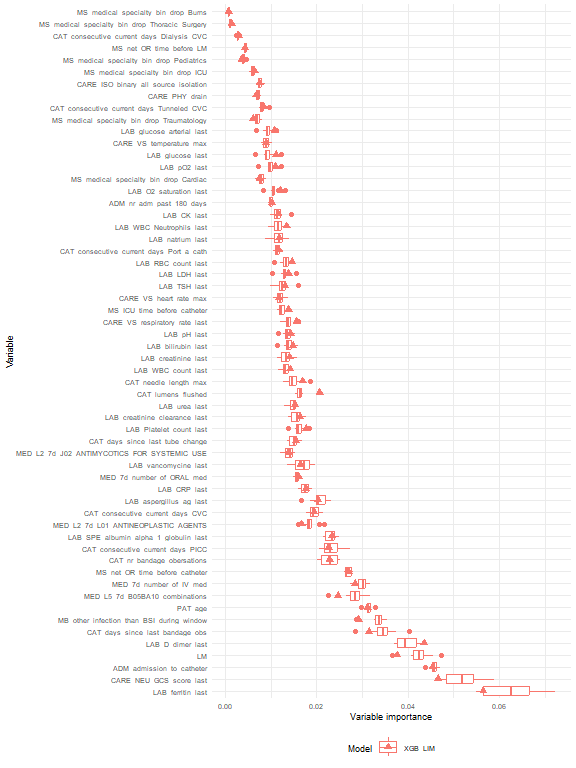

Variable importance (fractional contribution of each feature to the model based on the total gain of this feature’s splits) for cross-validation (boxplots) and training set (triangle shape) for XGB model on the limited set of variables

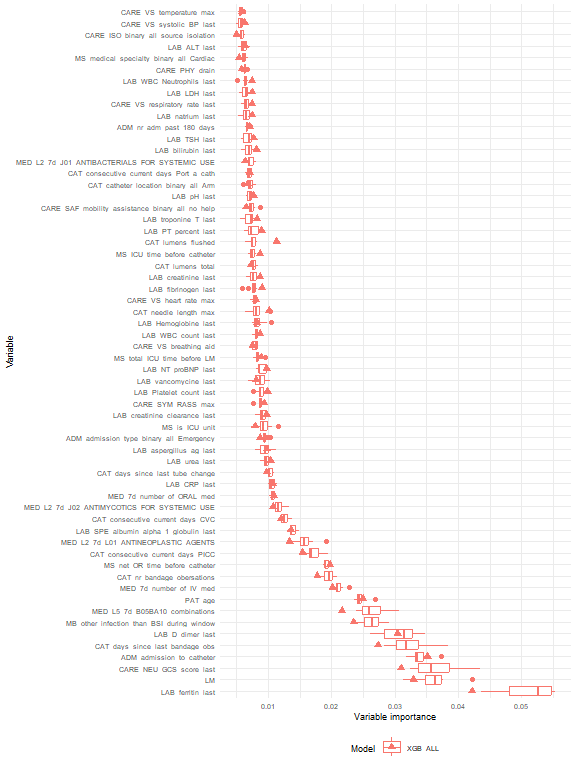

Variable importance (fractional contribution of each feature to the model based on the total gain of this feature’s splits) for cross-validation (boxplots) and training set (triangle shape) for XGB model on the complete set of variables. Only the most important 60 variables (as per the median CV variable importance) are shown.
